# Effectiveness and safety of psychosocial interventions for the treatment of cannabis use disorder: a systematic review and meta-analysis

**DOI:** 10.1101/2024.11.18.24317475

**Authors:** Monika Halicka, Thomas L Parkhouse, Katie Webster, Francesca Spiga, Lindsey A Hines, Tom P Freeman, Sabina Sanghera, Sarah Dawson, Craig Paterson, Jelena Savović, Julian PT Higgins, Deborah M Caldwell

## Abstract

**Aim:** To evaluate the effectiveness, safety, and cost-effectiveness of psychosocial interventions for cannabis use disorder (CUD).

**Design:** A systematic review of randomized controlled trials (RCTs; PROSPERO protocol CRD42024553382). We searched databases (MEDLINE/PsycInfo/Cochrane CENTRAL) to 12-Jun-2024. We assessed results using Risk of Bias 2 and conducted meta-analyses where possible.

**Setting:** Inpatient/outpatient/community-based.

**Participants:** Individuals with CUD aged ≥16 years.

**Interventions:** Psychosocial interventions lasting >4 sessions, delivered in real time.

**Measurements:** Primary outcomes were continuous- and point-abstinence, withdrawal intensity, treatment completion and adverse events.

**Findings:** We included 22 RCTs (3,304 participants). At the end of treatment, cognitive-behavioural therapy (CBT) increased point abstinence (odds ratio [OR]=18.27, 95% confidence interval [9.00; 37.07]), and continuous abstinence (OR=2.72 [1.20; 6.19]), but reduced treatment completion (OR=0.53 [0.35; 0.85]) versus inactive/nonspecific comparators. Dialectical behavioural/acceptance and commitment therapy increased point abstinence versus inactive/nonspecific comparators (OR=4.34 [1.74; 10.80]). The effect of CBT plus affect management versus CBT on point abstinence was OR=7.85 [0.38; 163.52]. The effect of CBT plus abstinence-based contingency management versus CBT on point abstinence was OR=3.78 [0.83; 17.25], and on continuous abstinence OR=1.81 [0.61; 5.41]. For CBT plus abstinence-contingency management versus CBT plus attendance-contingency management, the effect on point abstinence was OR=1.61 [0.72; 3.60], and on continuous abstinence OR=2.04 [0.75; 5.58]. The effect of community reinforcement on point abstinence was OR=0.29 [0.04; 1.90] versus CBT, and on continuous abstinence OR=47.36 [16.00; 140.21] versus nonspecific comparator. Interventions other than CBT may not affect treatment completion. No adverse events were reported. No study reported withdrawal intensity. Two economic evaluations reported higher costs for more complex psychosocial interventions and contingency management.

**Conclusions:** Cognitive-behavioural and dialectical behavioural/acceptance and commitment therapies may increase abstinence relative to inactive/nonspecific comparators. The conclusions remain tentative due to low to very low certainty of evidence and small number of studies.

## BACKGROUND

Worldwide, cannabis is the most widely used illicit drug. In 2022, the number of people aged 15-64 years using cannabis was estimated as 228 million, representing 4.4% of the global population.^1^ The risk of developing dependence on cannabis significantly increases with increasing frequency of use.^2^ The diagnosis of cannabis use disorder (CUD) outlined in the Diagnostic and Statistical Manual of Mental Disorders, Fifth Edition (DSM-5),^3^ requires the presence of at least two of eleven criteria. These include hazardous use, social or interpersonal problems related to use, neglect of major roles, withdrawal, tolerance and cravings, amongst other features. DSM-5 CUD amalgamated previous diagnoses of cannabis dependence and cannabis abuse included in DSM-IV.^4^ Cannabis dependence is also listed in the International Classification of Diseases (ICD).^5^

The prevalence of CUD has been escalating globally, affecting over 15 million men and over 8 million women in 2019.^6^ The burden of CUD is the highest among young adults aged 20-24 years.^6^ The number of people enrolling in treatment for cannabis use has been increasing globally^7^ and it is also the most frequently cited problem drug among people entering drug treatment.^1,8^ For instance, across Europe, number of adults seeking treatment increased from 27 per 100,000 in 2010 to 35 per 100,000 in 2019.^9^

According to the World Health Organization (WHO), adults using cannabis should be offered brief interventions, focused on individualized feedback and advice.^10^ However, for people diagnosed with CUD or dependence, such brief interventions have limited benefit and WHO recommend they should be referred for specialist cannabis-specific treatment.^10^ Psychosocial interventions (PSIs) are, currently, the only recommended treatment for people with CUD.^7,11,12^ This recommendation is supported by evidence from systematic reviews that suggest PSIs are effective for treatment of CUD.^13–17^ In contrast, evidence for the use of pharmacological treatments for CUD is lacking.^18^ However, these previous reviews provide limited insight into the specific types of PSIs that are most effective for treating CUD. For example, some reviews aggregate various types of PSIs for comparison against inactive controls in pairwise meta-analysis,^13,14^ some provide a descriptive summary of results from individual randomized controlled trials (RCTs),^17^ while others report an overview of findings from published systematic reviews.^19,20^

Quantitative estimates of intervention effect, safety, and cost-effectiveness are important to inform policy and clinical decision-making. However, to date, reviews have not included safety outcomes or economic evaluations of PSIs for treatment of CUD. The purpose of the present review is to provide an up-to-date and rigorous review of the evidence for clinical effectiveness, safety, and cost-effectiveness of PSIs for the treatment of CUD in adults and young people aged ≥16 years.

## METHODS

The review protocol was prospectively registered with PROSPERO (CRD42024553382).^21^ The review is reported following PRISMA guidelines.^22^

### Eligibility criteria

Study eligibility criteria are presented in **Table 1**. PSIs were grouped based on shared theoretical underpinning and the therapeutic techniques used. Intervention and comparator categories are summarised in **Table 2**, with more detail provided in Supporting Information 1. Explanation of outcome operationalization and the hierarchy of preference followed for studies reporting multiple measures and/or follow-up timepoints, are outlined in Supporting Information 2.

**Table 1.** Eligibility criteria.

| Domain | Eligibility criteria |
| --- | --- |
| Publication type | Original research reports |
|  | Trial registrations for which no linked publication could be identified were eligible only if they provided outcome data |
|  | Conference abstracts, theses, or dissertations were *not* eligible |
| Study design | Individually- or cluster-randomized controlled trials (RCTs), or the first period of cross-over RCT (prior to cross-over) |
|  | Trial-based full economic evaluations, including cost-effectiveness, cost-benefit, and cost-utility analyses |
| Population | Adults and young people aged $\geq 16$ years (on average or $>50\%$ of participants) |
|  | Diagnosis of cannabis use disorder (CUD), or cannabis dependence or abuse based on one of:<br>- recognized diagnostic criteria (e.g. any version of DSM or ICD),<br>- diagnostic cut-off on a clinically validated scale (e.g. Cannabis Abuse Screening Test),<br>- a trialist-defined level of cannabis use indicating dependence if $\geq 80\%$ of participants met the diagnostic criteria |
|  | Studies specifically recruiting participants with co-occurring schizophrenia, delirium, or psychosis, or targeting individuals with co-dependence on other substances (except or tobacco) were *not* eligible |
|  | Participants mandated to treatment by the criminal justice system were *not* eligible |
| Interventions | Any psychosocial or psychological intervention for the treatment of CUD |
|  | Lasting more than four sessions or at least four weeks, if the number of sessions was unclear (brief interventions were *not* eligible) |
|  | Delivered synchronously (in real-time), without restrictions on qualification or profession of the person delivering the intervention |
|  | Individual or group-based |
|  | Asynchronous interventions, peer support, or multi-aid programmes were *not* eligible |
|  | Studies in which the same psychosocial intervention was an adjunct to an ineligible intervention were *not* included, as such studies do not contribute relevant comparisons for this review (e.g., a pharmacological plus a psychosocial intervention compared with a psychosocial intervention alone) |
| Comparators | Inactive or nonspecific intervention; or other active psychosocial or pharmacological intervention, alone or in combination |
| Outcomes and timepoints | Primary outcomes:<br>(1) point abstinence at end of treatment<br>(2) continuous abstinence at the end of treatment<br>(3) intensity of withdrawal and/or craving<br>(4) completion of scheduled treatment<br>(5) adverse events at any time<br>(6) dropout due to adverse events |
|  | Secondary outcomes:<br>(1) point abstinence at medium follow-up (up to 6 months post-treatment) and long follow-up (over 6 months post-treatment)<br>(2) continuous abstinence at medium and long follow-up<br>(3) duration of the longest continuous abstinence during treatment, medium, and long follow-up (not pre-specified)<br>(4) frequency and quantity of use at the end of treatment, medium, and long follow-up<br>(5) number of participants engaging in further treatment at any time post-treatment<br>(6) economic outcomes |
| Setting | Inpatient, outpatient, or community-based treatment setting |
|  | Studies set in residential research laboratories were *not* eligible |

**Table 2.** Intervention and comparator categories.

| Category | Description |
| --- | --- |
| 'CBT' | Interventions using cognitive-behavioural therapy (CBT) techniques such as cognitive restructuring and skills training, in the context of substance use commonly combined with motivation enhancement, also including relapse prevention |
| 'CBT-affect' | CBT techniques combined with affect management |
| 'DBT/ACT' | Third/fourth-wave psychotherapies such as dialectical behavioural therapy (DBT) or acceptance and commitment therapy (ACT), using psychoeducation, mindfulness, emotion regulation, skills training, and acceptance |
| 'CM-abstinence' | Abstinence-based contingency management (CM), where participants receive rewards (e.g. lottery draws or vouchers) for providing urine specimens negative for cannabinoids |
| 'CM-attendance' | Attendance-based CM, where participants receive rewards for attending intervention sessions, providing urine samples (regardless of the results), or completing homework assignments |
| 'MDFT' | Multidimensional family therapy (MDFT) focusing on improving functioning across multiple domains and systems, from intrapersonal, through parenting and family environment, to community systems |
| 'Community reinforcement' | Using existing community resources and developing new support systems to rearrange environmental contingencies for supporting abstinence |
| 'Inactive/nonspecific' comparator | Waitlist or no intervention controls, where participants do not receive any treatment, at least until the end of the waitlist period (inactive comparators); or interventions aiming to control for the common features of therapies such as support or educational content but not including training in techniques thought of as being therapeutic (nonspecific comparators) |

### Searches

We searched Ovid MEDLINE-ALL and PsycInfo, and the Cochrane Central Register of Controlled Trials (CENTRAL) in the Cochrane Library using relevant subject headings, text-words and search syntax appropriate to each resource (all available years to 12-Jun-2024). Reports of RCTs from Embase and CINAHL were captured via our search of CENTRAL.^23^ To identify potentially relevant economic evaluations, we ran separate searches in Ovid MEDLINE and Embase (all available years to 30 July 2024). Search strategies are provided in Supporting Information 3.

### Study selection

Titles and abstracts were screened independently by at least two reviewers using the Rayyan platform.^24^ Potentially relevant texts were retrieved in full, and assessed independently by at least two reviewers using the LaserAI platform.^25^ Discrepancies were resolved by discussion with a third reviewer, or the wider review team.

### Data extraction

Using piloted, standardized forms created in LaserAI,^25^ we extracted details on study design and conduct, eligibility criteria, participant demographics and PROGRESS-Plus characteristics,^26^ intervention and comparator details, and outcome data. We used LaserAI’s AI-enhanced suggestions to support extraction of study characteristics.^25^ However, all suggestions were verified and amended, if needed, by a reviewer.

Arm-level numerical data for dichotomous outcomes were extracted as the number of participants with event, number with available outcome data, and number randomized into each arm (the denominator used in the analysis depended on outcome – see synthesis of results). For continuous outcomes, mean (M) with standard deviation (SD) and number of participants analysed in each arm were extracted for end of treatment and follow-up. Data processing steps in preparation for synthesis are outlined in Supporting Information 4.

Study characteristics and numerical data were extracted by a single reviewer and checked in detail by a second reviewer. Discrepancies were resolved through discussion, or with a third reviewer.

### Risk of bias assessment

We assessed risk of bias (RoB) using the RoB2^27^ tool at the outcome level for each study, for all primary and secondary effectiveness outcomes reported at the end of treatment, and safety outcomes related to adverse events at any time. RoB2 was assessed initially by two reviewers independently. Once consistency had been achieved, RoB2 was assessed by one reviewer and checked by a second. Two reviewers independently assessed economic outcomes using the Drummond and Jefferson critical appraisal checklist.^28^

### Synthesis of results

Pairwise random-effects meta-analyses were conducted in R software version 4.3.1, using ‘meta’ package version 7.0-0.^29–31^ Effect estimates were pooled if there were at least two studies contributing data for the same comparison; otherwise, study-level effect estimates are presented. Statistical heterogeneity was assessed using the I^2^ statistic to quantify inconsistency, with strength of evidence quantified using the p-value from the chi^2^ test. The between-study variance, τ^2^, was estimated using the restricted maximum-likelihood (REML) method. We assumed a common τ^2^ across all comparisons within the same outcome and timepoint (as is done in a network meta-analysis).^32^ This was because there were too few studies to estimate τ^2^ reliably within each comparison^33^ and we had no reason to expect that the between-study variance would differ across comparisons. To estimate a common τ^2^, we used residual τ^2^ from a meta-regression with comparison included as a covariate. A fixed continuity correction (0.5) was added to studies with zero events in one arm. We present the results as odds ratios (ORs) for dichotomous outcomes and ratios of means (RoMs) for continuous outcomes,^34–36^ with 95% confidence intervals (CIs).

Primary meta-analyses were conducted at the end of treatment. Additional timepoints were medium (≤6 months) and long follow-up (>6 months post-treatment). Analyses were based on the number of participants with available outcome data, except for completion of treatment which was based on the number of participants randomized. To minimize multiplicity of analyses (e.g. by reusing the same intervention arms across different comparisons), the comparisons reported in the Results section were prioritised for synthesis, and study-level effect estimates for other potential comparisons are presented in Supporting Information 5.

#### Sensitivity and subgroup analyses

Sensitivity analyses addressed fixed-effect meta-analyses, and imputing missing outcome data as abstinent or non-abstinent for dichotomous abstinence outcomes. We planned subgroup analyses to explore heterogeneity using the following potential effect modifiers: intensity and duration of cannabis use, mental health co-morbidities, intervention intensity, treatment setting, use of adjunct interventions or booster sessions, and PROGRESS-Plus characteristics.^26^

### Certainty of evidence

We used the GRADE framework^37^ to assess the certainty of evidence for effectiveness of PSIs on primary and secondary outcomes at the end of treatment. There are no established thresholds representing minimal clinically important differences for these outcomes. In this review, we describe intervention effects as clinically meaningful if they represent a 10% increase or reduction of risk in the intervention group relative to the comparator for dichotomous outcomes. For continuous outcomes effects are assessed as clinically meaningful if an intervention halves or doubles the frequency, quantity, or duration of outcome relative to a comparator. The criteria considered for grading the certainty are outlined in Supporting Information 6.

## RESULTS

### Included studies

Thirty-two reports of k=22 studies (participants n=3,304) were included (**Figure 1**). Details of excluded reports are listed in Supporting Information 7.

**Figure 1.**
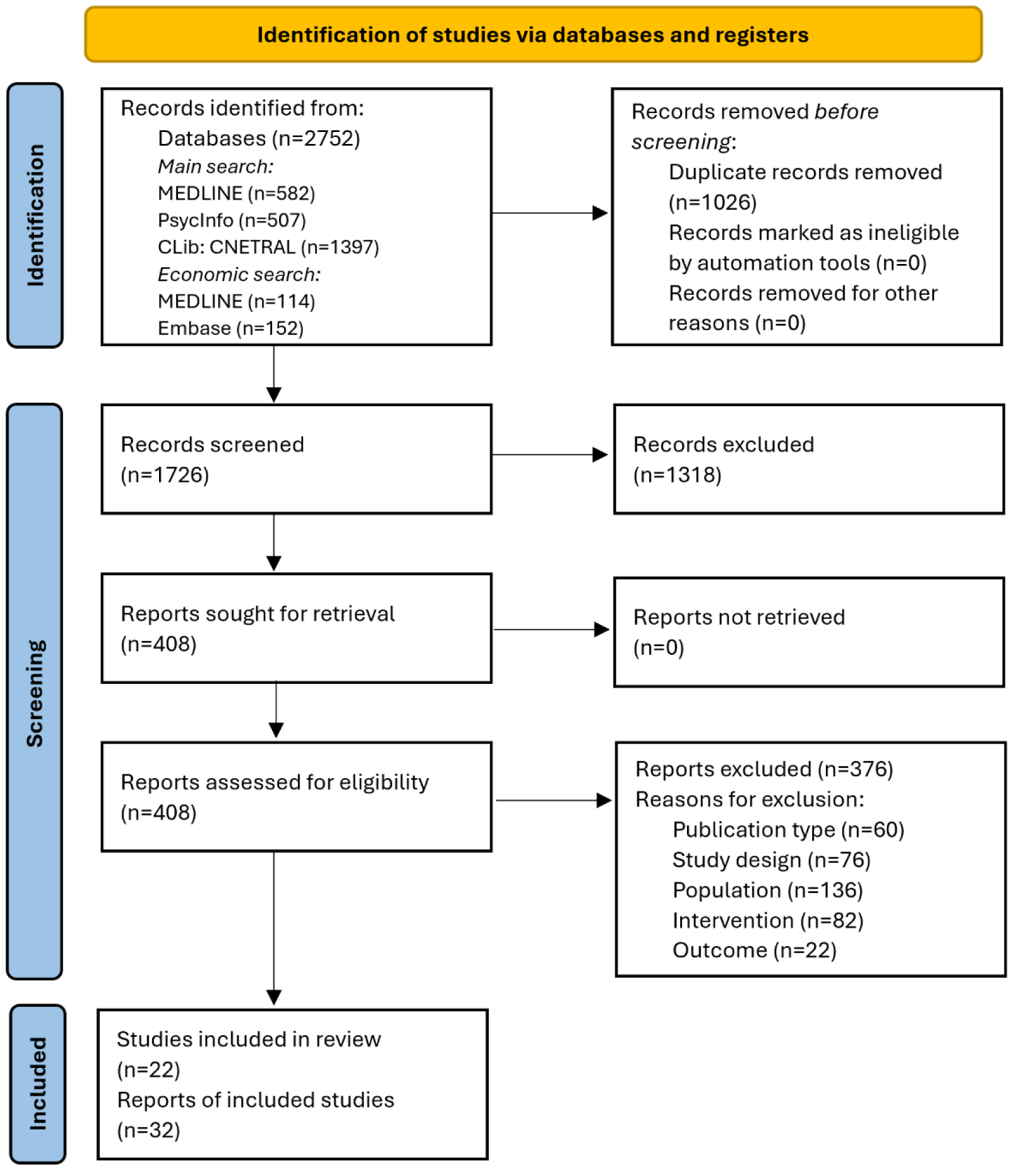
PRISMA flow diagram. Citation searching did not identify any additional records, therefore, identification of studies via other methods is not displayed.

Characteristics of included studies are presented in **Table 3**. Study sample size ranged from 40 to 450 participants (M=150, SD=103). Most studies were conducted in the United States (k=15) and in outpatient settings (k=15). Participants were mostly male (M=80%, range 56-100%) and of white ethnicity. Mean age ranged from 16 to 48 years (M=28, SD=8). Education ranged from secondary school to degree-level (k=10) or mean 13.5 years of education (k=7), most participants were employed (k=12), and on average 36% were married or co-habiting (k=10). Several studies excluded individuals with excessive commuting distance/transportation difficulties or unstable living situations (k=5), those with serious mental health issues (k=14), or those who required inpatient treatment or had serious medical problems (k=8). A detailed summary of the PROGRESS-Plus^26^ characteristics relating to equity is presented in Supporting Information 8. Participants met diagnostic criteria for CUD (k=5), cannabis dependence (k=10), abuse (k=1), dependence *or* abuse (k=4), or a diagnostic screening cut-off (k=2). On average, participants used cannabis on 74% of days (SD=21%, k=15).

**Table 3.** Characteristics of included studies.

| Study ID<sup>a</sup>; country | Total number randomized<sup>b</sup> (per arm) | CUD characteristics (percentage or mean [SD]) | Duration and intensity of cannabis use (mean [SD]) | Age in years (mean [SD], range); Sex (%) | Interventions<sup>c</sup> (duration, frequency) | Outcome(s) (timepoints<sup>d</sup>) | Notes |
| --- | --- | --- | --- | --- | --- | --- | --- |
| Babor 2004<sup>51</sup><br>United States | 304 (148, 156) | 100% DSM-IV diagnosis of current cannabis dependence; N of dependence symptoms 5.59 (1.25); N of abuse symptoms 2.09 (0.81); MPS 9.27 (3.52) | Age at first use 18.21 (4.95); years of regular use 17.9 (NR); joints per day 2.78 (2.27); ounces per week 0.40 (0.46); days of use 88.74% (15.74), using on at least 40/90 past days | 36.45 (8.45), 18-62<br>71% male | 1. Wait;<br>2. MET/CBT;<br>(4 months, 1 x week-4 weeks) | Continuous abstinence (end)<br>Completion of treatment (end)<br>Level of cannabis use (frequency and quantity; end) | Excluded participants with dependence on alcohol or other drugs |
| Buckner 2019<sup>43</sup><br>United States | 55 (28, 27) | 100% met DSM-5 diagnostic criteria for CUD; MPS 12.97 (6.79) | Age at first use 16.07 (3.36); years of use 6.73 (7.63); joints in past month 64.14 (52.63) | 23.15 (7.38), 18-65*<br>56% male | 1. MET/CBT;<br>2. ICART;<br>(12 weeks, 1 x week) | Point abstinence (end)<br>Completion of treatment (end)<br>Level of cannabis use (quantity; end) | Participants all met DSM-5 diagnosis for CUD and an anxiety disorder; 25.5% diagnosed with alcohol use disorder |
| Budak 2024<sup>56</sup><br>Turkey | 70 (35, 35) | 100% DSM-IV diagnosis of substance abuse | NR | 18-28 = 51.6%;<br>29-39 = 35%;<br>40-50 = 13.3%<br>100% male | 1. Wait;<br>2. Mind/Edu;<br>(4 weeks, 2 x week) | Completion of treatment (end) | Excluded participants using substances other than cannabis |
| Budney 2000<sup>44</sup><br>United States | 40 (20, 20) | 100% met DSM-III-R diagnostic criteria for current cannabis dependence; N of DSM-III dependence criteria 6.45 (2.22); Addiction severity index composite score for drug domain 0.21 (0.01) | Years of cannabis use 15.1 (8.68); days use per month 22.25 (8.79) | 32.85 (8.52), ≥18*<br>85% male | 1. MET/BT;<br>2. MET/BT/CM-ab;<br>(14 weeks, 1-2 x week) | Continuous abstinence (end)<br>Point abstinence (end)<br>Duration of continuous abstinence (end)<br>Level of cannabis use (frequency; end) | Excluded participants with dependence on alcohol or any other drug except nicotine; 30% diagnosed with antisocial personality disorder |
| Budney 2006<sup>47</sup><br>United States | 90 (30, 30, 30) | 100% met DSM-IV diagnostic criteria for current cannabis dependence; N of DSM-IV criteria for cannabis dependence 4.87 (1.39); MPS 7.83 (4.36) | Years of regular use 13.77 (9.34); times used per day 3.90 (2.48); days used in prior 30 days 25.60 (7.16) | 33.1 (10.3), ≥18*<br>77% male | 1. CM-ab;<br>2. MET/CBT/CM-at;<br>3. MET/CBT/CM-ab;<br>(14 weeks, 1-2 x week) | Continuous abstinence (end; medium, 6 months; long, 12 months)<br>Point abstinence (end; medium, 6 months; long, 12 months);<br>Completion of treatment (end) | Excluded participants with dependence on alcohol or any other drug except nicotine |
|  |  |  |  |  |  | Level of cannabis use (frequency; end; medium, 6 months; long, 12 months)<br>Duration of continuous abstinence (end) |  |
| Carroll 2006<sup>55,61,77</sup><br>United States | 136 (33, 34, 36, 33) | 100% met DSM-IV diagnostic criteria for current cannabis dependence | Age of first use 14; days used in prior 28 days 13 (10.3) | 21 (2.1), 18-25*<br>90% male | 1. NS;<br>2. CM-ab-at/NS;<br>3. MET/CBT;<br>4. MET/CBT/CM-ab-at;<br>(8 weeks, 1 x week) | Completion of treatment (end)<br>Continuous abstinence (medium, 6 months)<br>Point abstinence (medium, 6 months)<br>Duration of continuous abstinence (end)<br>Level of cannabis use (frequency; end; medium, 6 months)<br>Cost-effectiveness (end; medium, 6 months) | All participants referred by probation service; 5% met criteria for current DSM-IV alcohol use disorder (24.4% lifetime); other lifetime diagnoses: 11% depressive disorder, 22% anxiety disorder, 43% antisocial personality disorder |
| Carroll 2012<sup>58</sup><br>United States | 127 (27, 36, 32, 32) | 100% met DSM-IV diagnostic criteria for current cannabis dependence; Addiction Severity Index cannabis composite score 0.31 (0.26) | Years of cannabis use 10.5 (7.3); days used in prior 28 days 16.4 (9.7) | 25.7 (7.1), ≥18*<br>84% male | 1. CM-ab;<br>2. MET/CBT;<br>3. MET/CBT/CM-at;<br>4. MET/CBT/CM-ab;<br>(12 weeks, 1 x week) | Completion of treatment (end)<br>Level of cannabis use (frequency; end; medium, 6 months; long, 13 months)<br>Duration of continuous abstinence (end) | 93.7% referred by criminal justice system; excluded participants with dependence on other drugs/alcohol; other lifetime diagnoses: 4.7% major depressive disorder, 12.6% anxiety disorder, 25.2% antisocial personality disorder |
| Copeland 2001<sup>78,79</sup><br>Australia | 147 (69, 78) | 96.4% met DSM-IV diagnostic criteria for cannabis dependence; 100% were dependent according to the SDS; SDS score 9.25 (2.92) | Years of regular cannabis use 13.9 (7.0); median age at first use 15 (range 7-45); median age at first regular use 18 (range 11-47); median 8 waterpipes a day | 32.3 (7.9), 18-59<br>69% male | 1. Wait;<br>2. MET/CBT;<br>(6 weeks, 1 x week) | Continuous abstinence (medium, 24 weeks)<br>Level of cannabis use (frequency; medium, 24 weeks) | Excluded participants who reported more than weekly use of drugs other than cannabis, nicotine, or alcohol in the past six months, or with a score >15 |
|  |  |  | (range 0.1 – 125); all smoking for at least 3 days per week |  |  | Level of cannabis use (quantity; medium, 24 weeks) | on the Alcohol Use Disorders Identification Test |
| Davoudi 2021a<sup>41</sup><br>Iran | 61 (31, 30) | 100% psychiatrist diagnosis of CUD | Months of cannabis use 18.49 (6.01) | 26.41 (6.65), 18-45*<br>100% male | 1. NS;<br>2. DBT;<br>(12 weeks, 1 x week) | Completion of treatment (end)<br>Point abstinence (end; medium, 2 months)<br>Level of cannabis use (frequency; end + 4 weeks; medium, 2 months)<br>Craving (end; medium, 2 months) | Excluded participants consuming methamphetamine, amphetamine, cannabis, methadone, benzodiazepines, or morphine during the research stages |
| Davoudi 2021b<sup>42</sup><br>Iran | 50 (25, 25) | 100% DSM-5 diagnosis of CUD | Months of cannabis use 23.7 (6.84) | 25.85 (4.99), 18-45*<br>100% male | 1. NS;<br>2. ACT;<br>(12 weeks, 1 x week) | Point abstinence (end; medium, 3 months)<br>Completion of treatment (end)<br>Level of cannabis use (frequency; end; medium, 2 months) | Excluded participants who used other drugs during the intervention and follow-up stages of the research; all participants had a score of $\geq 13$ on the Beck depression and anxiety inventory |
| Hoch 2014<sup>40</sup><br>Germany | 279 (130, 149) | 87.1% lifetime (56% past 4 weeks) ICD-10 diagnosis of cannabis dependence; ICD-10 N of symptoms: lifetime 4.9 (2.0); past 4 weeks 3.3 (2.3) | Age at onset of cannabis use 15.2 (3.7); age at first regular use 18.8 (6); days use over past 4 weeks 18.8 (9.7) | 26.6 (8.2), 16-63<br>87% male | 1. Wait;<br>2. MET/CBT;<br>(10 weeks, NR) | Point abstinence (end) | Excluded participants with ICD-10 dependence on alcohol or any other illicit drug (apart from cannabis) |
| Kadden 2007<sup>48,80</sup><br>United States | 240 (62, 54, 61, 63) | 100% DSM-IV diagnosis of cannabis dependence; MPS 13.88 (6.75) | Joints per day 4.5 (4.93); days of use 89% (15) | 32.7 (9.6), $\geq 18^*$<br>71% male | 1. NS;<br>2. CM-ab;<br>3. MET/CBT;<br>4. MET/CBT/CM-ab;<br>(9 weeks, 1 x week) | Continuous abstinence (end; medium, 6 months; long, 12 months)<br>Completion of treatment (end)<br>Duration of continuous abstinence (end; long, 12 months)<br>Level of cannabis use (frequency; end; medium, | Excluded participants with dependence on alcohol or other drugs |
|  |  |  |  |  |  | 6 months; long, 12 months) |  |
| Kaminer 2017<sup>46</sup> **<br>United States | 75 (40, 35) | 100% DSM-IV diagnosis of current CUD (i.e. cannabis dependence or abuse) | NR | 16.11 (NR), 13-18<br>83% male | 1. MET/CBT;<br>2. ComReinf;<br>(10 weeks, 1 x week) | Point abstinence (end)<br>Completion of treatment (end) | Excluded participants with any substance dependence criteria other than nicotine or alcohol |
| Khalily 2023<sup>54</sup><br>Pakistan | 120 (60, 60) | 100% attained a score on the CAST instrument of >2 for cannabis abuse; SDS 10.87 (2.59) | NR | 24.7 (3.4), 18-30*<br>95% male | 1. NS;<br>2. ComReinf;<br>(6 weeks, 1 x week) | Continuous abstinence (end; medium, 18 weeks; long, 30 weeks)<br>Completion of treatment (end) | Excluded participants meeting DSM-5 criteria for misuse of other psycho-active substances including alcohol |
| Litt 2013<sup>49</sup><br>United States | 215 (71, 73, 71) | 100% DSM-IV diagnosis of cannabis dependence or abuse; MPS 16.28 (6.76) | Joints per day 1.8 (2.8);<br>days used in prior 90 days 70.9 (29.1) | 32.7 (10), ≥18*<br>68% male | 1. NS;<br>2. MET/CBT/CM-ab;<br>3. MET/CBT/CM-at;<br>(2 months, 1 x week) | Continuous abstinence (end; medium, 6 months; long, 12 months)<br>Level of cannabis use (frequency; end; medium, 6 months; long, 12 months)<br>Duration of continuous abstinence (end) | Excluded participants dependent on drugs other than cannabis or nicotine |
| Litt 2020<sup>50</sup><br>United States | 198 (49, 51, 48, 50) | 100% DSM-IV diagnosis for cannabis dependence (corresponds to DSM-V diagnoses of moderate-severe CUD) | Grams per day 2.06 (2.32);<br>days used cannabis in prior 90 days 81.8 (13.7) | 36 (12), ≥18*<br>58% male | 1. MET/CBT;<br>2. MET/CBT/CM-ab;<br>3. MET/CBT;<br>4. MET/CBT /CM-ab;***<br>(12 weeks, 1-2 x week) | Continuous abstinence (end; medium, 6 months; long, 12 months)<br>Completion of treatment (end)<br>Level of cannabis use (frequency; end; medium, 6 months; long, 12 months)<br>Duration of continuous abstinence (end) | Participants could meet criteria for dependence on other substances, but must have reported that marijuana was their primary substance of abuse |
| NCT02102230 2014<sup>39</sup><br>United States | 111 (41, 42, 28) | 100% met DSM-5 diagnostic criteria for CUD | Cannabis use episodes per week 10.3 (12.0) | 48.34 (15.83), 19-64<br>95% male | 1. NS;<br>2. CBT-I;<br>3. CBT-I;*** | Point abstinence (end + 2 weeks; medium, 6 months) | Participants were veterans recruited through a Veterans Affairs outpatient substance abuse treatment program; |
|  |  |  |  |  | (6 weeks, 1 x week) | Adverse events (medium, 6 months)<br>Completion of treatment (end)<br>Level of cannabis use (frequency; end; medium, 6 months) | the trial was terminated but the data collected up to the point of termination was available from the registration record |
| Rigter 2013<sup>59,60,81-83</sup><br>Belgium, France, Germany, Netherlands, Switzerland | 450 (238, 212) | 84% DSM-IV diagnosis for cannabis dependence (at least 3 of 7 dependence criteria met); 16% DSM-IV diagnosis for cannabis abuse (at least 1 of 4 abuse criteria met) | Days used in past 90 days 60.70 (25.34) | 16.3 (1.2), 13-18*<br>85% male | 1. MET/CBT;<br>2. MDFT;<br>(6 months, 2 x week) | Level of cannabis use (frequency; end; medium, 6 months)<br>Cost-utility (long, 12 months)<br>Cost-effectiveness (long, 12 months) | 40% of participants had an AUD; <5% had substance use disorders for other drugs |
| Stanger 2009<sup>45</sup><br>United States | 69 (33, 36) | 45% DSM-IV diagnosis for cannabis abuse; 43% DSM-IV diagnosis for cannabis dependence | Uses per day 1.8 (1.4); days used in previous month 13.3 (10.3) | 16 (1.05), 12-18*<br>83% male | 1. MET/CBT/CM-at/NS;<br>2. MET/CBT/CM-ab/NS;<br>(14 weeks, 1-2 x week) | Continuous abstinence (end)<br>Point abstinence (end; medium, 6 months; long, 9 months)<br>Completion of treatment (end)<br>Level of cannabis use (frequency; end; medium, 6 months; long, 9 months)<br>Duration of continuous abstinence (end)<br>Adverse events (long, 9 months) | Other DSM-IV diagnoses: 22% alcohol abuse, 1.4% opiate abuse, 1.4% sedative abuse; endorsed by a parent: 59% ODD/CD, 48% ADHD, 43% major depression and/or GAD; endorsed by youth: 26% ODD/CD, 26% ADHD, 17% major depression and/or GAD |
| Stephens 1994<sup>53,84</sup><br>United States | 212 (106, 106) | 89% scoring above the diagnostic cut-point of 5 on the DAST; DAST 8.88 (2.86) | Age at first use 16.17 (4.25); age at daily use 19.94 (5.55); years of cannabis use 15.39 (5.06); days used in past 90 days 80.67 (15.47) | 31.91 (NR), 18-65<br>76% male | 1. NS;<br>2. RelPrev;<br>(12 weeks, 1 x week-2 weeks) | Continuous abstinence (end; medium, 6 months; long, 12 months)<br>Level of cannabis use (frequency; end + 1 month; medium, 6 months; long, 12 months) | Excluded participants dependent on alcohol or other drugs; included booster sessions 3 and 6 months post-treatment |
| Stephens 2000<sup>52</sup><br>United States | 203 (86, 117) | 98% DSM-III-R diagnosis of cannabis dependence; dependence symptoms 6.74 (1.97) (out of 9); N of marijuana-related problems 9.88 (2.97) (out of 11) | Age of first use 15.93 (3.90); age of first daily use 19.60 (5.6); years of cannabis use 17.35 (5.21); days used over past 90 days 74.64 (18.54) | 34 (6.85), NR<br>77% male | 1. Wait;<br>2. RelPrev;<br>(4 months, 1 x week-2 weeks) | Continuous abstinence (end)<br>Completion of treatment (end)<br>Level of cannabis use (frequency; end) | Excluded participants with alcohol or other drug abuse |
| Wolitzky-Taylor 2022<sup>57</sup><br>United States | 52 (25, 27) | 100% met MINI diagnostic criteria for CUD; 1.9% mild, 7.7% moderate, 90.4% severe CUD; CAST 4.46 (1.61) | Days used in past 30 days 21.81 (8.71) | 22.16 (1.98), 18-25*<br>58% male | 1. MET/CBT;<br>2. AMT;<br>(12 weeks, 1 x week) | Completion of treatment (end)<br>Level of cannabis use (frequency; end; medium, 3 months) | Excluded participants whose primary substance of dependence was not cannabis; other MINI diagnoses: 48% AUD, 6% non-CUD SUD, 63% GAD, 40% social anxiety disorder, 15% panic disorder, 25% agoraphobia, 27% OCD, 23% PTSD |

<sup>a</sup>References include related articles from which additional relevant information was extracted or used for risk of bias assessment; <sup>b</sup>Only includes participants randomized to eligible study arms; <sup>c</sup>Only includes intervention arms eligible for the current review; <sup>d</sup>For end of treatment assessment ('end'), see intervention duration; for medium- and long-term assessments, months from end of treatment;
\*Represents trial eligibility criteria rather than actual characteristics of included participants.
\*\*Participant characteristics are an approximation based on those included in phase 1 of the study, while only phase 2 was relevant for the current review.
\*\*\*Arms 1 & 3, and arms 2 & 4 from Litt 2020, and arms 2 & 3 from NCT02102230, were pooled for synthesis.
ACT, acceptance and commitment therapy; ADHD, attention deficit hyperactivity disorder; AMT, affect management treatment; AUD, alcohol use disorder; CAST, Cannabis Abuse Screening Test; CBT, cognitive-behavioural therapy; CBT-I, cognitive-behavioral therapy for insomnia; CD, conduct disorder; CM-ab, contingency management-abstinence; CM-at, contingency management-attendance; ComReinf, community reinforcement; CUD, cannabis use disorder; DAST, Drug Abuse Screening Test; DBT, dialectical behavioural therapy; DSM, Diagnostic and Statistical Manual of Mental Disorders; GAD, generalized anxiety disorder; IATP, individualized assessment and treatment program; ICART, integrated cannabis and anxiety reduction treatment; ICD, International Statistical Classification of Diseases and Related Health Problems; MDFT, multidimensional family therapy; MET/BT, motivational enhancement/behavioural therapy; MET, motivation enhancement therapy; Mind/Edu, mindfulness-based psychoeducation; MINI, Mini International Neuropsychiatric Interview, MPS, Marijuana Problems Scale; N, number; NR, not reported; NS, nonspecific comparator; OCD, obsessive-compulsive disorder; ODD, oppositional-defiant disorder; PTSD, post-traumatic stress disorder; RelPrev, relapse prevention; SD, standard deviation; SDS, Severity of Dependence Scale; SUD, substance use disorder; Wait, waitlist comparator.

CBT was the most commonly evaluated PSI (k=15), followed by abstinence-based contingency management (CM-abstinence; k=8), attendance-based CM (CM-attendance; k=4), dialectical behavioural/acceptance and commitment therapies (DBT/ACT; k=3), CBT with affect management (CBT-affect; k=2), community reinforcement (k=2), and multidimensional family therapy (MDFT; k=1). Inactive/nonspecific comparators were used in k=13 studies. Interventions were delivered over 1-6 months (M=2.77, SD=1.09) and consisted of 6-52 sessions (M=13.84, SD=10.81). Most sessions occurred weekly (60%). All interventions were delivered in person, either individually (61%), as a group (19%), or mixed (10%) format (10% were not reported).

Included studies reported point- (k=10), and continuous-abstinence (k=12), duration of continuous abstinence (k=8), completion of treatment (k=16), frequency (k=17) and quantity of cannabis use (k=3), craving (k=1), adverse events (k=2), and cost-effectiveness outcomes (k=2) at any timepoint. No studies reported on intensity of withdrawal, engagement in further treatment, or dropout due to adverse events.

### Risk of bias

Across all effectiveness outcomes at the end of treatment and safety outcomes, we judged 70% to be at high RoB, 21% to have some concerns, and only 9% to be at low risk of bias. The main concerns were bias in selection of the reported result (e.g. lack of pre-specified analysis plan), bias due to missing outcome data (e.g. high attrition likely dependant on participant relapse), and bias in measurement of outcome (e.g. self-report by unblinded participants). Supporting Information 9 includes detailed assessments for each RoB2 domain.

### Results synthesis

The effectiveness results reported below are for the end of treatment timepoint only (4-24 weeks). Results for medium and long follow-up timepoints are reported in Supporting Information 10. Subgroup analyses were not possible due to an insufficient number of studies, but the relevant characteristics are reported in **Table *3***. Sensitivity analyses are reported in Supporting Information 11. Summary of findings tables are presented in Supporting Information 12. Interpretation of findings is based on minimally important clinical differences, and takes into account the GRADE assessments of certainty of evidence. Interpretations are not based on statistical significance.^38^

#### Point abstinence

Nine studies^39–47^ were included in the analysis for point abstinence. Seven measured abstinence using urine tests, one with self-report,^39^ and one used both.^46^ Two studies defined point abstinence as seven days of abstinence.^39,40^ Evidence of effectiveness is of very low certainty due to concerns over high RoB and imprecision, and for some comparisons also due to indirectness (Supporting Information 12). The common τ^2^ was estimated as 0.00 (standard error, SE=0.38). Meta-analyses included a maximum of two studies per comparison (**Figure 2**). CBT relative to a waitlist comparator (OR=18.27, 95% CI [9.00; 37.07]), and DBT/ACT relative to a nonspecific comparator (OR=4.34 [1.74; 10.80]), may lead to clinically meaningful increases in point abstinence. CBT plus CM-abstinence may improve abstinence compared with CBT (OR=3.78 [0.83; 17.25]) but the CIs are also consistent with a decrease in abstinence (i.e. favouring CBT). There is little to no evidence of an effect of CM-abstinence relative to CM-attendance when both are delivered with CBT (OR=1.61 [0.72; 3.60]). Community reinforcement may be associated with a meaningful *decrease* in abstinence when compared with CBT (OR=0.29 [0.04; 1.90]), although the CIs are also consistent with an increase in abstinence. The comparison of CBT-affect relative to CBT is based on a single study with zero events in the comparator group and the effect estimate is highly uncertain (OR=7.85 [0.38; 163.52]).

**Figure 2.**
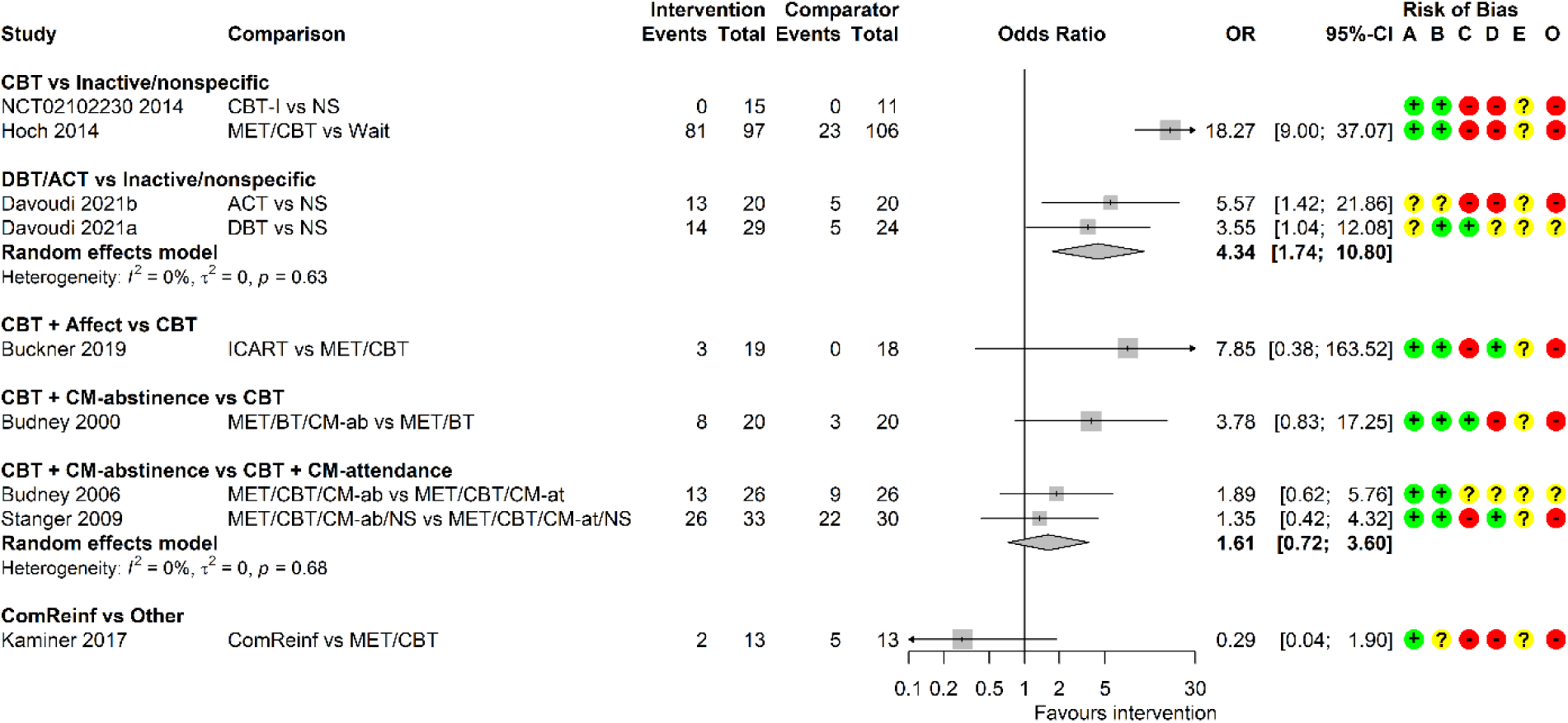
Forest plot for random-effects meta-analyses of point abstinence at end of treatment. ACT, acceptance and commitment therapy; BT, behavioural therapy; CBT, cognitive-behavioural therapy; CBT-I, CBT for insomnia; CI, confidence interval; CM-ab/at, contingency management based on abstinence/attendance; ComReinf, community reinforcement; DBT, dialectical behavioural therapy; ICART, integrated cannabis and anxiety reduction treatment; MET, motivation enhancement therapy; NS, nonspecific comparator; OR, odds ratio; Wait, waitlist. Risk of bias (A) arising from the randomization process, (B) due to deviations from intended interventions, (C) due to missing outcome data, (D) in measurement of the outcome, (E) in selection of the reported result, (O) overall; ‘+’, low risk, ‘?’, some concerns, ‘-‘, high risk of bias.

#### Continuous abstinence

Ten studies measured continuous abstinence (lasting 6-14 weeks) up to the end of treatment.^44,45,47–54^ Most used self-report measures, with only two using consecutive negative urine tests,^44,47^ and one verifying self-reports with urine tests.^50^ Evidence of effectiveness is of low to very low certainty due to concerns over high RoB, imprecision and inconsistency (see Supporting Information 12 for comparison-specific assessments). Meta-analyses included up to four studies per comparison (**Figure 3**). The common τ^2^ was estimated as 0.49 (SE=0.42). CBT may increase continuous abstinence relative to inactive/nonspecific comparators (OR=2.72 [1.20; 6.19]). This analysis is characterized by high heterogeneity (I^2^=82%) that may be explained by comparator type (waitlist or nonspecific). CBT plus CM-abstinence may increase continuous abstinence relative to CBT plus CM-attendance, although CIs are also consistent with a decrease in abstinence (i.e. favouring CBT plus CM-attendance; OR=2.04 [0.75; 5.58]). There is little to no evidence of an effect of CBT plus CM-abstinence relative to CBT (OR=1.81 [0.61; 5.41]). The comparison of community reinforcement versus nonspecific comparator is based on one study^54^ with unclear definition of continuous abstinence and the effect estimate is highly uncertain (OR=47.36 [16.00; 140.21]).

**Figure 3.**
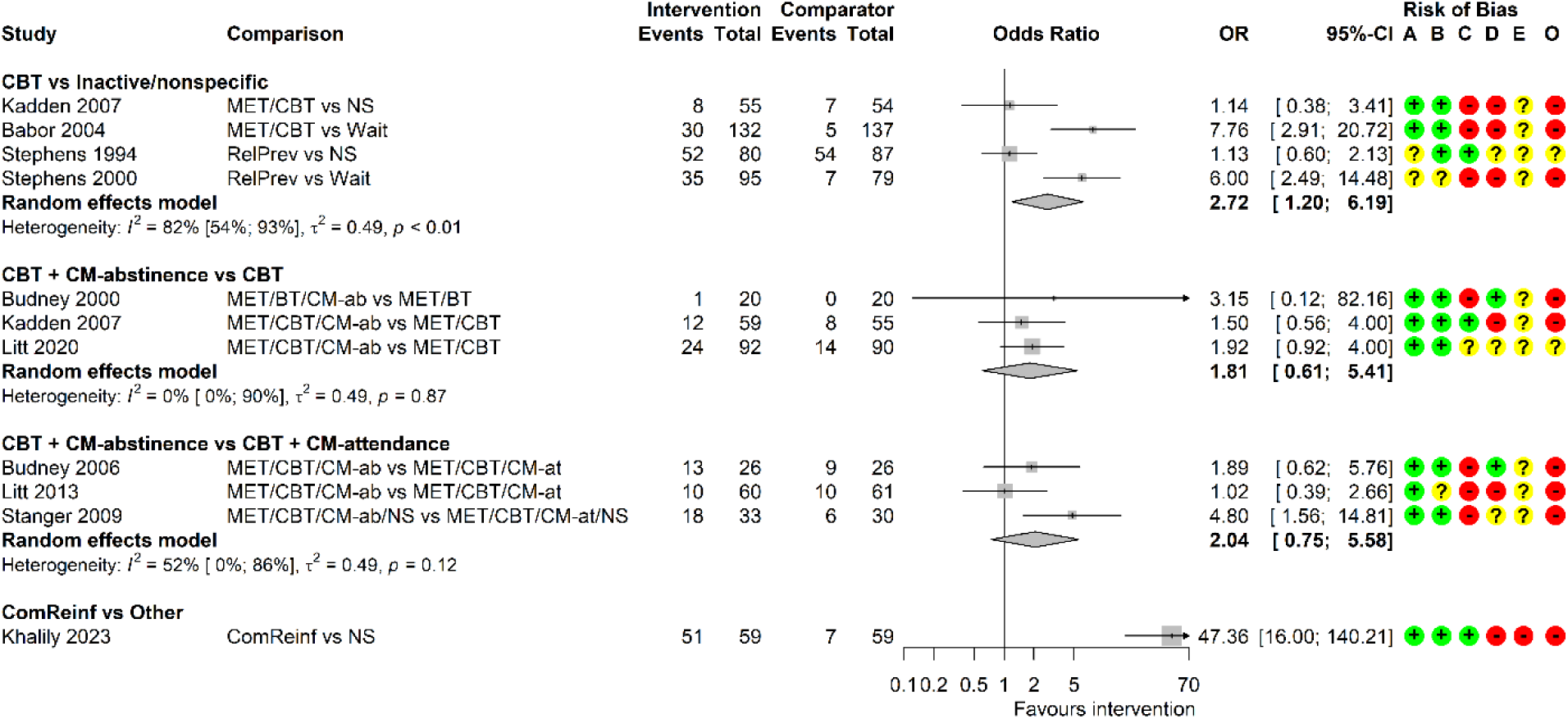
Forest plot for random-effects meta-analyses of continuous abstinence at end of treatment. BT, behavioural therapy; CBT, cognitive-behavioural therapy; CI, confidence interval; CM-ab/at, contingency management based on abstinence/attendance; ComReinf, community reinforcement; MET, motivation enhancement therapy; NS, nonspecific comparator; OR, odds ratio; RelPrev, relapse prevention; Wait, waitlist. Risk of bias (A) arising from the randomization process, (B) due to deviations from intended interventions, (C) due to missing outcome data, (D) in measurement of the outcome, (E) in selection of the reported result, (O) overall; ‘+’, low risk, ‘?’, some concerns, ‘-‘, high risk of bias.

For the related outcome of mean duration of continuous abstinence, there is very low certainty evidence of little to no effect for CBT versus nonspecific comparator, CBT plus CM-abstinence versus CBT, and CBT plus CM-abstinence versus CBT plus CM-attendance (RoMs range 1.24-1.40; Supporting Information 10).

#### Completion of treatment

The number of participants who completed treatment was reported in 16 studies.^39,41–43,45–48,50–52,54–58^ Meta-analyses included a maximum of five studies (**Figure 4**). The common τ^2^ was estimated as 0.00 (SE=0.13). There is low certainty evidence that CBT may be associated with lower completion rates than inactive/nonspecific comparators (OR=0.53 [0.35; 0.82]). We found low certainty evidence for CBT plus CM-abstinence compared with CBT (OR=1.58 [0.85; 2.94]), and for community reinforcement relative to CBT or nonspecific comparator (OR=1.20 [0.49; 2.96]). The certainty of evidence is very low for DBT/ACT versus inactive/nonspecific comparators (OR=1.42 [0.59; 3.43]), CBT-affect versus CBT (OR=1.03 [0.45; 2.32]), and CBT plus CM-abstinence versus CBT plus CM-attendance (OR=1.12 [0.48; 2.62]). We had concerns over indirectness across all comparisons, and RoB and imprecision for most comparisons (Supporting Information 12).

**Figure 4.**
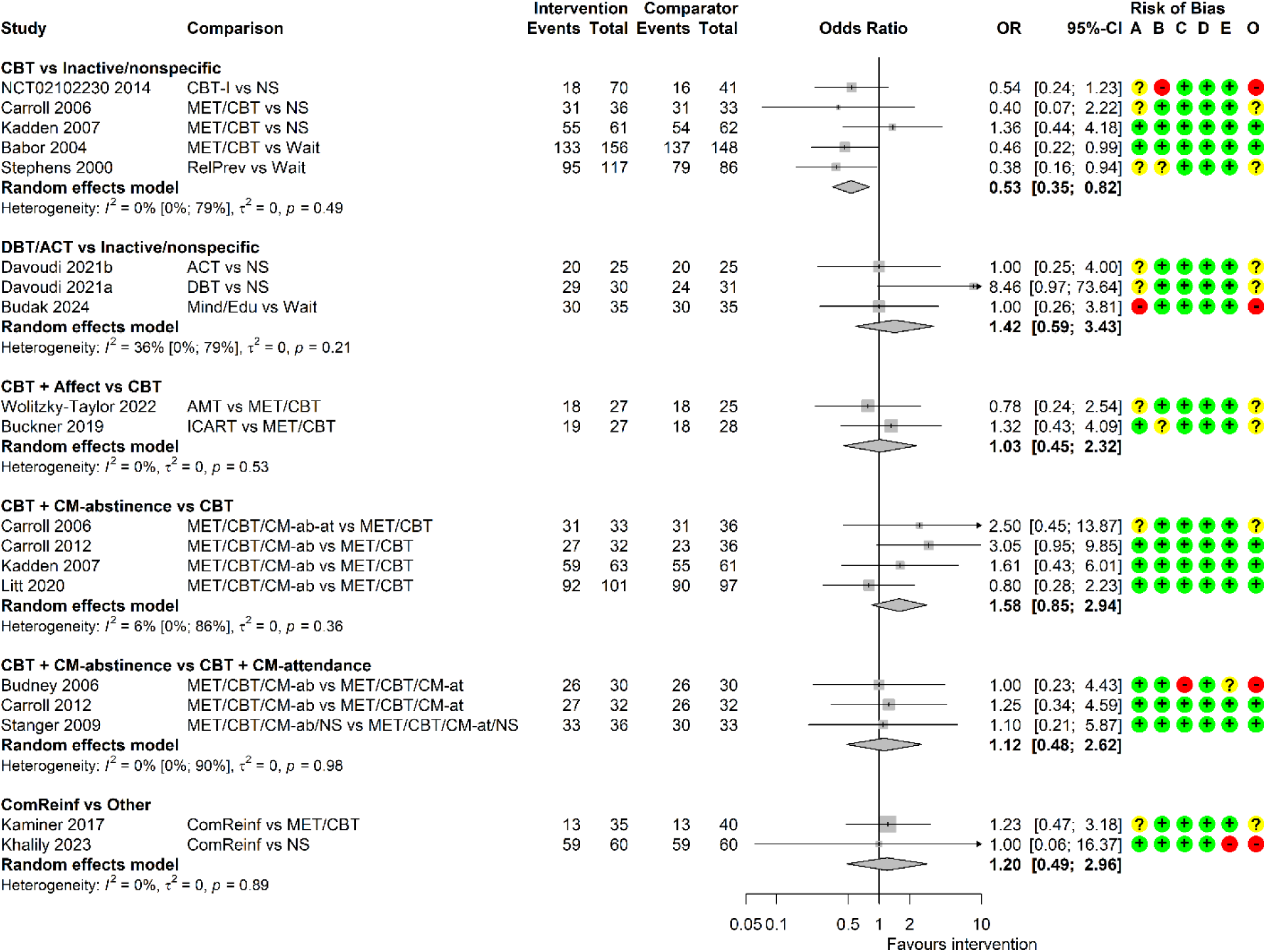
Forest plot for random-effects meta-analyses of completion of treatment. ACT, acceptance and commitment therapy; AMT, affect management therapy; CBT, cognitive-behavioural therapy; CBT-I, CBT for insomnia; CI, confidence interval; CM-ab/at, contingency management based on abstinence/attendance; ComReinf, community reinforcement; DBT, dialectical behavioural therapy; ICART, integrated cannabis and anxiety reduction treatment; MET, motivation enhancement therapy; Mind/Edu, mindfulness psychoeducation; NS, nonspecific comparator; OR, odds ratio; RelPrev, relapse prevention; Wait, waitlist. Risk of bias (A) arising from the randomization process, (B) due to deviations from intended interventions, (C) due to missing outcome data, (D) in measurement of the outcome, (E) in selection of the reported result, (O) overall; ‘+’, low risk, ‘?’, some concerns, ‘-‘, high risk of bias.

#### Frequency of cannabis use

Sixteen studies^39,41,42,44,45,47–53,55,57–59^ reported frequency of use measured over past 7-90 days. Fifteen used self-report, and one used weekly urine tests.^55^ Meta-analyses included up to six studies per comparison (**Figure 5**). The common τ^2^ was estimated as 0.08 (SE=0.05). The evidence is of very low certainty due to high RoB for all comparisons, and due to imprecision, indirectness, and inconsistency among some comparisons (Supporting Information 12). DBT/ACT may have a clinically meaningful effect on reducing frequency of use relative to nonspecific comparators (RoM=0.39, 95% CI [0.25; 0.60]). For other intervention comparisons there is little to no evidence of an effect, i.e., none were estimated to halve or double the frequency of use. This includes CBT versus inactive/nonspecific comparators (RoM=0.63 [0.48; 0.83]), CBT-affect versus CBT (RoM=0.93 [0.56; 1.55]), CBT plus CM-abstinence versus CBT (RoM=0.88 [0.65; 1.19]), CBT plus CM-abstinence versus CBT plus CM-attendance (RoM=0.98 [0.68; 1.40]), and MDFT versus CBT (RoM=0.81 [0.69; 0.95]). The analysis of CBT versus inactive/nonspecific comparators is characterized by high heterogeneity (I^2^=83%) that may be explained by comparator type.

**Figure 5.**
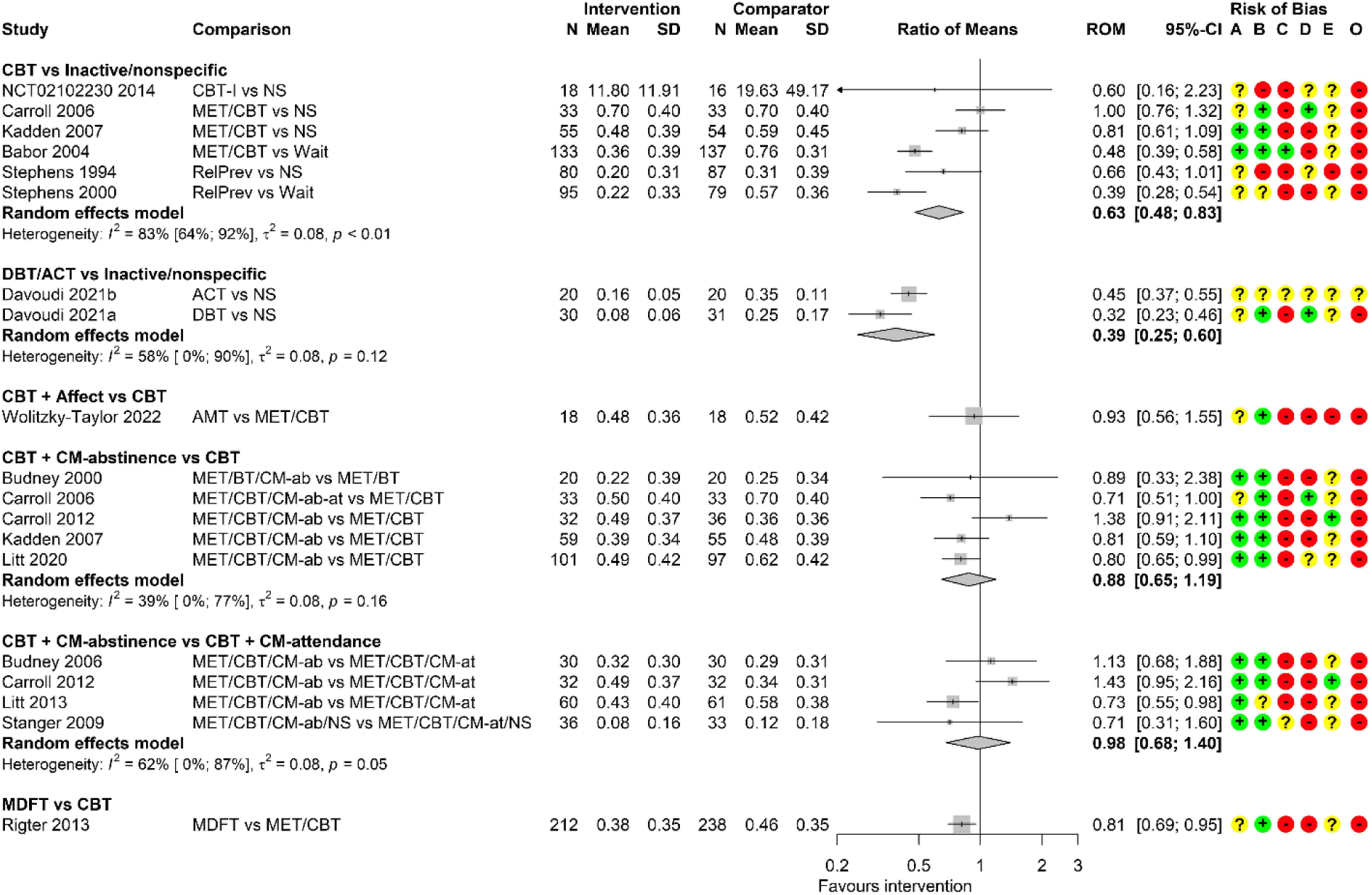
Forest plot for random-effects meta-analyses of frequency of cannabis use at the end treatment. Frequency of use is expressed as proportion of days using for most studies, except for proportion of weeks using in Carroll 2006, and number of uses in NCT02102230. ACT, acceptance and commitment therapy; AMT, affect management therapy; BT, behavioural therapy; CBT, cognitive-behavioural therapy; CBT-I, CBT for insomnia; CI, confidence interval; CM-ab/at, contingency management based on abstinence/attendance; DBT, dialectical behavioural therapy; MDFT, multidimensional family therapy; MET, motivation enhancement therapy; NS, nonspecific comparator; RelPrev, relapse prevention; ROM, ratio of means; Wait, waitlist. Risk of bias (A) arising from the randomization process, (B) due to deviations from intended interventions, (C) due to missing outcome data, (D) in measurement of the outcome, (E) in selection of the reported result, (O) overall; ‘+’, low risk, ‘?’, some concerns, ‘-‘, high risk of bias.

#### Quantity of cannabis use

Two studies reported quantity of cannabis use. This evidence is of very low certainty due to concerns over high RoB, imprecision and, for Buckner 2019^43^, indirectness. Babor 2024^51^ measured self-reported number of joints smoked per typical day of use over past 90 days. The results suggest that CBT may have a clinically meaningful effect on reducing quantity of use when compared with a waitlist control (RoM=0.49 [0.35; 0.69]). Buckner 2019^43^ measured self-reported total number of joints smoked over past 30 days. The evidence indicates that CBT-affect may reduce the quantity of use when compared with CBT (RoM=0.49 [0.17; 1.38]), although CIs are also consistent with an increase in the quantity of use.

#### Craving

A single study measured current cannabis craving, using Marijuana Craving Questionnaire short-form (Davoudi 2021a).^41^ Results indicate there may be little to no evidence of an effect of DBT on reducing craving relative to a nonspecific comparator (RoM=0.95 [0.86; 1.04]; very low certainty evidence due to RoB, imprecision, and indirectness).

#### Adverse events

Two studies reported adverse events.^39,45^ Stanger 2009^45^ compared CBT plus CM-abstinence with CBT plus CM-attendance and NCT02102230 2014^39^ compared CBT with a nonspecific comparator. Certainty of evidence is very low due to concerns over RoB, imprecision, and indirectness. For both studies, we could not estimate intervention effects due to the lack of adverse events in either group.

#### Cost-effectiveness outcomes

We identified two trial-based economic evaluations,^60,61^ both carried out from a healthcare perspective. Full details of these studies and accompanying critical appraisal are presented in Supporting Information 13. Goorden 2016^60^ conducted cost-effectiveness analyses of MDFT compared with CBT for adolescents, based on a single Dutch site of a multicentre trial.^59^ Over a 12-month time horizon, MDFT was associated with higher costs than CBT, better quality of life (EQ-5D-3L), and increased recovery rate (difference in recovery was not statistically significant). The incremental cost-effectiveness ratio (ICER) was €54,308 per quality-adjusted life years (QALYs) gained, and €43,405 per additional recovered patient.

Olmstead 2007^61^ conducted cost-effectiveness analyses of CBT plus CM-abstinence/attendance, CBT alone, CM-abstinence/attendance, and nonspecific comparator (counselling) for young adults referred by the criminal justice system, based on a multi-arm trial^55^ from the United States. Over the initial eight-week treatment period and an additional eight-month time horizon, interventions did not differ significantly in effectiveness, but the costs were the highest for CBT plus CM, followed by CM, CBT, and nonspecific comparator. ICERs for CBT relative to nonspecific comparator were $102 USD per additional week of continuous abstinence (reduced to $34 USD at follow-up) and $159 USD per additional negative urine sample. For CM relative to CBT, ICER was $1104 USD per additional week of continuous abstinence. For CBT plus CM relative to CBT, ICERs were $1333 USD per additional week of continuous abstinence (reduced to $915 USD at follow-up), and $942 USD per additional negative urine sample.

#### Effectiveness outcomes at follow-up

Based on a smaller number of studies, there is some evidence that beneficial effects of CBT and DBT/ACT versus inactive/nonspecific comparators for point abstinence, of CBT plus CM-abstinence versus CBT plus CM-attendance for point and continuous abstinence, and of community reinforcement versus other interventions for continuous abstinence, may be maintained up to six months post-treatment. The relative advantages of the latter two may still be present over six months post-treatment (Supporting Information 10).

#### Sensitivity analyses

Results of the sensitivity analyses using fixed-effect meta-analyses were broadly consistent with random-effects across all effectiveness outcomes. Sensitivity analyses imputing missing observations as abstinent at the end of treatment suggest the effects of some PSIs on point abstinence may be reduced, but remain similar for continuous abstinence. Imputing missing observations as non-abstinent had little impact on the results (Supporting Information 11).

## DISCUSSION

This review aimed to evaluate the effectiveness, safety, and cost-effectiveness of PSIs for CUD in people aged ≥16 years. We included 22 RCTs (3304 participants). We judged the certainty of the evidence to be low to very low, due to concerns of high RoB and imprecision of the estimated treatment effects. Various effectiveness outcomes were reported across the studies and only two reported safety outcomes.

CBT-based interventions were the most commonly evaluated PSI, followed by CM and DBT/ACT. At the end of treatment, we found CBT led to a clinically meaningful benefit for point and continuous abstinence but was associated with lower treatment completion. CBT-affect and CBT plus CM-abstinence compared with CBT may have clinically meaningful effects on point abstinence, but there was no evidence that CBT plus CM-abstinence improved continuous abstinence. CBT plus CM-abstinence may improve continuous abstinence compared with CBT plus CM-attendance. The impact of CBT or CBT plus CM on adverse events (safety) was unclear.

We found DBT/ACT compared with inactive/nonspecific comparators may improve point abstinence at the end of treatment. Community reinforcement was less likely to improve point abstinence when compared with CBT.^46^ Although we found a clinically meaningful effect of community reinforcement on continuous abstinence relative to a nonspecific comparator, the definition of continuous abstinence used was unclear and this finding is highly uncertain.^54^ There was no evidence that PSIs other than CBT affected completion of treatment.

For secondary outcomes at the end of treatment, we found some evidence that CBT-based interventions may reduce the quantity of cannabis used. Except for DBT/ACT, other PSIs were unlikely to reduce frequency of use. There was little evidence that CBT-based interventions could increase the duration of continuous abstinence when compared with inactive/nonspecific or active comparators.

Two trial-based economic evaluations reported higher costs for CBT and CM relative to a nonspecific comparator,^61^ and for MDFT compared with CBT.^60^ The trialists reported little difference in effectiveness, but quality of life (EQ-5D-3L) was improved for MDFT. No studies reported intensity of withdrawal, engagement in further treatment, or dropout due to adverse events.

While our findings are consistent with previous reviews of PSIs for point abstinence^13,14^, they are not consistent for frequency of use, where only DBT/ACT showed potential benefit in the present review. Unlike previous meta-analyses, we did not aggregate PSIs into a single intervention category but analyzed them based on shared theoretical underpinning or techniques used.^62^ Previous reviews either did not assess RoB^14^ or used earlier versions of the Cochrane RoB tool.^13^ We used the RoB2 tool, which focuses on bias at the outcome- and not study-level. Earlier meta-analyses also employed random-effects models but used a DerSimonian-Laird between-study variance estimator, which may be negatively biased when study sample sizes are small and heterogeneity is large. We used a more robust REML estimator.^38,63^ These methodological differences may also explain why we rated the certainty of the evidence base as low to very low, in contrast to a previous assessment of low to moderate certainty.^13^

### Strengths and limitations

Our review followed a rigorous methodology. The protocol was prospectively registered and the database searches were comprehensive and recently conducted (12-Jun-2024). However, the number of studies included per meta-analysis was small (up to six), and some relative intervention effects were based on single studies. This means that our pre-planned subgroup analyses were not possible. We also excluded some publication types, such as conference abstracts, or trial registration reports lacking outcome data.

We implemented clearly defined eligibility criteria. However, the included studies used a variety of participant inclusion criteria regarding CUD, which may have contributed to the heterogeneity of the current findings. This variability may reflect changes in the diagnostic criteria over time. However, even within the same diagnostic categories, the severity of the disorder may vary and this was not consistently reported across all studies (**Table 3**).

To combine studies in a meta-analysis, similar interventions and comparators were grouped. However, these groupings could impact interpretation of estimated effects. For instance, the study-level effects were greater when CBT was compared with a waitlist control, and smaller when compared with a nonspecific comparator (Figures 2, 3, 5). This ‘waitlist’ effect is well-known in behavioural intervention trials.^64,65^ Unfortunately, our review lacked sufficient studies for appropriately powered subgroup analyses by comparator type.

Although there has been some work on outcome toolkits,^66^ there is no agreed core outcome set for CUD and we found considerable variability in outcome definitions and measures across all studies. Additionally, prospectively registered protocols were not available for 15 of 22 studies. As such, it is unclear whether our selected outcomes were not measured, or were measured but not reported. Abstinence was measured using self-report or urine tests, with limited details on thresholds for detecting presence of cannabinoids, and which measure had informed the outcome assessment when both were collected. The definition of ‘continuous’ abstinence also varied across studies (6-14 weeks). Using different measures and timeframes within the abstinence outcomes likely contributed to moderate between-study heterogeneity, which in turn reduces the certainty of findings.

Only two studies assessed safety outcome and both reported that no adverse events had occurred. Neither provided a definition of ‘adverse event’ or details on measurement. Reasons for participant dropout were poorly reported across all studies. In combination with substantial missing data, this was a common source of high RoB in the results. Cannabis craving was reported in one study only,^41^ and none reported on the intensity of withdrawal. This was unexpected considering the nature of the PSIs, some of which included training in affect management and dealing with withdrawal. For individuals with CUD, withdrawal symptoms can occur within the first week of ceasing cannabis use.^3^ Except for the studies of CM-abstinence,^44,45,47–50,55,58^ it was not clear whether or when participants were expected to stop using cannabis. Therapeutic goals may extend beyond abstinence and include reduction in cannabis use or improvement in functioning, where withdrawal symptoms may be less likely to occur.

In addition to the absence of a core outcome set, there is no established consensus for what constitutes a clinically meaningful change in outcomes. For this review, we considered a 10% increase in abstinence and completion of treatment and halving the level of cannabis use in the intervention group would represent clinically meaningful changes. However, these thresholds may be considered high, especially from a harm-reduction perspective.^67^ Abstinence is difficult to achieve^68^ and a smaller reduction in cannabis use may be meaningful for people with CUD. It is possible that some PSIs would have been considered effective if lower thresholds had been used.

### Implications for research and practice

Several PSIs demonstrated clinically meaningful effects on abstinence and level of cannabis use and, in the absence of alternative treatments, it would be reasonable to suggest they are offered for CUD. However, due to the low certainty of evidence and small number of studies this recommendation for clinical practice is tentative.

To improve the robustness of the evidence base, and inform policy and practice, additional high-quality RCTs are needed. This is the same conclusion reached by Gates et al in their 2016 Cochrane review.^13^ Studies should be prospectively registered with published trial protocols to minimize bias from selection of the reported results. Studies should be adequately powered, and ensure the assessors are blinded, at least to the alternative intervention in case of self-reported outcomes. To reduce the impact of missing outcome data, trials should incorporate strategies to retain and follow-up participants and clearly report information concerning those who have withdrawn. Future research on core outcome sets and standardized measurement of outcomes should also involve people with CUD.

Many trials in this review were conducted over a decade ago and may not generalise to contemporary cannabis use. In recent years, there has been a surge in CUD incidence among adolescents and young adults.^6^ While we had insufficient data to explore whether age may moderate the effectiveness of PSIs, the specific needs of young people should be considered within treatment services. The potency of cannabis has also increased over time,^69,70^ and is associated with risk of CUD and more severe dependence,^71,72^ and may reduce treatment effectiveness. Future studies should also ensure participants from more diverse backgrounds are recruited, as the current evidence base predominantly includes white male participants. Only three studies included people with affective problems,^42,43,57^ and four others reported mental health co-morbidities in some participants,^44,45,55,58^ whereas most excluded people with severe mental health problems (e.g. suicide risk, psychosis). Mental health disorders commonly co-occur with CUD,^73,74^ as does nicotine dependence. Evaluation of integrated treatments may be valuable to improve outcomes for people with CUD.^75,76^

Such research would underpin the development of cannabis-specific, evidence-based, practice guidelines.

### Conclusions

This review found that CBT, DBT/ACT and community reinforcement may be superior to inactive/nonspecific interventions for the treatment of CUD. Effectiveness and cost-effectiveness of other therapies and their combinations were less clear. The evidence for the effectiveness and safety of PSIs is of low to very low certainty. Methodologically robust trials conducted in representative samples of people with CUD are needed to inform more certain recommendations for policy and clinical practice.

## Supporting information

SuppInfo11

SuppInfo12

SuppInfo13

SuppInfo01

SuppInfo02

SuppInfo03

SuppInfo04

SuppInfo05

SuppInfo06

SuppInfo07

SuppInfo08

SuppInfo09

SuppInfo10

## ADDITIONAL INFORMATION

### CRediT statement

Conceptualization: FS, SS, JS, JH, DC; Data Curation: MH, TP, KW, FS, SD, CP; Formal analysis: MH; Funding acquisition: JS, JH, DC; Investigation: MH, TP, KW, FS, DC; Methodology: MH, LAH, TPF, SS, SD, JS, JH, DC; Project administration: MH, DC; Resources: MH; Software: MH; Supervision: LAH, TPF, SS, JH, DC; Validation: MH, TP, KW; Visualization: MH, TP, KW; Writing – Original draft: MH, KW, DC; Writing – reviewing and editing: MH, TP, KW, FS, LAH, TPF, SS, JS, JH, DC.

## Data availability statement

The data to support the findings of this review are available in Table 3, Figures 2-5, and Supporting Information 8-13.

## SUPPORTING INFORMATION LEGENDS

Supporting Information 1. Intervention groupings (.docx)

Supporting Information 2. Operationalization of outcomes and hierarchy of preference for outcome measures and timepoints (.docx)

Supporting Information 3. Search strategies (.docx)

Supporting Information 4. Data processing (.docx)

Supporting Information 5. Additional comparisons (.docx)

Supporting Information 6. Certainty of evidence (GRADE) criteria (.docx)

Supporting Information 7. Excluded studies (.docx)

Supporting Information 8. PROGRESS-Plus characteristics (.docx)

Supporting Information 9. Risk of bias assessment (.docx)

Supporting Information 10. Results synthesis for outcomes assessed at medium and long follow-up (.docx)

Supporting Information 11. Results of sensitivity analyses (.docx)

Supporting Information 12. Summary of findings tables (.docx)

Supporting Information 13. Economic evaluation studies (.docx)

