## Supplementary material for "Effectiveness and safety of psychosocial interventions for the treatment of cannabis use disorder: a systematic review and meta-analysis": SuppInfo11

### SUPPORTING INFORMATION 11. RESULTS OF SENSITIVITY ANALYSES

Numbering of tables is specific to this Supporting Information document. References relating to this Supporting Information are included at the end of this document.

#### Random effects versus fixed effect meta-analysis

**Table 1.** Relative effect estimates from random-effects and fixed-effect meta-analyses

| Outcome | Comparison (number of studies) | Random effects | Fixed effect |
|---|---|---|---|
| **Dichotomous outcomes (OR [95% CI])** | | | |
| Point abstinence at end of treatment* | CBT vs Inactive/nonspecific (1) | 18.27 [9.00; 37.07] | 18.27 [9.00; 37.07] |
| | DBT/ACT vs Inactive/nonspecific (2) | 4.34 [1.74; 10.80] | 4.34 [1.74; 10.80] |
| | CBT + Affect vs CBT (1) | 7.85 [0.38; 163.52] | 7.85 [0.38; 163.52] |
| | CBT + CM-abstinence vs CBT (1) | 3.78 [0.83; 17.25] | 3.78 [0.83; 17.25] |
| | CBT + CM-abstinence vs CBT + CM-attendance (2) | 1.61 [0.72; 3.60] | 1.61 [0.72; 3.60] |
| | ComReinf vs Other (1) | 0.29 [0.04; 1.90] | 0.29 [0.04; 1.90] |
| Point abstinence at medium follow-up* | CBT vs Inactive/nonspecific (1) | 2.29 [0.78; 6.69] | 2.29 [0.78; 6.69] |
| | DBT/ACT vs Inactive/nonspecific (2) | 5.19 [1.83; 14.67] | 5.19 [1.83; 14.67] |
| | CBT + CM-abstinence vs CBT (1) | 0.63 [0.21; 1.88] | 0.63 [0.21; 1.88] |
| | CBT + CM-abstinence vs CBT + CM-attendance (2) | 3.93 [1.57; 9.82] | 3.93 [1.57; 9.82] |
| Point abstinence at long follow-up* | CBT + CM-abstinence vs CBT + CM-attendance (2) | 2.55 [1.12; 5.81] | 2.55 [1.12; 5.81] |
| Continuous abstinence at end of treatment | CBT vs Inactive/nonspecific (4) | 2.72 [1.20; 6.19] | 2.62 [1.77; 3.88] |
| | CBT + CM-abstinence vs CBT (3) | 1.81 [0.61; 5.41] | 1.79 [1.01; 3.20] |
| | CBT + CM-abstinence vs CBT + CM-attendance (3) | 2.04 [0.75; 5.58] | 1.96 [1.08; 3.55] |
| | ComReinf vs Other (1) | 47.36 [8.19; 273.74] | 47.36 [16.00; 140.21] |
| Continuous abstinence at medium follow-up | CBT vs Inactive/nonspecific (4) | 1.87 [0.80; 4.35] | 1.65 [1.02; 2.66] |
| | CBT + CM-abstinence vs CBT (3) | 0.85 [0.36; 2.02] | 0.97 [0.61; 1.54] |
| | CBT + CM-abstinence vs CBT + CM-attendance (2) | 2.85 [0.87; 9.30] | 2.80 [1.30; 6.03] |
| | ComReinf vs Other (1) | 30.86 [6.06; 157.03] | 30.86 [10.27; 92.69] |
| Continuous abstinence at long follow-up* | CBT vs Inactive/nonspecific (2) | 1.28 [0.67; 2.45] | 1.28 [0.67; 2.45] |
| | CBT + CM-abstinence vs CBT (2) | 1.12 [0.66; 1.90] | 1.12 [0.66; 1.90] |
| | CBT + CM-abstinence vs CBT + CM-attendance (2) | 1.69 [0.77; 3.70] | 1.69 [0.77; 3.70] |
| | ComReinf vs Other (1) | 28.17 [9.72; 81.65] | 28.17 [9.72; 81.65] |
| Completion of treatment* | CBT vs Inactive/nonspecific (5) | 0.53 [0.35; 0.82] | 0.53 [0.35; 0.82] |
| | DBT/ACT vs Inactive/nonspecific (3) | 1.42 [0.59; 3.43] | 1.42 [0.59; 3.43] |
| | CBT + Affect vs CBT (2) | 1.03 [0.45; 2.32] | 1.03 [0.45; 2.32] |
| | CBT + CM-abstinence vs CBT (4) | 1.58 [0.85; 2.94] | 1.58 [0.85; 2.94] |
| | CBT + CM-abstinence vs CBT + CM-attendance (3) | 1.12 [0.48; 2.62] | 1.12 [0.48; 2.62] |
| | ComReinf vs Other (2) | 1.20 [0.49; 2.96] | 1.20 [0.49; 2.96] |
| **Continuous outcomes (RoM [95% CI])** | | | |
| Duration of continuous abstinence at end of treatment | CBT vs Inactive/nonspecific (2) | 1.24 [0.70; 2.19] | 1.21 [0.76; 1.91] |
| | CBT + CM-abstinence vs CBT (5) | 1.40 [1.02; 1.91] | 1.35 [1.06; 1.72] |
| | CBT + CM-abstinence vs CBT + CM-attendance (4) | 1.31 [0.97; 1.77] | 1.32 [1.07; 1.63] |
| Duration of continuous abstinence at long follow-up* | CBT vs Inactive/nonspecific (1) | 1.08 [0.71; 1.65] | 1.08 [0.71; 1.65] |
| | CBT + CM-abstinence vs CBT (1) | 1.17 [0.77; 1.79] | 1.17 [0.77; 1.79] |
| Frequency of cannabis use at end of treatment | CBT vs Inactive/nonspecific (6) | 0.63 [0.48; 0.83] | 0.61 [0.54; 0.69] |
| | DBT/ACT vs Inactive/nonspecific (2) | 0.39 [0.25; 0.60] | 0.41 [0.35; 0.49] |
| | CBT + Affect vs CBT (1) | 0.93 [0.44; 1.96] | 0.93 [0.56; 1.55] |

| Outcome | Comparison (number of studies) | Random effects | Fixed effect |
|---|---|---|---|
| | CBT + CM-abstinence vs CBT (5) | 0.88 [0.65; 1.19] | 0.84 [0.73; 0.97] |
| | CBT + CM-abstinence vs CBT + CM-attendance (4) | 0.98 [0.68; 1.40] | 0.93 [0.76; 1.15] |
| | MDFT vs CBT (1) | 0.81 [0.46; 1.43] | 0.81 [0.69; 0.95] |
| Frequency of cannabis use at medium follow-up | CBT vs Inactive/nonspecific (5) | 0.94 [0.73; 1.22] | 0.95 [0.81; 1.11] |
| | DBT/ACT vs Inactive/nonspecific (2) | 0.54 [0.38; 0.75] | 0.55 [0.47; 0.64] |
| | CBT + Affect vs CBT (1) | 1.28 [0.55; 2.94] | 1.28 [0.62; 2.63] |
| | CBT + CM-abstinence vs CBT (4) | 1.03 [0.78; 1.35] | 0.98 [0.83; 1.15] |
| | CBT + CM-abstinence vs CBT + CM-attendance (4) | 0.87 [0.64; 1.19] | 0.86 [0.70; 1.06] |
| | MDFT vs CBT (1) | 0.80 [0.51; 1.26] | 0.80 [0.68; 0.95] |
| Frequency of cannabis use at long follow-up | CBT vs Inactive/nonspecific (2) | 0.98 [0.81; 1.19] | 0.98 [0.81; 1.19] |
| | CBT + CM-abstinence vs CBT (3) | 0.97 [0.81; 1.17] | 0.97 [0.81; 1.17] |
| | CBT + CM-abstinence vs CBT + CM-attendance (3) | 0.80 [0.65; 0.98] | 0.80 [0.65; 0.98] |
| Quantity of cannabis use at end of treatment* | CBT vs Inactive/nonspecific (1) | 0.49 [0.35; 0.69] | 0.49 [0.35; 0.69] |
| | CBT + Affect vs CBT (1) | 0.49 [0.17; 1.38] | 0.49 [0.17; 1.38] |
| Quantity of cannabis use at medium follow-up* | CBT vs Inactive/nonspecific (1) | 0.61 [0.48; 0.76] | 0.61 [0.48; 0.76] |
| Cravings at end of treatment* | DBT/ACT vs Inactive/nonspecific (1) | 0.95 [0.86; 1.04] | 0.95 [0.86; 1.04] |
| Cravings at medium follow-up* | DBT/ACT vs Inactive/nonspecific (1) | 0.93 [0.84; 1.03] | 0.93 [0.84; 1.03] |

*Note that where between-study variance ($\tau^2$) was estimated as 0 for random-effects meta-analyses, fixed-effect meta-analyses provide equivalent effect estimates. Study-level estimates where only a single study contributed to a specific comparison are also included for completeness.

CBT, cognitive-behavioural therapy; CI, confidence interval; CM, contingency management; ComReinf, community reinforcement; DBT/ACT, dialectical behavioural therapy/acceptance and commitment therapy; MDFT, multidimensional family therapy; OR, odds ratio; RoM, ratio of means.

### Imputation of missing data for abstinence outcomes

The primary analyses for point and continuous abstinence outcomes used the number of participants for whom outcome data were assessed. An alternative approach would be to use the number of participants randomized (at baseline) and impute missing outcome data. We addressed the sensitivity of the primary results to this decision in the following analyses.

Sensitivity analyses were conducted using random-effects meta-analyses of abstinence outcomes based on the number of participants randomized to each study under (a) a pessimistic scenario, where all missing cases were imputed as non-abstinent, and (b) an optimistic scenario, where all missing cases were imputed as abstinent. Table 2 presents the effect estimates under each scenario alongside the estimates from the primary analysis.

**Table 2.** Relative effect estimates from sensitivity analyses of point and continuous abstinence outcomes

| Outcome | Comparison (number of studies) | Primary analysis (OR [95% CI]) | Sensitivity analysis – pessimistic (OR [95% CI]) | Sensitivity analysis – optimistic (OR [95% CI]) |
|---|---|---|---|---|
| Point abstinence at end of treatment | CBT vs Inactive/nonspecific (1) | 18.27 [9.00; 37.07] | 5.54 [3.18; 9.64] | 4.69 [1.06; 20.77] (N = 2); *14.68 [7.82; 27.56] |
| | DBT/ACT vs Inactive/nonspecific (2) | 4.34 [1.74; 10.80] | 4.45 [1.87; 10.57] | 2.43 [0.50; 11.90]; *2.31 [1.07; 5.00] |
| | CBT + Affect vs CBT (1) | 7.85 [0.38; 163.52] | 8.14 [0.40; 165.53] | 1.24 [0.13; 11.63]; *1.24 [0.42; 3.68] |
| | CBT + CM-abstinence vs CBT (1) | 3.78 [0.83; 17.25] | 3.78 [0.83; 17.25] | 3.78 [0.32; 45.02]; *3.78 [0.83; 17.25] |
| | CBT + CM-abstinence vs CBT + CM-attendance (2) | 1.61 [0.72; 3.60] | 1.51 [0.72; 3.17] | 1.51 [0.31; 7.35]; *1.53 [0.71; 3.28] |
| | ComReinf vs Other (1) | 0.29 [0.04; 1.90] | 0.42 [0.08; 2.34] | 0.55 [0.06; 5.04]; *0.55 [0.19; 1.56] |
| Point abstinence at medium follow-up | CBT vs Inactive/nonspecific (1) | 2.29 [0.78; 6.69] | 1.36 [0.52; 3.51] | 2.11 [0.95; 4.66] (N = 2); *2.82 [0.70; 11.32] |
| | DBT/ACT vs Inactive/nonspecific (2) | 5.19 [1.83; 14.67] | 5.99 [2.20; 16.35] | 1.73 [0.72; 4.15]; *1.77 [0.64; 4.89] |
| | CBT + CM-abstinence vs CBT (1) | 0.63 [0.21; 1.88] | 0.83 [0.32; 2.15] | 0.58 [0.18; 1.92]; *0.58 [0.14; 2.37] |
| | CBT + CM-abstinence vs CBT + CM-attendance (2) | 3.93 [1.57; 9.82] | 2.29 [1.06; 4.94] | 3.17 [1.30; 7.68]; *3.18 [1.14; 8.90] |
| Point abstinence at long follow-up | CBT + CM-abstinence vs CBT + CM-attendance (2) | 2.55 [1.12; 5.81] | 1.93 [0.93; 4.02] | 2.40 [1.15; 4.99] |
| Continuous abstinence at end of treatment | CBT vs Inactive/nonspecific (4) | 2.72 [1.20; 6.19] | 2.34 [1.05; 5.24] | 2.29 [1.18; 4.45] |
| | CBT + CM-abstinence vs CBT (3) | 1.81 [0.61; 5.41] | 1.81 [0.61; 5.33] | 1.52 [0.61; 3.82] |
| | CBT + CM-abstinence vs CBT + CM-attendance (3) | 2.04 [0.75; 5.58] | 1.93 [0.72; 5.17] | 1.87 [0.81; 4.34] |
| | ComReinf vs Other (1) | 47.36 [8.19; 273.74] | 42.90 [7.65; 240.49] | 42.25 [8.96; 199.12] |
| Continuous abstinence at medium follow-up | CBT vs Inactive/nonspecific (4) | 1.87 [0.80; 4.35] | 1.38 [0.74; 2.60] | 1.56 [0.93; 2.61] |
| | CBT + CM-abstinence vs CBT (3) | 0.85 [0.36; 2.02] | 0.95 [0.51; 1.77] | 0.89 [0.49; 1.63] |
| | CBT + CM-abstinence vs CBT + CM-attendance (2) | 2.85 [0.87; 9.30] | 2.52 [1.00; 6.34] | 2.07 [0.94; 4.56] |
| | ComReinf vs Other (1) | 30.86 [6.06; 157.03] | 29.57 [8.47; 103.28] | 22.00 [6.07; 79.68] |

| Outcome | Comparison (number of studies) | Primary analysis (OR [95% CI]) | Sensitivity analysis – pessimistic (OR [95% CI]) | Sensitivity analysis – optimistic (OR [95% CI]) |
|---|---|---|---|---|
| Continuous abstinence at long follow-up | CBT vs Inactive/nonspecific (2) | 1.28 [0.67; 2.45] | 1.18 [0.63; 2.22] | 1.07 [0.67; 1.71] |
| | CBT + CM-abstinence vs CBT (2) | 1.12 [0.66; 1.90] | 1.12 [0.68; 1.87] | 1.06 [0.68; 1.65] |
| | CBT + CM-abstinence vs CBT + CM-attendance (2) | 1.69 [0.77; 3.70] | 1.53 [0.71; 3.29] | 1.65 [0.92; 2.95] |
| | ComReinf vs Other (1) | 28.17 [9.72; 81.65] | 26.00 [9.79; 69.06] | 16.71 [6.17; 45.27] |

*Estimates after excluding NCT02102230 from estimation of common $\tau^2$ and CBT vs Inactive/nonspecific comparison.

CBT, cognitive-behavioural therapy; CI, confidence interval; CM, contingency management; ComReinf, community reinforcement; DBT/ACT, dialectical behavioural therapy/acceptance and commitment therapy; N, number of studies; OR, odds ratio.

For point abstinence, results of the sensitivity analysis under a pessimistic scenario were consistent with the primary meta-analysis, except for the reduced effect of cognitive-behavioural therapy (CBT) versus inactive/nonspecific comparator. However, in an optimistic scenario, effect estimates for CBT, dialectical behavioural/acceptance and commitment therapies (DBT/ACT), and CBT with affect management (CBT-affect) were closer to the null. Note that NCT02102230 trial[1] (which did not contribute to the primary meta-analysis due to zero events in either arm) was terminated early, and optimistic imputation changed the number of events/total observations from 0/26 to 85/111, and increased the between-study variance ($\tau^2$) from 0.00 (standard error, SE=0.38) to 1.00 (SE=1.02). Due to the common heterogeneity parameter across comparisons, this also impacted the DBT/ACT and CBT-affect comparisons. As such, we also report a post-hoc analysis excluding NCT02102230[1] (see additional estimates in Table 2), which brought the $\tau^2$ back to 0.00 (SE=0.31), and note the discrepant results. Specifically, the effect of CBT was reduced to a lesser extent, and the effects of DBT/ACT and CBT-affect to a similar extent, but without excessive widening of their 95% confidence intervals (CIs).

Results at medium follow-up were consistent with the primary analysis under the optimistic scenario, however, under a pessimistic scenario, effect estimates for CBT versus inactive/nonspecific comparator, and CBT plus abstinence-contingency management (CM) versus CBT plus CM-attendance were closer to the null. This was also the case for the latter comparison at long follow-up. While at medium follow-up, the optimistic imputation actually decreased $\tau^2$ (from 0.49 to 0.09), for completeness, we also report a post-hoc analysis excluding NCT02102230[1] for that timepoint ($\tau^2$=0.23; see additional estimates in Table 2).

For continuous abstinence, results of sensitivity analyses under either scenario were broadly consistent with the primary results, except for reduced effect of community reinforcement versus nonspecific comparator at medium and long follow-up under the optimistic scenario.
