## Supplementary material for "Effectiveness and safety of psychosocial interventions for the treatment of cannabis use disorder: a systematic review and meta-analysis": SuppInfo12

### SUPPORTING INFORMATION 12. SUMMARY OF FINDINGS TABLES

To aid the interpretation of results, in the summary of findings tables we additionally re-expressed odds ratios as risk ratios, as per review protocol. Numbering of tables is specific to this Supporting Information document. References relating to this Supporting Information are included at the end of this document.

**Table 1.** CBT versus inactive/nonspecific comparator for cannabis use disorder

| Certainty assessment | | | | | | | Number of participants | | Effect | | Certainty |
|---|---|---|---|---|---|---|---|---|---|---|---|
| Studies | Study design | Risk of bias | Inconsistency | Indirectness | Imprecision | Other considerations | CBT | Inactive/non-specific comparator | Relative [95% CI] | Absolute [95% CI] | |
| **Point abstinence at end of treatment** | | | | | | | | | | | |
| Hoch 2014,[1] NCT02102230 2014[2] | randomized trials | very serious[a] | not serious | not serious | serious[b] | none | 81/97 (83.5%) | 23/117 (19.7%) | **OR=18.27** [9.00; 37.07] **RR=4.10** [3.46; 4.51] | **621 more per 1,000** [from 491 more to 704 more] | ⊕◯◯◯ Very low[a,b] |
| **Continuous abstinence at end of treatment** | | | | | | | | | | | |
| Kadden 2007,[3] Babor 2004,[4] Stephens 1994,[5] Stephens 2000[6] | randomized trials | very serious[c] | not serious[d] | not serious | serious[e] | none | 115/362 (31.8%) | 73/357 (20.4%) | **OR=2.75** [0.99; 7.62] **RR=2.02** [1.15; 3.04] | **210 more per 1,000** [from 2 fewer to 458 more] | ⊕◯◯◯ Very low[c,d,e] |
| **Completion of treatment** | | | | | | | | | | | |
| Carroll 2006,[7] Kadden 2007,[3] Babor 2004,[4] Stephens 2000,[6] NCT02102230 2014[2] | randomized trials | serious[f] | not serious | serious[g] | not serious[h] | none | 332/440 (75.5%) | 317/370 (85.7%) | **OR=0.53** [0.35; 0.82] **RR 0.89** [0.79; 0.97] | **97 fewer per 1,000** [from 180 fewer to 26 fewer] | ⊕⊕◯◯ Low[f,g,h] |
| **Duration of continuous abstinence at end of treatment** | | | | | | | | | | | |
| Carroll 2006,[7] Kadden 2007[3] | randomized trials | very serious[a] | not serious | serious[i] | not serious[j] | none | 94 participants | 95 participants Assumed duration of abstinence 7 days | **RoM=1.21** [0.76; 1.91] | **1.47 days longer** [from 1.68 days fewer to 6.37 days more] | ⊕◯◯◯ Very low[a,i,j] |

| Certainty assessment | | | | | | | Number of participants | | Effect | | Certainty |
|---|---|---|---|---|---|---|---|---|---|---|---|
| **Studies** | **Study design** | **Risk of bias** | **Inconsistency** | **Indirectness** | **Imprecision** | **Other considerations** | **CBT** | **Inactive/non-specific comparator** | **Relative [95% CI]** | **Absolute [95% CI]** | |
| **Frequency of cannabis use at end of treatment** | | | | | | | | | | | |
| Carroll 2006,[7] Kadden 2007,[3] Babor 2004,[4] Stephens 1994,[5] Stephens 2000,[6] NCT02102230 2014[2] | randomized trials | very serious[k] | very serious[l] | not serious[m] | serious[n] | none | 414 participants | 406 participants Assumed proportion of days of use = 0.5, i.e. 3.5 days out of every 7 days | **RoM=0.63** [0.46; 0.8]) | **Proportion of days using lower by 0.185** [from 0.27 lower to 0.065 lower], i.e. a reduction in use to 2.2 days out of every 7 [from 1.61 days to 3.04 days] | ⊕◯◯◯ Very low[k,l,m,n] |
| **Quantity of cannabis use at end of treatment** | | | | | | | | | | | |
| Babor 2004[4] | randomized trials | very serious[o] | not serious | not serious | serious[p] | none | 133 participants | 137 participants Assumed quantity of use 2 joints per day | **RoM=0.49** [0.35; 0.69] | **Quantity of use reduced by 1.02 joints per day** [from 1.3 fewer to 0.62 fewer] | ⊕◯◯◯ Very low[o,p] |
| **Adverse events** | | | | | | | | | | | |
| NCT02102230 2014[2] | randomized trials | very serious[o] | not serious | serious[q] | very serious[r] | none | 0/36 (0%) | 0/24 (0%) | Not estimable | No adverse events occurred in either group. | ⊕◯◯◯ Very low[o,q,r] |

**CBT,** cognitive-behavioural therapy; **CI,** confidence interval; **OR,** odds ratio; **RoM**, ratio of means; **RR,** risk ratio.

**Explanations**

a. Both studies considered to be at high risk of bias.

b. Total sample size fails to meet optimal information size (estimated at 586 participants, based on control group risk of 20%, with power of 0.8 to detect an increase in abstinence of 10%).

c. Three studies rated at high risk of bias, one rated some concerns.

d. $I^2$=82%. Two studies indicate beneficial effects, two indicate very little effect. However, this variation is also reflected in the wide confidence intervals, therefore the certainty of evidence was not reduced twice for this concern.

e. Sample size is sufficient to meet OIS (estimated at 586 participants, based on control group risk of 20%, with power of 0.8 to detect an increase in abstinence of 10%). However, confidence intervals include both the possibility of no effect (2 fewer people abstinent per 1000), and the possibility of substantial benefit from CBT (458 more people abstinent per 1000).

f. One study at high risk of bias, two with some concerns, and two at low risk of bias.

g. One study only included individuals referred by the probation service, another only included participants with insomnia related to cannabis use.

h. Sample size is sufficient to meet OIS (estimated at 586 participants, based on completion of treatment 80% in control group, with power of 0.8 to detect a reduction of 10% in intervention group). Confidence intervals indicate that completion of treatment is lower in the intervention group, although the magnitude of this reduction is uncertain (from 26 to 180 fewer per 1000).

i. One study only included individuals referred by the probation service.

j. Optimal information size reached (estimated at 126 participants, for a change in duration from 7 days to 14 days, with a SD of 14 days). Confidence interval excludes the possibility of meaningful benefit (doubling of duration of abstinence) or harm (halving of duration of abstinence).

k. All studies considered to be at high risk of bias for this outcome.

l. $I^2$=83%. Some studies show marked reduction in frequency of use, others show no effect.

m. One study only included people with insomnia related to cannabis use, and another recruited individuals referred by the probation service. However, these studies do not carry substantial weight in the analysis, therefore the certainty of evidence was not reduced for this domain.

n. Sample size exceeds OIS (estimated at 46 participants, based on proportion of use 0.5 in control group, 0.25 in intervention group and SD of 0.3). Confidence intervals for effect include the possibility of substantial benefit from the intervention (more than 50% reduction in use), or of a smaller effect (less than 50% reduction in use).

o. Study considered to be at high risk of bias for this outcome.

p. Sample size exceeds OIS (estimated at 126 participants, based on frequency of use 2 joints per day in control group, 1 joint per day in intervention group and SD of 2). Confidence intervals for effect include the possibility of substantial benefit from the intervention (more than 50% reduction in use), or of a smaller effect (less than 50% reduction in use).

q. Study only included people with insomnia related to cannabis use.

r. Sample size very small (<100 participants). Unable to estimate relative or absolute effects as no events occurred in either group.

**Table 2.** DBT/ACT versus inactive/nonspecific comparator for cannabis use disorder

| Certainty assessment | | | | | | | Number of participants | | Effect | | Certainty |
|---|---|---|---|---|---|---|---|---|---|---|---|
| Studies | Study design | Risk of bias | Inconsistency | Indirectness | Imprecision | Other considerations | DBT/ACT | Inactive/non-specific comparator | Relative [95% CI] | Absolute [95% CI] | |
| **Point abstinence at end of treatment** | | | | | | | | | | | |
| Davoudi 2021a,[8] Davoudi 2021b[9] | randomized trials | serious[a] | not serious | serious[b] | very serious[c] | none | 27/49 (55.1%) | 10/44 (22.7%) | **OR=4.34** [1.74; 10.80] **RR=2.45** [1.49; 3.32] | **333 more per 1,000** [from 111 more to 533 more] | ⊕◯◯◯ Very low[a,b,c] |
| **Completion of treatment** | | | | | | | | | | | |
| Budak 2024,[10] Davoudi 2021a,[8] Davoudi 2021b[9] | randomized trials | serious[d] | not serious[e] | serious[f] | serious[g] | none | 79/90 (87.8%) | 74/91 (81.3%) | **OR=1.42** [0.59; 3.43) **RR=1.06** [0.88; 1.16] | **48 more per 1,000** [from 93 fewer to 124 more] | ⊕◯◯◯ Very low[d,e,f,g] |
| **Frequency of cannabis use at end of treatment** | | | | | | | | | | | |
| Davoudi 2021a,[8] Davoudi 2021b[9] | randomized trials | serious[a] | not serious[h] | serious[b] | serious[i] | none | 50 participants | 51 participants Assumed proportion of days using was 0.5 (i.e. use on 3.5 out of every 7 days) | **RoM=0.39** [0.25; 0.60] | **Proportion of days using was 0.305 fewer** [from 0.375 fewer to 0.2 fewer]. If the control group used cannabis on 3.5 days out of 7, the intervention group would use on 1.4 days out of 7 [from 0.9 days to 2.1 days]. | ⊕◯◯◯ Very low[a,b,h,i] |
| **Cravings at end of treatment (assessed with MCQ-SF)** | | | | | | | | | | | |
| Davoudi 2021a[8] | randomized trials | very serious[j] | not serious | serious[k] | serious[l] | none | 30 participants | 31 participants Mean score 44.48 out of possible 84 | **RoM=0.95** [0.86; 1.04] | **Scores on the MCQ-SF were 2.22 points lower** [from 6.23 lower to 1.78 higher] | ⊕◯◯◯ Very low[j,k,l] |

**ACT,** acceptance and commitment therapy, **CI:** confidence interval; **DBT,** dialectical and behavioural therapy; **MCQ-SF,** Marijuana Cravings Questionnaire-Short Form, **OR:** odds ratio; **RoM,** ratio of means; **RR,** risk ratio.

**Explanations**

a. One study at high risk of bias, one with some concerns.

b. Both studies only recruited males, and all participants had a relatively short duration of cannabis use (less than 2 years).

c. Optimal information size was not reached (estimated at 586 participants), and total sample size is <100.

d. One study at high risk of bias (due to randomization process), two with some concerns regarding risk of bias.

e. $I^2$=36%, confidence intervals are overlapping.

f. All participants were male. For two studies, duration of use was relatively short (less than 2 years).

g. Optimal information size was not reached (estimated at 586 participants).

h. $I^2$=58%. However, direction of effect is consistent, confidence intervals are overlapping, and both studies indicate a meaningful impact of the intervention.

i. Optimal information size was reached (estimated at 46 participants). However, confidence intervals indicate that the frequency of cannabis use may either be reduced by a meaningful amount (>50% reduction), but may also be reduced by a smaller amount (<50% reduction).

j. Study considered to be at high risk of bias.

k. All participants were male, and duration of cannabis use was relatively short (mean of 18 months).

l. Sample size less than the optimal information size (estimated at 126 participants).

**Table 3.** CBT with affect management versus standard CBT for cannabis use disorder

| Certainty assessment | | | | | | | Number of participants | | Effect | | Certainty |
|---|---|---|---|---|---|---|---|---|---|---|---|
| **Studies** | **Study design** | **Risk of bias** | **Inconsistency** | **Indirectness** | **Imprecision** | **Other considerations** | **CBT -affect** | **CBT** | **Relative [95% CI]** | **Absolute [95% CI]** | |
| **Point abstinence at end of treatment** | | | | | | | | | | | |
| Buckner 2019[11] | randomized trials | very serious[a] | not serious | serious[b] | very serious[c] | none | 3/19 (15.8%) | 0/18 (0.0%) | **OR=7.85** [0.38; 163.52] **RR** not estimated* | **462 more per 1,000** [from 113 fewer to 776 more, if control group abstinence assumed to be 20%] | ⊕◯◯◯ Very low[a,b,c] |
| **Completion of treatment** | | | | | | | | | | | |
| Buckner 2019,[11] Wolitzky-Taylor 2022[12] | randomized trials | serious[d] | not serious[e] | serious[f] | serious[g] | none | 37/54 (68.5%) | 36/53 (67.9%) | **OR=1.03** [0.45; 2.32] **RR=1.01** [0.72; 1.22] | **6 more per 1,000** [from 191 fewer to 152 more] | ⊕◯◯◯ Very low[d,e,f,g] |
| **Frequency of cannabis use at end of treatment** | | | | | | | | | | | |
| Wolitzky-Taylor 2022[12] | randomized trials | very serious[a] | not serious | not serious | serious[c] | none | 18 participants | 18 participants Assumed proportion of days using was 0.5 (i.e. use on 3.5 out of every 7 days) | **RoM=0.93** [0.56; 1.55] | **Proportion of days using was 0.04 fewer** [from 0.22 fewer to 0.28 more]. If the control group used cannabis on 3.5 days out of 7, the intervention group would use on 3.25 days out of 7 [from 1.96 days to 5.43 days]. | ⊕◯◯◯ Very low[a,c] |
| **Quantity of cannabis use at end of treatment** | | | | | | | | | | | |
| Buckner 2019[11] | randomized trials | very serious[a] | not serious | serious[b] | serious[h] | none | 19 participants | 18 participants Assumed quantity of use 2 joints per day | **RoM=0.49** [0.17; 1.38] | Quantity of use reduced by 1.02 joints per day [from 1.66 fewer to 0.76 more] | ⊕◯◯◯ Very low[a,b,h] |

* Odds ratio estimated using a continuity correction due to zero events in control group arm; note very wide confidence intervals. We did not consider it appropriate to re-express this result as a risk ratio.

**CBT,** cognitive-behavioural therapy; **CBT-affect**, CBT with affect management; **CI,** confidence interval; **OR,** odds ratio; **RoM,** ratio of means, **RR:** risk ratio.

**Explanations**

a. Study considered to be at high risk of bias for this outcome.

b. All participants were diagnosed with an anxiety disorder as well as cannabis use disorder.
c. Sample size less than optimal information size (estimated at 46 participants).
d. Both studies had some concerns regarding risk of bias for this outcome.
e. $I^2$=0%, confidence intervals overlapping.
f. One study only included participants who were diagnosed with an anxiety disorder as well as CUD.
g. Optimal information size not reached (estimated at 586 participants).
h. Sample size less than optimal information size (estimated at 126 participants).

**Table 4.** CBT plus contingency management for abstinence versus CBT alone for cannabis use disorder

| Certainty assessment | | | | | | | Number of participants | | Effect | | Certainty |
|---|---|---|---|---|---|---|---|---|---|---|---|
| **Studies** | **Study design** | **Risk of bias** | **Inconsistency** | **Indirectness** | **Imprecision** | **Other considerations** | **CBT plus CM-abstinence** | **CBT** | **Relative [95% CI]** | **Absolute [95% CI]** | |
| **Point abstinence at end of treatment** | | | | | | | | | | | |
| Budney 2000[13] | randomized trials | serious[a] | not serious | not serious | very serious[b] | none | 8/20 (40.0%) | 3/20 (15.0%) | **OR=3.78** [0.83; 17.25] **RR=2.67** [0.85; 5.02] | **250 more per 1,000** [from 22 fewer to 603 more] | ⊕◯◯◯ Very low[a,b] |
| **Continuous abstinence at end of treatment** | | | | | | | | | | | |
| Budney 2000,[13] Kadden 2007,[3] Litt 2020[14] | randomized trials | serious[c] | not serious | not serious | serious[d] | none | 37/171 (21.6%) | 22/165 (13.3%) | **OR=1.81** [0.61; 5.41] **RR=1.64** [0.64; 3.44] | **84 more per 1,000** [from 48 fewer to 321 more] | ⊕⊕◯◯ Low[c,d] |
| **Completion of treatment** | | | | | | | | | | | |
| Carroll 2006,[7] Carroll 2012,[15] Kadden 2007,[3] Litt 2020[14] | randomized trials | not serious | not serious[e] | serious[f] | serious[d] | none | 209/229 (91.3%) | 199/230 (86.5%) | **OR=1.58** [0.85; 2.94] **RR=1.05** [0.98; 1.09] | **45 more per 1,000** [from 20 fewer to 84 more] | ⊕⊕◯◯ Low[d,e,f] |
| **Duration of continuous abstinence at end of treatment** | | | | | | | | | | | |
| Budney 2000,[13] Carroll 2006,[7] Carroll 2012,[15] Kadden 2007,[3] Litt 2020[14] | randomized trials | very serious[g] | serious[h] | serious[f] | not serious[i] | none | 194 participants | 194 participants Assumed duration of abstinence 7 days | **RoM=1.40** [1.02; 1.91] | **2.8 days longer** [from 0.14 days longer to 6.37 days more] | ⊕◯◯◯ Very low[f,g,h,i] |
| **Frequency of cannabis use at end of treatment** | | | | | | | | | | | |

| Certainty assessment | | | | | | | Number of participants | | Effect | | Certainty |
|---|---|---|---|---|---|---|---|---|---|---|---|
| Studies | Study design | Risk of bias | Inconsistency | Indirectness | Imprecision | Other considerations | CBT plus CM-abstinence | CBT | Relative [95% CI] | Absolute [95% CI] | |
| Budney 2000,[13] Carroll 2006,[7] Carroll 2012,[15] Kadden 2007,[3] Litt 2020[14] | randomized trials | very serious[g] | not serious[j] | serious[f] | not serious[k] | none | 245 participants | 241 participants<br>Assumed proportion of days of use = 0.5, i.e. 3.5 days out of every 7 days | **RoM=0.88** [0.65; 1.19] | **Proportion of days using lower by 0.06** [from 0.175 lower to 0.095 higher].<br>If the control group used cannabis on 3.5 days out of 7, the intervention group would use on 3.08 days out of 7 [from 2.28 days to 4.17 days]. | ⊕◯◯◯ Very low[f,g,j,k] |

**CBT,** cognitive-behavioural therapy; **CI,** confidence interval; **CM-abstinence**, contingency management based on abstinence; **OR,** odds ratio; **RoM,** ratio of means, **RR:** risk ratio.

**Explanations**

a. Study assessed at high risk of bias for this outcome.

b. Sample size <100 participants.

c. Study with highest weight in the analysis rated at some concerns with risk of bias. Remaining two studies rated at high risk of bias.

d. Sample size less than optimal information size (estimated at 586 participants).

e. $I^2$=6%, confidence intervals overlapping.

f. Two studies recruited participants referred by the criminal justice system.

g. All studies rated at high risk of bias for this outcome.

h. $I^2$=51%, showing moderate heterogeneity. Confidence intervals are overlapping, but direction of effect does differ for one trial.

i. Sample size exceeds optimal information size (estimated at 126 participants). Confidence intervals exclude the possibility of a meaningful effect (either a doubling or halving of duration of abstinence).

j. $I^2$=39%. Confidence intervals overlapping, however, effect sizes from all trials indicate that meaningful effect sizes are not reached.

k. Sample size exceeds optimal information size (estimated at 46 participants). Confidence intervals exclude the possibility of a meaningful effect (either a doubling or halving of duration of abstinence).

**Table 5.** CBT plus contingency management for abstinence versus CBT plus contingency management for attendance for cannabis use disorder

| Certainty assessment | | | | | | | Number of participants | | Effect | | Certainty |
|---|---|---|---|---|---|---|---|---|---|---|---|
| **Studies** | **Study design** | **Risk of bias** | **Inconsistency** | **Indirectness** | **Imprecision** | **Other considerations** | **CBT plus CM-abstinence** | **CBT plus CM-attendance** | **Relative [95% CI]** | **Absolute [95% CI]** | |
| **Point abstinence at end of treatment** | | | | | | | | | | | |
| Budney 2006,[16] Stanger 2009[17] | randomized trials | serious[a] | not serious | serious[b] | serious[c] | none | 33/59 (55.9%) | 31/56 (55.4%) | **OR=1.61** [0.72; 3.60] **RR=1.21** [0.85; 1.48] | **113 more per 1,000** [from 82 fewer to 263 more] | ⨁◯◯◯ Very low[a,b,c] |
| **Continuous abstinence at end of treatment** | | | | | | | | | | | |
| Budney 2006,[16] Litt 2013,[18] Stanger 2009[17] | randomized trials | very serious[d] | serious[e] | not serious[f] | serious[c] | none | 41/119 (34.5%) | 25/117 (21.4%) | **OR=2.04** [0.75; 5.58] **RR=1.67** [0.79; 2.84] | **143 more per 1,000** [from 44 fewer to 389 more] | ⨁◯◯◯ Very low[c,d,e,f] |
| **Completion of treatment** | | | | | | | | | | | |
| Budney 2006,[16] Carroll 2012,[15] Stanger 2009[17] | randomized trials | serious[g] | not serious | serious[b,h] | serious[c] | none | 86/98 (87.8%) | 82/95 (86.3%) | **OR=1.12** [0.48; 2.62] **RR=1.02** [0.87; 1.09] | **13 more per 1,000** [from 111 fewer to 80 more] | ⨁◯◯◯ Very low[b,c,g,h] |
| **Duration of continuous abstinence at end of treatment** | | | | | | | | | | | |
| Budney 2006,[16] Carroll 2012,[15] Litt 2013,[18] Stanger 2009[17] | randomized trials | very serious[d] | serious[i] | serious[b,h] | not serious[j] | none | 158 participants | 156 participants Assumed duration of abstinence 7 days | **RoM=1.31** [0.97; 1.77] | **2.17 days longer** [from 0.21 days shorter to 5.39 days more] | ⨁◯◯◯ Very low[b,d,h,i,j] |
| **Frequency of use at end of treatment** | | | | | | | | | | | |

| Certainty assessment | | | | | | | Number of participants | | Effect | | Certainty |
|---|---|---|---|---|---|---|---|---|---|---|---|
| Studies | Study design | Risk of bias | Inconsistency | Indirectness | Imprecision | Other considerations | CBT plus CM-abstinence | CBT plus CM-attendance | Relative [95% CI] | Absolute [95% CI] | |
| Budney 2006,[16] Carroll 2012,[15] Litt 2013,[18] Stanger 2009[17] | randomized trials | very serious[d] | serious[k] | serious[b,h] | not serious[l] | none | 158 participants | 156 participants Assumed proportion of days using was 0.5 (i.e. use on 3.5 out of every 7 days) | **RoM=0.98** [0.68; 1.40] | **Proportion of days using was 0.01 fewer** [from 0.16 fewer to 0.2 more]. If the control group used on 3.5 days out of 7, the intervention group would use on 3.43 days out of 7 [from 2.38 days to 4.9 days]. | ⊕◯◯◯ Very low[b,d,h,k,l] |
| **Adverse effects** | | | | | | | | | | | |
| Stanger 2009[17] | randomized trials | serious[m] | not serious | serious[b] | very serious[n] | none | 0/28 (0%) | 0/26 (0%) | Not estimable | No adverse events occurred in either group. | ⊕◯◯◯ Very low[b,m,n] |

**CBT,** cognitive-behavioural therapy; **CI,** confidence interval; **CM-abstinence**, contingency management based on abstinence; **CM-attendance**, contingency management based on attendance; **OR,** odds ratio; **RoM,** ratio of means; **RR,** risk ratio.

**Explanations**

a. One study considered to be at high risk of bias, one with some concerns.

b. One study included only younger participants, aged 12-18 years.

c. Sample size less than optimal information size (estimated at 586 participants).

d. All studies considered to be at high risk of bias for this outcome.

e. $I^2$=52%. Confidence intervals do overlap, but magnitude and direction of effect does vary (one study indicating significant benefit, one indicating no effect).

f. Although one study included only younger participants, this contributes relatively small weight to the meta-analysis, therefore not downgraded for indirectness.

g. One study considered at high risk of bias, two studies with no concerns.

h. One study included a majority of participants who were referred by the criminal justice system.

i. $I^2$=40%. Direction of effect does vary, but all studies show results which indicate a very small impact on duration of abstinence, therefore heterogeneity does not impact on conclusions.

j. Sample size exceeds the optimal information size (estimated at 126 participants). Confidence intervals exclude the possibility of a meaningful difference (either a doubling or halving of duration of abstinence).

k. $I^2$=62%. Direction of effect does vary, but all studies show results which indicate a very small impact on frequency of use, therefore heterogeneity does not impact on conclusions.

l. Sample size exceeds the optimal information size (estimated at 46 participants). Confidence intervals exclude the possibility of a meaningful difference (either a doubling or halving of frequency of use).

m. Study assessed as having some concerns regarding risk of bias.

n. Sample size very small (<100 participants). Unable to estimate relative or absolute effects as no events occurred in either group.

**Table 6.** Multidimensional family therapy versus CBT for cannabis use disorder

| Certainty assessment | | | | | | | Number of participants | | Effect | | Certainty |
|---|---|---|---|---|---|---|---|---|---|---|---|
| Studies | Study design | Risk of bias | Inconsistency | Indirectness | Imprecision | Other considerations | Multidimensional family therapy | CBT | Relative [95% CI] | Absolute [95% CI] | |
| **Frequency of cannabis use at end of treatment** | | | | | | | | | | | |
| Rigter 2013[19] | randomized trials | very serious[a] | not serious | serious[b] | serious[c] | none | 212 participants | 238 participants Assumed proportion of days of use = 0.5, i.e. 3.5 days out of every 7 days | **RoM=0.81** [0.69; 0.95] | **Proportion of days using lower by 0.1** [from 0.16 lower to 0.03 lower], i.e. a reduction in use to 2.8 days out of every 7 [from 2.4 days to 3.3 days] | ⊕◯◯◯ Very low[a,b,c] |

**CBT,** cognitive-behavioural therapy; **CI,** confidence interval; **RoM,** ratio of means.

**Explanations**

a. Study considered to be at high risk of bias for this outcome.

b. Study conducted in multiple countries, and comparator group included different interventions across the locations. All included some CBT, but additional treatments were available for some participants. All participants were aged 13-18 years.

c. Sample size exceeds optimal information size (estimated at 46 participants). Confidence intervals exclude the possibility of significant benefit (halving of cannabis use) or harm (doubling of cannabis use).

**Table 7.** Community reinforcement versus alternative intervention for cannabis use disorder

| Certainty assessment | | | | | | | Number of participants | | Effect | | Certainty |
|---|---|---|---|---|---|---|---|---|---|---|---|
| **Studies** | **Study design** | **Risk of bias** | **Inconsistency** | **Indirectness** | **Imprecision** | **Other considerations** | **Community reinforcement** | **Alternative intervention** | **Relative [95% CI]** | **Absolute [95% CI]** | |
| **Point abstinence at end of treatment** | | | | | | | | | | | |
| Kaminer 2017[20] | randomized trials | very serious[a] | not serious | serious[b] | very serious[c] | none | 2/13 (15.4%) | 5/13 (38.5%) | **OR=0.29** [0.04; 1.90] **RR=0.40** [0.07; 1.41] | **231 fewer per 1,000** [from 360 fewer to 158 more] | ⊕◯◯◯ Very low[a,b,c] |
| **Continuous abstinence at end of treatment** | | | | | | | | | | | |
| Khalily 2023[21] | randomized trials | very serious[a] | not serious | not serious | serious[d] | none | 51/59 (86.4%) | 7/59 (11.9%) | **OR=47.36** [16.00; 140.21] **RR=7.22** [4.40; 8.12] | **746 more per 1,000** [from 564 more to 831 more] | ⊕◯◯◯ Very low[a,d] |
| **Completion of treatment** | | | | | | | | | | | |
| Kaminer 2017,[20] Khalily 2023[21] | randomized trials | serious[e] | not serious | serious[f] | not serious | none | 72/95 (75.8%) | 72/100 (72.0%) | **OR=1.20** [0.49; 2.96] **RR=1.05** [0.77; 1.23] | **35 more per 1,000** [from 162 fewer to 164 more] | ⊕⊕◯◯ Low[e,f] |

**CI** confidence interval; **OR,** odds ratio; **RR,** risk ratio.

**Explanations**

a. Study assessed at high risk of bias for this outcome.

b. All participants had failed to achieve abstinence during a seven week motivational-enhancement therapy and CBT course before entering this study. Some participants were aged less than 16 years.

c. Sample size <100 participants.

d. Sample size fails to meet optimal information size (estimated at 586 participants).

e. One study assessed at high risk of bias, and one with some concerns for this outcome.

f. One study only included participants who had already failed to achieve abstinence in a previous trial.
