## Supplementary material for "Effectiveness and safety of psychosocial interventions for the treatment of cannabis use disorder: a systematic review and meta-analysis": SuppInfo13

### SUPPORTING INFORMATION 12. ECONOMIC EVALUATION STUDIES

We identified two economic evaluations based on the included trials. Relevant cost and effectiveness data are presented in Table 1 (numbering of tables is specific to this Supporting Information document). Goorden 2016[1] analysed cost-utility and cost-effectiveness of multidimensional family therapy (MDFT) compared with cognitive-behavioural therapy (CBT) for adolescents, based on a single Dutch site of a multicentre trial.[2] The evaluation was conducted using a healthcare perspective over a 12-month time horizon. Results were reported as cost per quality-adjusted life years (QALYs) gained using the EuroQol 5 Dimensions (EQ-5D-3L), and cost per recovered patient, defined as living in the community and abstinent from cannabis and other substances in the past 30 days. Costs were expressed as direct medical costs for self-reported number of contacts with healthcare providers in the three months prior to the assessment. MDFT was more costly but also more effective than CBT in terms of quality of life, whereas improvement in recovery was not statistically significant. Incremental cost-effectiveness ratios (ICERs) are reported in Table 1. The probability that MDFT would be cost-effective reached 90% at €100,000 EUR per QALYs gained. A secondary analysis which included costs of delinquency (for the self-reported type and number of crimes in the three months prior to the assessment) reduced the ICERs (see Table 1). The probability that MDFT would be cost-effective then reached 80% at €100,000 EUR per QALYs gained.

Olmstead 2007[3] evaluated cost-effectiveness of CBT plus contingency management (CM) based on abstinence/attendance, CBT alone, CM-abstinence/attendance, and counselling (nonspecific comparator) for young adults referred by the criminal justice system, based on a multi-arm trial[4] from the United States. The primary cost and effectiveness outcomes were assessed over the eight-week treatment period, and additional analysis was conducted over an eight-month time horizon, both using a healthcare perspective. Effectiveness was defined as the longest duration of continuous abstinence (based on urine tests at end of treatment, and self-reported at follow-up), and as the number of cannabis-negative urine samples. Costs were expressed as clinic costs including labour costs, urinalysis materials, and voucher system for CM. While the interventions did not differ significantly in effectiveness, the costs were the highest for CBT plus CM, followed by CM, CBT, and counselling. ICERs are reported in Table 1. CM-abstinence/attendance was excluded from the following ICER calculations: for the longest duration of continuous abstinence at follow-up, as CM-abstinence/attendance was strictly dominated by CBT (CBT was less costly and more effective); and for the number of negative urine samples, as CM-abstinence/attendance was extendedly dominated (the ICER was greater than the next, more effective, treatment option) by the combination of CBT and CBT plus CM-abstinence/attendance.

References relating to this Supporting Information are included at the end of this document.

**Table 1.** Cost-effectiveness outcomes

| Study; country | Perspective; time horizon | Intervention | Comparator | Cost measure | Intervention cost | Comparator cost | Effectiveness measure | Intervention effectiveness | Comparator effectiveness | ICER | Uncertainty analysis |
|---|---|---|---|---|---|---|---|---|---|---|---|
| Goorden 2016[1] (based on Rigter 2013[2] single site); The Netherlands | Healthcare; 12 months | MDFT (n=49)[a] | CBT (n=47) | Direct medical costs, price year 2012 (mean [95% CI]) | €5446 [4159 to 7092] EUR | €2015 [1397 to 2714] EUR | Change in health-related quality of life (EQ-5D-3L)[c] over 12 months (mean [95% CI]) | 0.04 [0.03; 0.06] | -0.2 [-0.05; 0.02] | €54,308 EUR/QALYs gained | Probabilistic (bootstrapping, cost-effectiveness acceptability curve) |
| | Healthcare + delinquency; 12 months | MDFT (n=49) | CBT (n=47) | Direct medical costs + delinquency costs, price year 2012 (mean [95% CI]) | €21,915 [16,273 to 28,181] EUR | €21,330 [12,389 to 32,894] EUR | Change in health-related quality of life (EQ-5D-3L)[c] over 12 months (mean [95% CI]) | 0.04 [0.03; 0.06] | -0.2 [-0.05; 0.02] | €9266 EUR/QALYs gained | Probabilistic (bootstrapping, cost-effectiveness acceptability curve); Sensitivity analysis (excluding costs of traffic offences), ICER=€65,823 EUR/QALYs gained |
| | Healthcare; 12 months | MDFT (n=49) | CBT (n=47) | Direct medical costs, price year 2012 (mean [95% CI]) | €5446 [4159 to 7092] EUR | €2015 [1397 to 2714] EUR | Proportion of recovered[d] patients at 12 months | 14.30% | 6.40% | €43,405 EUR/ recovered patient | |
| | Healthcare + delinquency; 12 months | MDFT (n=49) | CBT (n=47) | Direct medical costs + delinquency costs, price year 2012 (mean [95% CI]) | €21,915 [16,273 to 28,181] EUR | €21,330 [12,389 to 32,894] EUR | Proportion of recovered [d] patients at 12 months | 14.30% | 6.40% | €7491 EUR/ recovered patient | |
| Olmstead 2007[3] (based on Carrol 2006);[4] United States | Healthcare; 8 weeks | MET/CBT (n=32)[b] | NS (n=32) | Clinic costs, price year NR (mean, SD) | $305 (130) USD | $243 (124) USD | Longest duration of continuous abstinence[e] over 8 weeks (mean) | 3.08 weeks | 2.47 weeks | $102 USD/ additional week of abstinence | Probabilistic (bootstrapping, cost-effectiveness acceptability curve); Scenario (assuming alternative costs), ICER=$77 USD/additional week of abstinence |
| | Healthcare; 8 weeks | CM-ab-at (n=32) | MET/CBT (n=32) | Clinic costs, price year NR (mean, SD) | $1078 (352) USD | $305 (130) USD | Longest duration of continuous abstinence over 8 weeks (mean) | 3.78 weeks | 3.08 weeks | $1104 USD /additional week of abstinence | Probabilistic (bootstrapping, cost-effectiveness acceptability curve); Scenario (assuming alternative costs), |

| Study; country | Perspective; time horizon | Intervention | Comparator | Cost measure | Intervention cost | Comparator cost | Effectiveness measure | Intervention effectiveness | Comparator effectiveness | ICER | Uncertainty analysis |
|---|---|---|---|---|---|---|---|---|---|---|---|
| | | | | | | | | | | | ICER=$766 USD/additional week of abstinence |
| | Healthcare; 8 weeks | MET/CBT/CM-ab-at (n=33) | CM-ab-at (n=32) | Clinic costs, price year NR (mean, SD) | $1238 (397) USD | $1078 (352) USD | Longest duration of continuous abstinence over 8 weeks (mean) | 3.90 weeks | 3.78 weeks | $1333 USD /additional week of abstinence | Probabilistic (bootstrapping, cost-effectiveness acceptability curve); Scenario (assuming alternative costs), ICER=$942 USD/additional week of abstinence |
| | Healthcare; 8 months | MET/CBT (n=27) | NS (n=24) | Clinic costs, price year NR (mean, SD) | $305 (130) USD | $243 (124) USD | Longest duration of continuous abstinence[f] at 8 months (mean) | 9.56 weeks | 7.73 weeks | $34 USD/ additional week of abstinence [g] | Probabilistic (bootstrapping, cost-effectiveness acceptability curve) |
| | Healthcare; 8 months | MET/CBT/CM-ab-at (n=27) | MET/CBT (n=27) | Clinic costs, price year NR (mean, SD) | $1238 (397) USD | $305 (130) USD | Longest duration of continuous abstinence[f] at 8 months (mean) | 10.58 weeks | 9.56 weeks | $915 USD/ additional week of abstinence [g] | Probabilistic (bootstrapping, cost-effectiveness acceptability curve) |
| | Healthcare; 8 weeks | MET/CBT (n=32) | NS (n=32) | Clinic costs, price year NR (mean, SD) | $305 (130) USD | $243 (124) USD | Number of cannabis-negative urine samples over 8 weeks (mean) | 1.27 | 0.88 | $159 USD/ additional negative sample[h] | Probabilistic (bootstrapping, cost-effectiveness acceptability curve); Scenario (assuming alternative costs), ICER=$121 USD/additional negative sample |
| | Healthcare; 8 weeks | MET/CBT/CM-ab-at (n=33) | MET/CBT (n=32) | Clinic costs, price year NR (mean, SD) | $1238 (397) USD | $305 (130) USD | Number of cannabis-negative urine samples over 8 weeks (mean) | 2.26 | 1.27 | $942 USD/ additional negative sample[h] | Probabilistic (bootstrapping, cost-effectiveness acceptability curve); Scenario (assuming alternative costs), ICER=$656 USD/additional negative sample |

[a] 12% of participants across study arms were excluded from this analysis due to insufficient costs and effectiveness data.

[b] 2% of participants across study arms were excluded from the analysis at end of treatment assessment, and 6% were missing at 6-months follow-up.

[c] Quality of life was measured using EQ-5D-3L, as change from baseline to the 12-months assessment (6 months post-treatment), and linked to empirical valuations of the Dutch general public.

[d] Recovery was defined as living in the community and abstinent from cannabis, heavy alcohol use, and any other substance use in the 30 days prior to the 12 month assessment (6 months post-treatment).

[e] Continuous abstinence determined by weekly consecutive urine tests negative for cannabinoids, over 8 weeks treatment period; adjusted for baseline level of cannabis use.

[f] Continuous abstinence determined by self-report of daily cannabis use collected at 6 months post-treatment; adjusted for baseline level of cannabis use.
[g] For the longest duration of continuous abstinence at follow-up, CM-ab-at was strictly dominated by CBT, as CBT was less costly and more effective, therefore, CM-ab-at was excluded from the ICER calculations.
[h] For the number of negative urine samples, CM-ab-at was extendedly dominated (the ICER is greater than the next, more effective, treatment option) by the combination of CBT and CBT plus CM-ab-at, therefore, CM-ab-at was excluded from the ICER calculations.
CBT, cognitive behavioural therapy; CI, confidence interval; CM-ab-at, contingency management based on abstinence and attendance; EQ-5D-3L, EuroQol 5 Dimensions 3 Levels; EUR, Euros; ICER, incremental cost-effectiveness ratio; MDFT, multidimensional family therapy; MET, motivation enhancement therapy; n, number of participants analysed; NR, not reported; NS, nonspecific comparator (in this instance, counselling); SD, standard deviation; QALY, quality-adjusted life years; USD, United States dollars.

#### Critical appraisal for economic evaluation studies

Critical appraisal[5] of the two economic evaluation studies did not identify major concerns. Goorden 2016[1] was missing separately reported quantities of resources (therapy contacts). Olmstead 2007[3] did not report whether/how valuation of health benefit as an additional week of continuous abstinence was conducted; price year for the estimates of unit costs or details of any currency or price adjustments; and confidence intervals for effectiveness measures and ICERs. Both studies lacked an explanation for not discounting the costs of benefits, although discounting would only be applicable to a time horizon of more than one year. There were no other issues related to study design, data collection, or analysis and interpretation of results.
