## Supplementary material for "Effectiveness and safety of psychosocial interventions for the treatment of cannabis use disorder: a systematic review and meta-analysis": SuppInfo01

### SUPPORTING INFORMATION 1. INTERVENTION GROUPINGS

Here we provide a description of the intervention types and groupings. Abbreviations are those used in the main article, tables and forest plots and throughout the Supporting Information documents. References relating to this Supporting Information are at the end of this document.

*CBT*

Therapies using cognitive-behavioural techniques such as cognitive restructuring, behavioural self-monitoring, coping skills, problem solving and decision-making skills. The majority of studies (n = 11) implemented cognitive-behavioural therapy (CBT) targeting issues related to cannabis use (e.g. cognitive restructuring and skills training to understand patterns of use and identify high risk situations, deal with craving or lapses, maintain abstinence, promote assertiveness or manage negative emotions). One study used CBT to target insomnia related to cannabis use.[1] CBT was delivered in combination with elements of motivation enhancement therapy (MET), such as motivational interviewing and developing commitment to change. Other interventions included in the CBT category were a combination of MET and behavioural therapy focusing on coping skills,[2] and relapse prevention using MET, cognitive restructuring, and skills training focused on cannabis use.[3,4] Note that one of the CBT interventions was described as treatment-as-usual that differed between the sites of a multi-centre trial[5] but each protocol included common MET and CBT elements and individual substance abuse counselling.
Duration: 1.5-6 months (mean [M]=2.95, standard deviation [SD]=1.09). Number of sessions: 6-14 (M=9.86, SD=2.48).

*CBT + Affect*

Two studies evaluated CBT-based interventions with a specific emphasis on affect management: integrated cannabis and anxiety reduction treatment (ICART)[6] and affect management treatment (AMT).[7] These treatments targeted false safety behaviours, avoidance, negative urgency, distress intolerance and misappraisal.
Duration: 3 months. Number of sessions: 12.

*DBT/ACT*

Third/fourth-wave psychotherapies included dialectical behavioural therapy (DBT),[8] acceptance and commitment therapy (ACT)[9] and mindfulness-based psychoeducation,[10] implemented in one study each. These interventions included elements of cannabis use focused psychoeducation as well as training in mindfulness, emotion regulation, distress tolerance, interpersonal skills, problem solving and acceptance.
Duration: 1-4 months (M=2.67, SD=1.53). Number of sessions: 8-16 (M=12, SD=4).

*CM-abstinence*

Four studies used contingency management (CM) based on abstinence as a standalone intervention.[11-14] Participants received lottery draws or vouchers for providing cannabinoid-negative urine specimens, usually starting from week 2 of treatment (allowing for a sufficient wash-out period). The value of potential rewards varied from approximately $1 to $100 USD, and typically increased over the duration of the study, according to the length of continuous abstinence. Providing a urine test positive for cannabinoids reset the rewards to the baseline level. Note that Carroll 2006[12] used a combination of CM based on abstinence and attendance, however, rewards for each were independent and this intervention was classified as CM-

abstinence assuming that it would be a stronger therapeutic component.
Duration: 2-3.5 months (M=2.69, SD=0.69). Number of sessions: 8-29 (M=14.5, SD=9.81).

*CBT + CM-abstinence*

In eight studies, CM-abstinence was delivered in addition to CBT, both as described above.
Duration: 2-3.5 months (M=2.84, SD=0.67). Number of sessions: 8-43 (M=20, SD=15.34).

*CBT + CM-attendance*

CM based on attendance was delivered in addition to CBT in four studies. Participants gained rewards for attending intervention sessions, providing urine samples (regardless of the test result), and/or homework completion. Rewards were lottery draws for prizes ($1-100 USD value, amount of draws escalating with consecutive attendance)[13,15] or fixed-amount vouchers ($5 USD).[11,16]
Duration: 2-3.5 months (M=3, SD=0.71). Number of sessions: 9-41 (M=19.25, SD=15.31).

*ComReinf*

Community reinforcement focused on reducing environmental contingencies that maintain cannabis use and finding new (or enhancing existing) reinforcers for staying abstinent. Interventions involved existing community resources and developing new positive support systems. They included functional analysis and behavioural skills training (such as communication, problem solving, social skills, vocational training).[17,18]
Duration: 1.5-2.5 months (M=2, SD=0.71). Number of sessions: 6-10 (M=8, SD=2.83).

*MDFT*

We considered multidimensional family therapy (MDFT) as a standalone intervention, distinct from the above categories. MDFT focuses on improving multiple life domains, including adolescents' developmental and intrapersonal issues, individual functioning of their parents and parenting style, the broader family environment, and community systems (such as school, social services, criminal justice), through blended family and individual therapy, drug counselling, and system-oriented interventions.[5]
Duration: 6 months. Number of sessions: 52.

*Inactive/nonspecific comparators*

Inactive comparator represents waitlist control conditions,[3,6,19-21] where no intervention was provided to participants during the waitlist period, but they were offered an alternative intervention after that period. None of the included studies used 'no intervention' control. Nonspecific comparators aim to control for the common features of therapies such as support or educational content but they do not include training in techniques thought of as being therapeutic. [22,23] These included social support,[4] case management,[14,15] and sham sleep improvement treatment.[1] Even though we pre-specified counselling and education as example interventions of interest, the included studies that used these conditions described them as routine care, therefore, we classified them as nonspecific comparators.[8,9,12,18] However, in some settings, a nonspecific comparator may be described as routine care but be considered an active intervention in another setting. Where possible, classification of interventions and comparators was based on author reported content. If insufficient detail was reported, the trialists' definition was used (e.g., if 'standard care' arm received a course of CBT, it was classified as CBT and not as a nonspecific comparator).[5]

Duration and number of sessions (not applicable to waitlist) were typically matched to the active intervention within the same study.
