## Supplementary material for "Effectiveness and safety of psychosocial interventions for the treatment of cannabis use disorder: a systematic review and meta-analysis": SuppInfo02

### SUPPORTING INFORMATION 2. OPERATIONALIZATION OF OUTCOMES AND HIERARCHY OF PREFERENCE FOR OUTCOME MEASURES AND TIMEPOINTS

#### Operationalization and explanation of outcomes

Primary outcomes:

(1) point abstinence at end of treatment - proportion of participants abstinent at the time of assessment, with abstinence determined by urine tests or self-report;
(2) continuous abstinence at the end of treatment - the proportion of participants with a specified duration of continuous abstinence up until the time of assessment, with abstinence determined as above;
(3) intensity of withdrawal and/or craving - self-reported or assessed by clinicians;
(4) completion of scheduled treatment - proportion of participants attending the end of treatment assessment (regardless of treatment adherence);
(5) adverse events - number of participants with any adverse events (i.e. unfavourable or harmful health-related outcomes that occur during/after the intervention but are not necessarily caused by it) at any time (up to the latest reported follow-up), and the nature of adverse events; and
(6) dropout due to adverse events - number of participants withdrawn from the trial due to adverse events, at any time.

Secondary outcomes:

(1) point abstinence at medium follow-up (up to six months post-treatment) and long follow-up (over six months post-treatment);
(2) continuous abstinence at medium and long follow-up;
(3) duration of the longest continuous abstinence over the treatment period and medium and long follow-up (complementary to continuous abstinence, but not pre-specified in the review protocol);
(4) level of cannabis use - (a) frequency and (b) quantity of use measured by self-report or urine tests at the end of treatment, medium, and long follow-up;
(5) number of participants engaging in further treatment following completion of the intervention, at any time post-treatment; and
(6) economic outcomes - incremental cost and effectiveness measures.

#### Hierarchy of preference

For studies that reported multiple eligible measures of the same outcome, we followed a hierarchy of preference to include a single measure within each outcome category.

Where abstinence or level of cannabis use were measured both via urine drug tests and self-report, we only used the results of urine tests.

For studies that reported proportion of participants with continuous abstinence for different durations (e.g. six weeks of continuous abstinence versus 10 weeks of continuous abstinence), we used the longest reported period leading up to the end of treatment or follow-up timepoint.

Where studies reported craving separately from withdrawal symptoms, we treated these as distinct outcomes.

Self-reported level of cannabis use was reported in terms of frequency and quantity of use, and we treated these as two distinct 'level of use' outcomes. For studies reporting multiple measures of level of use, we included up to one frequency measure and up to one quantity measure at each eligible timepoint, according to the following hierarchy of preference:

- Frequency: (1) number of days/weeks used, (2) proportion of days used, (3) number of periods or times per use day, (4) categorical frequency as daily, weekly, monthly use;
- Quantity: number of (1) grams, (2) standard cannabis units (e.g. three cones or a joint containing 0.5 grams), (3) total cannabis units consumed, (4) joints/cones, (5) waterpipes.

The timepoints of interest in this review were (1) end of treatment (i.e. following the last intervention session), (2) medium follow-up (up to six months from the end of treatment), and (3) long follow-up (over six months from the end of treatment). For studies that reported multiple follow-up timepoints, we used the longest available timepoint within each medium- and long-term category of interest.

End of treatment was a pre-specified primary timepoint for abstinence outcomes, and medium- and long-term follow-up were pre-specified secondary timepoints for abstinence outcomes.
