## Supplementary material for "Effectiveness and safety of psychosocial interventions for the treatment of cannabis use disorder: a systematic review and meta-analysis": SuppInfo03

### SUPPORTING INFORMATION 3. SEARCH STRATEGIES

#### Main search strategies

**Ovid MEDLINE(R) ALL** <1946 to June 12, 2024>

1 Marijuana Abuse/ 7227
2 Marijuana Smoking/dt, th 100
3 ((cannabi* or marihuana or marijuana or bhang or ganja or ganjah or hashish) adj3 (problem* use* or overus* or abus* or misus* or dependen* or addict* or use* disorder* or overdos or craving* or cessation* or detox* or withdraw* or abstin*)).tw,id. 6114
4 ((cannabi* or marihuana or marijuana or bhang or ganja or ganjah or hashish) adj3 (high* or heavy or freq*) adj3 (use* or overuse* or risk* or smok*)).tw,kf. 3305
5 (CUD and (cannabi* or marihuana or marijuana or bhang or ganja or ganjah or hashish)).tw,kf.716
6 ((substance adj2 disorder*) and (cannabi* or marihuana or marijuana or bhang or ganja or ganjah or hashish)).ti,kf,hw. 6925
7 (((relaps* adj1 prevent*) or maintenance) adj5 (cannabi* or marihuana or marijuana or bhang or ganja or ganjah or hashish)).tw,kf. 94
8 or/1-7 17209
9 exp Randomized Controlled Trial/ 616616
10 randomized controlled trial.pt.615041
11 (randomi#ed or randomi#ation or randomi#ing).tw,kf. 867120
12 (RCT or "at random" or (random* adj3 (administ* or allocat* or assign* or class* or cluster or crossover or cross-over or control* or determine* or divide* or division or distribut* or expose* or fashion or number* or place* or pragmatic or quasi or recruit* or split or substitut* or treat*))).tw,kf. 774763
13 Random Allocation/ 107314
14 randomly.ab. 435329
15 double-blind method/ or single-blind method/ 211424
16 ((single or double or triple or treble) adj2 (blind* or mask* or dummy)).tw,kf. 207158
17 trial.ti. 311159
18 (placebo and control* and trial).ab,kf. 71132
19 (placebo adj5 (control* or group*)).tw,kf. 150098
20 or/9-19 1576036
21 8 and 20 1330
22 ((allocat* or assign*) and (group* or control*)).tw,kf. and (Marijuana Abuse/dt, th or Marijuana Smoking/dt, th) 58
23 21 or 22 1335
24 exp animals/ not humans.sh. 5230386
25 ((animal model* or mouse or mice or murine* or rat or rats or rodent* or muridae or murids or rabbit* or leporine* or leporidae or guineapig* or cavies or caviidae or hamster* or cricetidae or gerbil* or gerbillinae or cat or cats or feline* or felidae or dog or dogs or canine* or canidae or pig or pigs or piglet* or minipig* or swine* or porcine* or suidae or horse or horses or donkey or donkies or burros or asses or equine* or equidae or sheep or lamb or lambs or ovine or ovidae or goat or goats or cow or cows or cattle or bovine* or bovidae or primate* or monkey or monkeys or macaque or macaques or marmoset or marmosets) not human*).ti. 2474021
26 24 or 25 5641233
27 23 not 26 1326
28 (review not randomized controlled trial).pt. 3333282
29 ((systematic review or meta analysis) not randomized controlled trial).pt. 349813
30 27 not (28 or 29) 1175
31 Psychosocial Intervention/ 1169

32 (psychosocial* or psycho-social*).tw,kf. 134314
33 exp Psychotherapy/ 224250
34 (psychotherap* or psycho-therap*).tw,kf. 56450
35 (psychological adj3 (therap* or train* or treat* or trial* or intervention*)).tw,kf. 26785
36 (((behavi* or cognitive) adj3 (therap* or train* or treat* or trial* or intervention*)) or CBT*).tw,kf. 120326
37 behavi* activation.tw,kf. 2671
38 exp Biofeedback, Psychology/ 13356
39 (biofeedback or bio-feedback).tw,kf. 8721
40 exp Counseling/ 49928
41 counsel?ing.tw,kf. 125062
42 exp conditioning, psychological/ or avoidance learning/ 75311
43 (avoidance adj (activit* or learning or train*)).tw,kf. 3273
44 (avoid* adj3 (drug* or cannabi* or marihuana or marijuana or bhang or ganja or ganjah or hashish)).tw,kf. 4456
45 *Motivation/ 34045
46 directive counseling/ or motivational interviewing/ 5106
47 (motivation* adj3 (interview* or enhanc* or therap* or treat* or trial* or intervention*)).tw,kf. 12587
48 (behavio* adj3 (chang* or modif*)).tw,kf. 103651
49 Mindfulness/ 6987
50 (mindful?ness* or (mind adj3 train*)).tw,kf. 14844
51 exp reinforcement, psychology/ or exp reward/ 63218
52 (contingen* adj (manag* or reinforc*)).tw,kf. 1704
53 community reinforcement*.tw,kf. 232
54 exp Psychotherapy, Group/ 28023
55 ((family or families or social) adj3 (support* or therap* or intervention*)).tw,kf. 115303
56 (adaptive adj3 (treatment* or intervention*)).tw,kf. 2175
57 ((monetary or voucher* or prize*) adj5 (incentiv* or contingen* or reinforc* or reward* or treat* or trial* or intervention*)).tw,kf. 4682
58 (vouchers or prizes or rewards).tw,kf. 18571
59 (incentive* or reward* or reinforcement* or motivation* or contingent*).ti,kf. 74896
60 (brief adj3 (therap* or train* or treat* or trial* or intervention*)).tw,kf. 17031
61 Marijuana Abuse/th or Marijuana Smoking/th 566
62 Substance Withdrawal Syndrome/th 1009
63 intervention.ti. or ((cannabi* or marihuana or marijuana or bhang or ganja or ganjah or hashish or addict* or depend* or use* disorder*) adj5 intervention*).ab,kf. 133190
64 or/31-63 1051049
65 30 and 64 582

**Ovid APA PsycInfo** <1806 to June Week 1 2024>
1 *"cannabis use"/ 3527
2 "cannabis use disorder"/ 882
3 (exp Cannabis/ or "Cannabis Use"/) and (Drug Abuse/ or Drug Dependency/ or "Substance Use Disorder"/) 3761
4 ((cannabi* or marihuana or marijuana or bhang or ganja or ganjah or hashish) adj3 (problem* use* or overus* or abus* or misus* or dependen* or addict* or use* disorder* or overdos or craving* or cessation* or detox* or withdraw* or abstin*)).tw,id. 5025

5 ((cannabi* or marihuana or marijuana or bhang or ganja or ganjah or hashish) adj3 (high* or heavy or freq*) adj3 (use* or overuse* or risk* or smok*)).tw,id. 2820
6 (CUD and (cannabi* or marihuana or marijuana or bhang or ganja or ganjah or hashish)).tw,id. 541
7 (((substance adj2 disorder*) or substance use* treatment*) and (cannabi* or marihuana or marijuana or bhang or ganja or ganjah or hashish)).ti,id,hw. 1307
8 (((relaps* adj1 prevent*) or maintenance) adj5 (cannabi* or marihuana or marijuana or bhang or ganja or ganjah or hashish)).tw,id. 94
9 or/1-8 11712
10 randomized controlled trials/ 1077
11 (randomi#ed or randomi#ation or randomi#ing).tw,id. 116232
12 (RCT or "at random" or (random* adj3 (administ* or allocat* or assign* or class* or cluster or crossover or cross-over or control* or determine* or divide* or division or distribut* or expose* or fashion or number* or place* or pragmatic or quasi or recruit* or split or substitut* or treat*))).tw,id. 135759
13 randomly.ab. 86984
14 ((single or double or triple or treble) adj2 (blind* or mask* or dummy)).tw,id. 30071
15 trial.ti. 40759
16 (placebo and control* and trial).ab,id. 8886
17 (placebo adj5 (control* or group*)).tw,id. 23667
18 or/10-17 224211
19 9 and 18 1010
20 ((allocat* or assign*) and (group* or control*)).tw,id. and ((cannabis or marijuana) and (abuse or dependency or "substance use disorder" or "substance use treatment")).hw. 85
21 19 or 20 1036
22 ((animal model* or mouse or mice or murine* or rat or rats or rodent* or muridae or murids or rabbit* or leporine* or leporidae or guineapig* or cavies or caviidae or hamster* or cricetidae or gerbil* or gerbillinae or cat or cats or feline* or felidae or dog or dogs or canine* or canidae or pig or pigs or piglet* or minipig* or swine* or porcine* or suidae or horse or horses or donkey or donkies or burros or asses or equine* or equidae or sheep or lamb or lambs or ovine or ovidae or goat or goats or cow or cows or cattle or bovine* or bovidae or primate* or monkey or monkeys or macaque or macaques or marmoset or marmosets) not human*).ti. 182084
23 21 not 22 1033
24 (("literature review" or "systematic review" or meta analysis) not (clinical trial or empirical study or quantitative study or followup study)).md. 222299
25 23 and 24 69
26 23 not 25 964
27 exp psychosocial interventions/ 2210
28 (psychosocial* or psycho-social*).mp. 140248
29 exp psychotherapy/ 227225
30 (psychotherap* or psycho-therap*).mp. 219754
31 (psychological adj3 (therap* or train* or treat* or trial* or intervention*)).mp. 31119
32 exp behavior therapy/ 114739
33 (((behavi* or cognitive) adj3 (therap* or train* or treat* or trial* or intervention*)) or CBT*).mp. 151003
34 behavi* activation.mp. 3552
35 exp biofeedback/ 6983
36 (biofeedback or bio-feedback).mp. 7732
37 exp counseling/ 85712
38 counsel?ing.mp. 119036
39 exp operant conditioning/ 37838

40 (avoidance adj (activit* or learning or train*)).mp. 14356
41 (avoid* adj3 (drug* or cannabi* or marihuana or marijuana or bhang or ganja or ganjah or hashish)).mp. 787
42 motivation/ 65514
43 motivational interviewing/ 3214
44 (motivation* adj3 (interview* or enhanc* or therap* or treat* or trial* or intervention*)).mp. 13265
45 exp behavior modification/ 27653
46 exp behavior change/ 16564
47 (behavio* adj3 (chang* or modif*)).mp. 83572
48 readiness to change/ 1878
49 mindfulness/ 13493
50 (mindful?ness* or (mind adj3 train*)).mp. 22867
51 exp reinforcement/ 59486
52 exp rewards/ 21330
53 exp contingency management/ 3480
54 (contingen* adj (manag* or reinforc*)).mp. 4145
55 community reinforcement*.tw,id. 380
56 Family Therapy/ or Family Intervention/ 27274
57 social support/46216
58 ((family or families or social) adj3 (support* or therap* or intervention*)).mp. 183539
59 (adaptive adj3 (treatment* or intervention*)).mp. 846
60 ((monetary or voucher* or prize*) adj5 (incentiv* or contingen* or reinforc* or reward* or treat* or trial* or intervention*)).mp. 6636
61 (vouchers or prizes or rewards).mp. 36528
62 (incentive* or reward* or reinforcement* or motivation* or contingent*).ti,id. 126718
63 brief interventions/ 458
64 (brief adj3 (therap* or train* or treat* or trial* or intervention*)).mp. 16812
65 exp "substance use treatment"/ or addiction treatment/ 34900
66 exp Intervention/ 148956
67 intervention.ti. or ((cannabi* or marihuana or marijuana or bhang or ganja or ganjah or hashish or addict* or depend* or use* disorder*) adj5 intervention).mp. 63036
68 or/27-67 1169243
69 26 and 68 507

**Central Register of Controlled Trials (CENTRAL) on the Cochrane Library** (Issue 6, 2024; searched 12-June-2024)

ID Search Hits
#1 MeSH descriptor: [Marijuana Abuse] this term only 849
#2 MeSH descriptor: [Marijuana Smoking] this term only and with qualifier(s): [drug therapy - DT, therapy - TH] 51
#3 ((cannabi* or marihuana or marijuana or bhang or ganja or ganjah or hashish) NEAR/3 ((problem* NEXT use*) or overus* or abus* or misus* or dependen* or addict* or (use* NEXT disorder*) or overdos or craving* or cessation* or detox* or withdraw* or abstin*)):ti,ab,kw 1604
#4 ((cannabi* or marihuana or marijuana or bhang or ganja or ganjah or hashish) NEAR/3 (high* or heavy or freq*) NEAR/3 (use* or overuse* or risk* or smok*)):ti,ab,kw 403
#5 (CUD and (cannabi* or marihuana or marijuana or bhang or ganja or ganjah or hashish)):ti,ab,kw 167

#6 ((substance NEAR/2 disorder*) and (cannabi* or marihuana or marijuana or bhang or ganja or ganjah or hashish)):ti,ab,kw 945
#7 (((relaps* NEAR/2 prevent*) or maintenance) NEAR (cannabi* or marihuana or marijuana or bhang or ganja or ganjah or hashish)):ti,ab,kw 46
#8 #1 or #2 or #3 or #4 or #5 or #6 or #7 2346
#9 (psychosocial* or psycho-social*):ti,ab,kw 22061
#10 MeSH descriptor: [Psychotherapy] explode all trees 35556
#11 (psychotherap* or psycho-therap*):ti,ab,kw 17762
#12 (psychological NEAR (therap* or train* or treat* or trial* or intervention*)):ti,ab,kw 21766
#13 (((behavi* or cognitive) NEAR (therap* or train* or treat* or trial* or intervention*)) or CBT*):ti,ab,kw 93878
#14 (behavi* NEXT activation):ti,ab,kw 1405
#15 MeSH descriptor: [Biofeedback, Psychology] explode all trees 2172
#16 (biofeedback or bio-feedback):ti,ab,kw 4551
#17 MeSH descriptor: [Counseling] explode all trees 7565
#18 (counseling or counselling):ti,ab,kw 27992
#19 MeSH descriptor: [Conditioning, Psychological] explode all trees 1325
#20 MeSH descriptor: [Avoidance Learning] this term only 381
#21 avoidance:ti,ab,kw 8299
#22 (avoid* NEAR (drug* or cannabi* or marihuana or marijuana or bhang or ganja or ganjah or hashish)):ti,ab,kw 894
#23 MeSH descriptor: [Motivation] explode all trees 12337
#24 MeSH descriptor: [Motivational Interviewing] this term only 1354
#25 MeSH descriptor: [Directive Counseling] this term only 472
#26 (motivation* NEAR (interview* or enhanc* or therap* or treat* or trial* or intervention*)):ti,ab,kw 11475
#27 (behavio* NEAR (chang* or modif*)):ti,ab,kw 23055
#28 MeSH descriptor: [Mindfulness] explode all trees 2314
#29 (mindfulness* or mindfullness* or (mind NEAR train*)):ti,ab,kw 10050
#30 MeSH descriptor: [Reinforcement, Psychology] explode all trees 3079
#31 MeSH descriptor: [Reward] explode all trees 1490
#32 (contingen* NEXT (manag* or reinforc*)):ti,ab,kw 1088
#33 community reinforcement*:ti,ab,kw 573
#34 ((family or families or social) NEAR (support* or therap* or intervention*)):ti,ab,kw 31868
#35 (adaptive AND (treatment* or intervention*)):ti,ab,kw 8137
#36 ((monetary or voucher* or prize*) AND (incentiv* or contingen* or reinforc* or reward* or treat* or trial* or intervention*)):ti,ab,kw 3419
#37 (vouchers or prizes or rewards):ti,ab,kw 2599
#38 (incentiv* or reward* or reinforce* or motivat* or contingent*):ti,kw 21427
#39 (brief NEAR (therap* or train* or treat* or trial* or intervention*)):ti,ab,kw 12911
#40 MeSH descriptor: [Marijuana Abuse] explode all trees and with qualifier(s): [therapy - TH] 160
#41 MeSH descriptor: [Marijuana Smoking] explode all trees and with qualifier(s): [therapy - TH] 31
#42 MeSH descriptor: [Substance Withdrawal Syndrome] explode all trees and with qualifier(s): [therapy - TH] 120
#43 intervention*:ti 85301
#44 ((cannabi* or marihuana or marijuana or bhang or ganja or ganjah or hashish or addict* or depend* or use* disorder*) NEAR intervention*):ti,ab,kw 42639

#45 #9 OR #10 OR #11 OR #12 OR #13 OR #14 OR #15 OR #16 OR #17 OR #18 OR #19 OR #20 OR #21 OR #22 OR #23 OR #24 OR #25 OR #26 OR #27 OR #28 OR #29 OR #30 OR #31 OR #32 OR #33 OR #34 OR #35 OR #36 OR #37 OR #38 OR #39 OR #40 OR #41 OR #42 OR #43 OR #44 289344
#46 #8 AND #45 1412
[Trials, n=1397]
#47 #8 NOT #46 934
[Trials, n=931]

#### Economic search strategies

**Ovid MEDLINE(R) ALL** <1946 to July 30, 2024>
1 Marijuana Abuse/ 7240
2 Marijuana Smoking/dt, th 100
3 ((cannabi* or marihuana or marijuana or bhang or ganja or ganjah or hashish) adj3 (problem* use* or overus* or abus* or misus* or dependen* or addict* or use* disorder* or overdos or craving* or cessation* or detox* or withdraw* or abstin*)).tw,kf. 6284
4 ((cannabi* or marihuana or marijuana or bhang or ganja or ganjah or hashish) adj3 (high* or heavy or freq*) adj3 (use* or overuse* or risk* or smok*)).tw,kf. 3354
5 (CUD and (cannabi* or marihuana or marijuana or bhang or ganja or ganjah or hashish)).tw,kf.731
6 ((substance adj2 disorder*) and (cannabi* or marihuana or marijuana or bhang or ganja or ganjah or hashish)).ti,kf,hw. 6936
7 (((relaps* adj1 prevent*) or maintenance) adj5 (cannabi* or marihuana or marijuana or bhang or ganja or ganjah or hashish)).tw,kf. 94
8 or/1-7 17367
9 Psychosocial Intervention/ 1194
10 (psychosocial* or psycho-social*).tw,kf. 135407
11 exp Psychotherapy/ 225117
12 (psychotherap* or psycho-therap*).tw,kf. 56765
13 (psychological adj3 (therap* or train* or treat* or trial* or intervention*)).tw,kf. 27112
14 (((behavi* or cognitive) adj3 (therap* or train* or treat* or trial* or intervention*)) or CBT*).tw,kf. 121488
15 behavi* activation.tw,kf. 2690
16 exp Biofeedback, Psychology/ 13410
17 (biofeedback or bio-feedback).tw,kf. 8797
18 exp Counseling/ 50061
19 counsel?ing.tw,kf. 126120
20 exp conditioning, psychological/ or avoidance learning/ 75393
21 (avoidance adj (activit* or learning or train*)).tw,kf. 3279
22 (avoid* adj3 (drug* or cannabi* or marihuana or marijuana or bhang or ganja or ganjah or hashish)).tw,kf. 4490
23 *Motivation/ 34236
24 directive counseling/ or motivational interviewing/ 5132
25 (motivation* adj3 (interview* or enhanc* or therap* or treat* or trial* or intervention*)).tw,kf. 12690
26 (behavio* adj3 (chang* or modif*)).tw,kf. 104473
27 Mindfulness/ 7102
28 (mindful?ness* or (mind adj3 train*)).tw,kf. 15099
29 exp reinforcement, psychology/ or exp reward/ 63424

30 (contingen* adj (manag* or reinforc*)).tw,kf. 1716
31 community reinforcement*.tw,kf. 234
32 exp Psychotherapy, Group/ 28087
33 ((family or families or social) adj3 (support* or therap* or intervention*)).tw,kf. 116550
34 (adaptive adj3 (treatment* or intervention*)).tw,kf. 2204
35 ((monetary or voucher* or prize*) adj5 (incentiv* or contingen* or reinforc* or reward* or treat* or trial* or intervention*)).tw,kf. 4727
36 (vouchers or prizes or rewards).tw,kf. 18741
37 (incentive* or reward* or reinforcement* or motivation* or contingent*).ti,kf. 75596
38 (brief adj3 (therap* or train* or treat* or trial* or intervention*)).tw,kf. 17138
39 Marijuana Abuse/th or Marijuana Smoking/th 568
40 Substance Withdrawal Syndrome/th 1009
41 intervention.ti. or ((cannabi* or marihuana or marijuana or bhang or ganja or ganjah or hashish or addict* or depend* or use* disorder*) adj5 intervention*).ab,kf. 134558
42 ((acceptance adj2 commitment therap*) or dialectical behavio* therap*).mp. 3410
43 (psychoeducat* or psycho-educat* or problem sol*).mp. 53845
44 or/9-43 1097376
45 8 and 44 3035
46 *Economics/ 10816
47 Value of life/ 5828
48 exp "costs and cost analysis"/ 272007
49 exp economics, medical/ 14440
50 exp "fees and charges"/ 31482
51 exp budgets/ 14234
52 budget*.tw,kf. 38439
53 intervention costs.tw,kf. 931
54 (cost? per adj2 (adolescent or adult or man or woman or male or female)).tw,kf. 214
55 economic*.ti. 62835
56 (cost* adj2 (effective* or utilit* or benefit* or minimi* or unit* or estimat* or variable*)).tw,kf. 241478
57 (value adj2 (money or monetary)).tw,kf. 3243
58 or/46-57 550119
59 exp Health Care Costs/ 73393
60 "Cost of Illness"/ 32712
61 Health Expenditures/ 24890
62 (cost? adj2 (illness or disease or sickness or health care or healthcare or treatment or direct or indirect or medical or resource)).tw,kf. 94847
63 (burden? adj2 economic*).tw,kf. 20068
64 (utili?ation adj2 (health or medical or resource)).tw,kf.38579
65 (out-of-pocket adj2 (payment? or expenditure? or cost? or spending or expense?)).tw,kf. 7665
66 (expenditure? adj3 (health or direct or indirect)).tw,kf. 12207
67 (healthcare cost* or health care cost* or healthcare utili?ation or health care utili?ation or cost of illness).tw,kf. 57334
68 (cost* adj2 (analy* or outcome or outcomes)).tw,kf. 56842
69 or/59-68 277953
70 quality-adjusted life years/ 16622
71 qaly*.tw,kf. 15700
72 quality adjusted life year*.tw,kf. 18466

73 (eq-5d or eq5d or eq-5 or eq5 or euroqual or euro qual or euro qual5d or euroqual5d or euro qol or euroqol or euro qol5d or euroqol5d or euro quol or euroquol or euro quol5d or euroquol5d or eur qol or eurqol or eur qol5d or eur qol5d or eur?qul or eur?qul5d or euro* quality of life or european qol).ti,ab,kf. 19310
74 (euro* adj3 (5 d or 5d or 5 dimension* or 5dimension* or 5 domain* or 5domain*)).tw,kf. 6604
75 ((hql* or hqol* or h qol* or hrqol* or hr qol* or quality of life) adj2 (increase* or decrease* or improv* or declin* or reduc* or high* or low* or effect or effects or worse or score or scores or change? or impact? or impacted or deteriorate*)).tw,kf. 130675
76 *"quality of life"/ and economics.fs. 2946
77 "quality of life"/ and ((quality or qol) adj3 (improv* or chang*)).tw,kf. 46773
78 (health utility* or utility score* or disutilit*).tw,kf. 4071
79 (utilities or (utilit$ adj3 (score? or value* or health* or cost* or analys* or measur* or disease* or mean or gain or gains or index or indices))).tw,kf. 32289
80 (hui or hui1 or hui-1 or hui2 or hui-2 or hui3 or hui-3).tw,kf. 2121
81 health* year* equivalent*.tw,kf. 40
82 (willingness to pay or time tradeoff or time trade off or tto or standard gamble*).tw,kf. 12543
83 (sf36* or sf-36* or sf 36 or sf6 or sf 6 or sf-6 or sf6d or sf 6d or sf-6d or sf8 or sf-8 or sf 8 or sf12 or sf-12 or sf 12 or sf16 or sf-16 or sf 16 or sf20 or sf-20 or sf 20 or sf thirtysix or sf thirty six).tw,kf. 38549
84 (visual analog* scale* or EQ-VAS).tw,kf. 80643
85 or/70-84 309052
86 exp models, economic/ 16429
87 (markov* or monte carlo).tw,kf. 92313
88 econom* model*.tw,kf. 6332
89 ((value adj2 information analysis) or (expected value adj3 perfect information) or (expected value adj3 sampl* information)).tw,kf. 484
90 (microsimulation? or micro-simulation?).tw,kf. 2331
91 discrete event? simulation?.tw,kf. 1056
92 discrete choice experiment*.tw,kf. 3252
93 or/86-92 116560
94 45 and 58 71
95 45 and 69 45
96 45 and 85 34
97 45 and 93 3
98 94 or 95 or 96 or 97 114

**Ovid Embase** <1974 to 2024 July 30>
1 cannabis addiction/ 12284
2 *cannabis smoking/ 1662
3 ((cannabi* or marihuana or marijuana or bhang or ganja or ganjah or hashish) adj3 (problem* use* or overus* or abus* or misus* or dependen* or addict* or use* disorder* or overdos or craving* or cessation* or detox* or withdraw* or abstin*)).tw,kf. 8928
4 ((cannabi* or marihuana or marijuana or bhang or ganja or ganjah or hashish) adj3 (high* or heavy or freq*) adj3 (use* or overuse* or risk* or smok*)).tw,kf. 4478
5 (CUD and (cannabi* or marihuana or marijuana or bhang or ganja or ganjah or hashish)).tw,kf.996
6 ((substance adj2 disorder*) and (cannabi* or marihuana or marijuana or bhang or ganja or ganjah or hashish)).ti,kf,hw. 2700

7 (((relaps* adj1 prevent*) or maintenance) adj5 (cannabi* or marihuana or marijuana or bhang or ganja or ganjah or hashish)).tw,kf. 118
8 or/1-7 22326
9 psychosocial intervention/ 2818
10 (psychosocial* or psycho-social*).tw,kf. 185802
11 exp psychotherapy/ 311693
12 (psychotherap* or psycho-therap*).tw,kf. 74720
13 (psychological adj3 (therap* or train* or treat* or trial* or intervention*)).tw,kf. 37940
14 (((behavi* or cognitive) adj3 (therap* or train* or treat* or trial* or intervention*)) or CBT*).tw,kf. 166615
15 behavi* activation.tw,kf. 3309
16 exp biofeedback/ 8865
17 (biofeedback or bio-feedback).tw,kf. 13164
18 exp counseling/ 212280
19 counsel?ing.tw,kf. 184901
20 exp "conditioning (psychology)"/ 2738
21 exp avoidance behavior/ 48060
22 (avoidance adj (activit* or learning or train*)).tw,kf. 3254
23 (avoid* adj3 (drug* or cannabi* or marihuana or marijuana or bhang or ganja or ganjah or hashish)).tw,kf. 6854
24 *motivation/ or incentive/ 38121
25 motivational interviewing/ 7538
26 (motivation* adj3 (interview* or enhanc* or therap* or treat* or trial* or intervention*)).tw,kf. 17437
27 (behavio* adj3 (chang* or modif*)).tw,kf. 129981
28 mindfulness/ or mindfulness meditation/ or mindfulness-based stress reduction/ or mindfulness-based cognitive therapy/ 18498
29 "reinforcement (psychology)"/ 5129
30 *reward/ or monetary reward/ 15444
31 (contingen* adj (manag* or reinforc*)).tw,kf. 2119
32 (contingence management or motivational enhancement therapy).dq. 240
33 community reinforcement*.tw,kf. 291
34 group therapy/ or exp family therapy/ 35024
35 ((family or families or social) adj3 (support* or therap* or intervention*)).tw,kf. 148553
36 (adaptive adj3 (treatment* or intervention*)).tw,kf. 3251
37 ((monetary or voucher* or prize*) adj5 (incentiv* or contingen* or reinforc* or reward* or treat* or trial* or intervention*)).tw,kf. 6122
38 voucher program/ 99
39 (vouchers or prizes or rewards).tw,kf. 21985
40 (incentive* or reward* or reinforcement* or motivation* or contingent*).ti,kf. 85342
41 intervention.ti. or ((cannabi* or marihuana or marijuana or bhang or ganja or ganjah or hashish or addict* or depend* or use* disorder*) adj5 intervention*).ab,kf. 179695
42 (mindful?ness* or (mind adj3 train*)).tw,kf. 18820
43 (brief adj3 (therap* or train* or treat* or trial* or intervention*)).tw,kf. 22350
44 ((acceptance adj2 commitment therap*) or dialectical behavio* therap*).mp. 5818
45 (psychoeducat* or psycho-educat* or problem sol*).mp. 74380
46 or/9-45 1417561
47 8 and 46 4241

48 *economics/ 27998
49 health economics/ 36699
50 exp economic evaluation/ 372502
51 "cost"/ 64837
52 exp fee/ 45491
53 budget/ 35031
54 *finance/ 4149
55 budget*.tw,kf. 50829
56 intervention costs.tw,kf. 1149
57 (cost? per adj2 (adolescent or adult or man or woman or male or female)).tw,kf. 271
58 (cost* adj2 (effective* or utilit* or benefit* or minimi* or unit* or estimat* or variable*)).tw,kf. 329813
59 economic*.ti. 77271
60 (value adj2 (money or monetary)).tw,kf. 4375
61 or/48-60 765125
62 *"health care cost"/ or health care financing/ 64373
63 "cost of illness"/ 21924
64 "cost benefit analysis"/ 97705
65 (cost? adj2 (illness or disease or sickness or health care or healthcare or treatment or direct or indirect or medical or resource)).tw,kf. 150863
66 (burden? adj2 economic*).tw,kf. 32092
67 (utili#ation adj2 (health or medical or resource)).tw,kf. 59923
68 (out-of-pocket adj2 (payment? or expenditure? or cost? or spending or expense?)).tw,kf. 10930
69 (expenditure? adj3 (health or direct or indirect)).tw,kf. 16027
70 (healthcare cost* or health care cost* or healthcare utili?ation or health care utili?ation or cost of illness).tw,kf. 90958
71 (cost* adj2 (analy* or outcome or outcomes)).tw,kf. 88109
72 or/62-71 420490
73 quality adjusted life year/ 38132
74 (quality adjusted life year? or qaly* or qald* or qale* or qtime*).tw,kf. 35637
75 (eq-5d or eq5d or eq-5 or eq5 or euroqual or euro qual or euro qual5d or euroqual5d or euro qol or euroqol or euro qol5d or euroqol5d or euro quol or euroquol or euro quol5d or euroquol5d or eur qol or eurqol or eur qol5d or eur qol5d or eur?qul or eur?qul5d or euro* quality of life or european qol).tw,kf. 34287
76 (euro* adj3 (5 d or 5d or 5 dimension* or 5dimension* or 5 domain* or 5domain*)).tw,kf. 9836
77 ((hql* or hqol* or h qol* or hrqol* or hr qol*) adj2 (increase* or decrease* or improv* or declin* or reduc* or high* or low* or effect or effects or worse or score or scores or change? or impact? or impacted or deteriorate*)).tw,kf. 20421
78 "quality of life"/ and economics/ 3589
79 (multiattribute* or multi attribute*).tw,kf. 1698
80 health utilit*.tw,kf. 4970
81 (utilit* adj2 (value* or cost* or health or analys* or index or indices)).tw,kf. 25861
82 disutilit$.tw,kf. 1385
83 (hsuv or hsuvs).tw,kf. 226
84 (health? year? equivalent? or hye?).tw,kf. 274
85 (hui or hui1 or hui2 or hui3 or hui-1 or hui-2 or hui-3).tw,kf. 3408
86 (hye or hyes).tw,kf. 200
87 rosser.tw,kf. 147
88 exp short form 36/ and (QoL or "quality of life").mp. 39025

89 (sf36$ or sf-36$ or sf 36 or sf6 or sf 6 or sf-6 or sf6d or sf 6d or sf-6d or sf8 or sf-8 or sf 8 or sf12 or sf-12 or sf 12 or sf16 or sf-16 or sf 16 or sf20 or sf-20 or sf 20 or sf thirtysix or sf thirty six).tw,kf. and (QoL or "quality of life").mp. 45288
90 (15d or 15-d or 15 dimension).tw,kf. 8032
91 standard gamble/ or time trade-off method/ or willingness to pay/ 5216
92 standard gamble*.tw,kf. 1242
93 ("time trade off?" or time tradeoff? or tto or timetradeoff?).tw,kf. 3770
94 "willingness to pay".tw,kf. 14467
95 EQ-VAS.tw,kf. 2800
96 or/73-95 173814
97 economic model/ or econometric model/ 4306
98 monte carlo method/ or markov chain monte carlo method/ 56196
99 econom* model*.tw,kf. 8942
100 (markov* or monte carlo).tw,kf. 104593
101 ((value adj2 information analysis) or (expected value adj3 perfect information) or (expected value adj3 sampl* information)).tw,kf. 689
102 (microsimulation? or micro-simulation?).tw,kf. 3548
103 discrete event? simulation?.tw,kf. 1611
104 discrete choice experiment?.tw,kf. 4724
105 or/97-104 135178
106 47 and 61 107
107 47 and 72 72
108 47 and 96 23
109 47 and 105 4
110 or/106-109 152
