## Supplementary material for "Effectiveness and safety of psychosocial interventions for the treatment of cannabis use disorder: a systematic review and meta-analysis": SuppInfo04

### SUPPORTING INFORMATION 4. DATA PROCESSING

To prepare the extracted data for quantitative synthesis, the following data processing took place. For dichotomous outcomes, when data were reported as a proportion or percentage of participants with event, we estimated the number of participants with event using the denominator specified by the authors as the number analysed. For continuous outcomes, where the number analysed was unclear, we conservatively assumed that the number of participants with available outcome data was used in the analysis. Missing standard deviations (SDs) were derived from other within-study statistics (e.g. standard errors, SEs) where possible, otherwise they were imputed based on the available means using a linear regression (having confirmed that there was a strong relationship between means and SDs).

For multi-arm trials including two or more of the same intervention types (e.g. two similar variants of cognitive behavioural therapy), we pooled the outcome data into a single arm for analysis. This was to avoid double-counting participants and including multiple correlated comparisons in analyses. Table 3 in the main article indicates which study arms were pooled for synthesis.
