## Supplementary material for "Effectiveness and safety of psychosocial interventions for the treatment of cannabis use disorder: a systematic review and meta-analysis": SuppInfo05

### SUPPORTING INFORMATION 5. ADDITIONAL COMPARISONS

The following pairwise comparisons were prioritized for meta-analyses and reported in the main article: (1) cognitive-behavioural therapy (CBT) versus inactive/nonspecific comparators; (2) dialectical behaviour/acceptance and commitment therapies (DBT/ACT) versus inactive/nonspecific comparators; (3) CBT plus affect management (CBT-affect) versus standard CBT; (4) CBT plus abstinence-based contingency management (CM-abstinence) versus CBT alone; (5) CBT plus CM-abstinence versus CBT plus attendance-based CM (CM-attendance); (6) multidimensional family therapy (MDFT) versus CBT; and (7) community reinforcement versus other active/nonspecific comparators. To reduce multiplicity of analyses (e.g. by reusing the same intervention arms across different comparisons), other potential comparisons were not prioritised for synthesis. Study-level effect estimates for these comparisons are reported in Table 1. Numbering of tables is specific to this Supporting Information document. References relating to this Supporting Information are at the end of this document.

**Table 1.** Study-level effect estimates for additional comparisons not included in the meta-analyses, for outcomes assessed at the end of treatment

| Comparison | Study | Study arms | Relative effect estimate [95% CI] |
|---|---|---|---|
| CM-abstinence vs Inactive/nonspecific | Kadden 2007[1] | CM-ab vs NS | Continuous abstinence: OR 2.12 [0.76; 5.91]<br>Completion of treatment: OR 1.85 [0.53; 6.53]<br>Frequency of cannabis use: RoM 0.55 [0.40; 0.76]<br>Duration of continuous abstinence: RoM 1.63 [1.02; 2.62] |
| | Carroll 2006[2] | CM-ab/at vs NS | Completion of treatment: OR 0.67 [0.10; 4.27]<br>Frequency of cannabis use: RoM 1.00 [0.71; 1.41]<br>Duration of continuous abstinence: RoM 2.25 [0.86; 5.89] |
| CBT+CM-abstinence vs CM-abstinence | Budney 2006[3] | MET/CBT/CM-ab vs CM-ab | Continuous abstinence: OR 1.00 [0.33; 3.03]<br>Point abstinence: OR 1.00 [0.33; 3.03]<br>Completion of treatment: OR 1.63 [0.41; 6.47]<br>Frequency of cannabis use: RoM 0.86 [0.54; 1.35] |
| | Carroll 2006[2] | MET/CBT/CM-ab-at vs CM-ab-at | Completion of treatment: OR 1.50 [0.23; 9.61]<br>Frequency of cannabis use: RoM 0.71 [0.48; 1.06]<br>Duration of continuous abstinence: RoM 1.22 [0.71; 2.10] |
| | Carroll 2012[4] | MET/CBT/CM-ab vs CM-ab | Completion of treatment: OR 0.43 [0.08; 2.43]<br>Frequency of cannabis use: RoM 1.55 [0.92; 2.60]<br>Duration of continuous abstinence: RoM 0.85 [0.52; 1.41] |
| | Kadden 2007[1] | MET/CBT/CM-ab vs CM-ab | Continuous abstinence: OR 0.81 [0.33; 2.00]<br>Completion of treatment: OR 1.18 [0.28; 4.96]<br>Frequency of cannabis use: RoM 1.19 [0.85; 1.67]<br>Duration of continuous abstinence: RoM 0.91 [0.60; 1.39] |
| CBT vs CM-abstinence | Carroll 2006[2] | MET/CBT vs CM-ab-at | Completion of treatment: OR 0.60 [0.13; 2.73]<br>Frequency of cannabis use: RoM 1.00 [0.71; 1.41]<br>Duration of continuous abstinence: RoM 0.72 [0.36; 1.43] |
| | Carroll 2012[4] | MET/CBT vs CM-ab | Completion of treatment: OR 0.14 [0.03; 0.70]<br>Frequency of cannabis use: RoM 1.12 [0.64; 1.95] |

| Comparison | Study | Study arms | Relative effect estimate [95% CI] |
|---|---|---|---|
| | | | Duration of continuous abstinence: RoM 1.06 [0.69; 1.64] |
| | Kadden 2007[1] | MET/CBT vs CM-ab | Continuous abstinence: OR 0.54 [0.20; 1.45]<br>Completion of treatment: OR 0.73 [0.20; 2.75]<br>Frequency of cannabis use: RoM 1.48 [1.06; 2.06]<br>Duration of continuous abstinence: RoM 0.68 [0.43; 1.08] |
| CBT+CM-attendance vs CBT | Carroll 2012[4] | MET/CBT/CM-at vs MET/CBT | Completion of treatment: OR 2.45 [0.80; 7.49]<br>Frequency of cannabis use: RoM 0.97 [0.61; 1.53]<br>Duration of continuous abstinence: RoM 0.95 [0.66; 1.36] |

CBT, cognitive-behavioural therapy; CI, confidence interval; CM, contingency management; CM-ab, contingency management based on abstinence; CM-at, contingency management based on attendance; CM-ab-at, contingency management based on abstinence and attendance; MET, motivation enhancement therapy; NS, nonspecific treatment; OR, odds ratio; RoM, ratio of means.
