## Supplementary material for "Effectiveness and safety of psychosocial interventions for the treatment of cannabis use disorder: a systematic review and meta-analysis": SuppInfo06

### SUPPORTING INFORMATION 6. CERTAINTY OF EVIDENCE (GRADE) CRITERIA

To assess the certainty of evidence, we followed the GRADE approach.[1] We outline the domains we considered and the steps taken to assess certainty within each domain. References relating to this Supporting Information are included at the end of this document.

*Study limitations (risk of bias)*

We assessed risk of bias using the Risk of Bias 2 (RoB 2)[2] tool for each study that contributed data for a specific outcome. If we had concerns regarding the risk of bias in the studies included in an analysis, the certainty of the evidence was reduced by either one or two levels, depending on the strength of concern, the number (and weight) of the studies affected.

*Inconsistency (heterogeneity)*

We assessed inconsistency using visual inspection of the forest plots (assessing the overlap in confidence intervals, and the direction and magnitude of effects in the individual studies) and the $I^2$ value. If a single study was included, we did not reduce the certainty of the evidence for inconsistency.

*Indirectness*

We considered the population, interventions, comparators and outcomes (PICO) included in every trial contributing to a specific outcome. If the PICO for individual studies differed slightly from that assessed in the review we considered the weight of these studies in the analysis. If studies with concerns over indirectness had a substantial impact on the analysis results we reduced the certainty of the evidence. Examples included recruitment of some adolescents aged below 16 years, or inclusion of specific populations of people with cannabis use disorder (CUD) (for example, only those with insomnia or anxiety disorders).

*Imprecision*

There are no established thresholds representing minimally important differences for the outcomes considered in this review. Therefore, we applied thresholds for what could constitute a clinically meaningful change, that had been derived in consultation with topic experts.

For dichotomous outcomes (point abstinence, continuous abstinence, completion of treatment) we assumed an event rate in the control group, and specified an event rate in the intervention group that we considered would represent an important difference between groups. For abstinence outcomes, we assumed a control group rate of 20%, and an intervention group rate of 30%. Sample size calculations indicated that each group would require approximately 293 participants, to detect this difference (with power of 0.8 at the 0.05 level). We therefore used a total sample size of 586 participants for the optimal information size (OIS). For completion of treatment, we assumed a control group rate of 70%, and an intervention group rate of 80%. This required a sample size of 293 participants for each group, therefore we used 588 as the OIS.

If the OIS was not reached then we reduced the certainty of the evidence by one level. If the sample size was extremely small (<100 participants) then we reduced the certainty of the evidence by two levels.

If the OIS was reached then we considered the breadth of the confidence intervals, taking both relative and absolute effects into account. We assumed a change in odds ratio of 25% would represent an important difference, i.e. if the confidence intervals for the odds ratio (OR) crossed the thresholds of either 0.8 or 1.25 we considered reducing the certainty of the evidence due to imprecision by one level. If both thresholds were crossed we considered reducing the certainty of the evidence by two levels. However, we also considered the change in absolute risk before applying these rules. Where large changes in relative risk (as assessed by confidence intervals surrounding the OR) did not equate to substantial changes in absolute risk, we did not reduce the certainty of the evidence.

For continuous data, we also estimated an OIS using assumed control group rates. For duration of continuous abstinence, we assumed a control group rate of 7 days, an intervention group rate of 14 days, and a standard deviation (SD) of 14 days (note the large SD in the data reported). This gave an estimated total sample size of 126 participants, which was used for the OIS. Similarly, for frequency of use we assumed a control group rate of 0.5 (proportion of days using), an intervention rate of 0.25, and a SD of 0.3, which gave an estimated total sample size of 46 participants. For quantity of cannabis use we assumed use of 2 joints per day in the control group, and 1 joint per day in the intervention group, with a SD of 2, which gave an estimated total sample size of 126 participants, which was used for the OIS.

Again, if the OIS was not reached then the certainty of the evidence was reduced by one level. If the OIS was reached then we considered the breadth of the confidence intervals. For continuous outcomes, we assumed that halving of either the quantity or frequency of use would represent an important difference, i.e. if the confidence intervals for the ratio of means (RoM) crossed the thresholds of either 0.5 or 2 we considered reducing the certainty of the evidence due to imprecision by one level. If both thresholds were crossed we considered reducing the certainty of the evidence by two levels.

Only one study reported on cannabis craving, using the Marijuana Craving Questionnaire short-form.[3] This questionnaire has 12 domains, each rated with a Likert scale on a score of 1-7, giving a total range of scores from 12-84, with higher scores representing worse cravings. We are not aware of an agreed minimally important difference for this questionnaire, therefore assumed that a change of 4 points would be clinically meaningful (the reported standard deviation was approximately 8 points, so this magnitude of change represents approximately half of the sample standard deviation). The required sample size for power of 0.8 at the 0.05 significance level was estimated at 126 participants.

*Publication bias*

We also considered the risk of publication bias in the results presented. Our searches were comprehensive, which reduces the likelihood that studies were omitted from the review. However, we did note that many studies did not report data for all outcomes of interest in this review. This may simply be because these outcomes were not assessed, but also raises the possibility of selective reporting, where authors have deliberately withheld data from publication. Most of the studies included did not report full trial registrations or protocols, and we were therefore unable to assess the trialists intentions. In addition, the trial registrations that were available did not typically report in sufficient detail to be able to establish whether all outcomes were fully reported. Consequently, although we did have some concerns regarding the risk of publication bias, we did not consider this sufficient justification to lower the certainty of evidence further.
