## Supplementary material for "Effectiveness and safety of psychosocial interventions for the treatment of cannabis use disorder: a systematic review and meta-analysis": SuppInfo07

### SUPPORTING INFORMATION 7. EXCLUDED STUDIES

Numbering of tables is specific to this Supporting Information document.

**Table 1.** List of excluded reports

| Reference | Reason for exclusion |
|---|---|
| Abroms LC, Fishman M, Vo H, Chiang SC, Somerville V, Rakhmanov L*, et al.* A Motion-Activated Video Game for Prevention of Substance Use Disorder Relapse in Youth: Pilot Randomized Controlled Trial. *JMIR serious games* 2019;**7**:e11716. https://doi.org/10.2196/11716 | Ineligible population |
| ACTRN. A randomised controlled trial of a novel combination pharmacotherapy program for the treatment of cannabis dependence. *https://trialsearchwhoint/Trial2aspx?TrialID=ACTRN12609000581268* 2009. | Ineligible publication type |
| ACTRN. Double blind, randomised, placebo controlled trial of lithium carbonate for the management of cannabis withdrawal in adult humans. *http://wwwwhoint/trialsearch/Trial2aspx?TrialID=ACTRN12610000182099* 2010. | Ineligible publication type |
| ACTRN. Intranasal oxytocin for the treatment of cannabis and alcohol dependence. *http://wwwwhoint/trialsearch/Trial2aspx?TrialID=ACTRN12610000742077* 2010. | Ineligible publication type |
| ACTRN. Feasibility Study of a Brief Telephone-based Cannabis Intervention. *http://wwwwhoint/trialsearch/Trial2aspx?TrialID=ACTRN12615001174572* 2015. | Ineligible publication type |
| ACTRN. Cannabidiol (CBD) for Cannabis and Mood Disorders in Adolescence (CCAMDA). *http://wwwwhoint/trialsearch/Trial2aspx?TrialID=ACTRN12616001001482* 2016. | Ineligible publication type |
| ACTRN. Pilot Study: effect of electrical brain stimulation on mental skills of individuals with long-term cannabis use. *https://trialsearchwhoint/Trial2aspx?TrialID=ACTRN12619000462189* 2019. | Ineligible publication type |
| Adams TR, Arnsten JH, Ning Y, Nahvi S. Feasibility and Preliminary Effectiveness of Varenicline for Treating Co-Occurring Cannabis and Tobacco Use. *Journal of psychoactive drugs* 2018;**50**:12-8. https://doi.org/10.1080/02791072.2017.1370746 | Ineligible population |
| Aharonovich E, Campbell ANC, Shulman M, Hu MC, Kyle T, Winhusen T*, et al.* Neurocognitive profiling of adult treatment seekers enrolled in a clinical trial of a web-delivered intervention for substance use disorders. *Journal of addiction medicine* 2018;**12**:99-106. https://doi.org/10.1097/ADM.0000000000000372 | Ineligible study design |
| Aharonovich E, Greenstein E, O'Leary A, Johnston B, Seol SG, Hasin DS. HealthCall: technology-based extension of motivational interviewing to reduce non-injection drug use in HIV primary care patients - a pilot study. *AIDS care* 2012;**24**:1461-9. https://doi.org/10.1080/09540121.2012.663882 | Ineligible population |
| Ahlers J, Baumgartner C, Augsburger M, Wenger A, Malischnig D, Boumparis N*, et al.* Cannabis Use in Adults Who Screen Positive for Attention Deficit/Hyperactivity Disorder: CANreduce 2.0 Randomized Controlled Trial Subgroup Analysis. *Journal of medical Internet research* 2022;**24**:e30138. https://doi.org/10.2196/30138 | Ineligible study design |
| Allsop DJ, Copeland J, Lintzeris N, Dunlop AJ, Montebello M, Sadler C*, et al.* Nabiximols as an agonist replacement therapy during cannabis withdrawal: a randomized clinical trial. *JAMA Psychiatry* 2014;**71**:281-91. https://doi.org/10.1001/jamapsychiatry.2013.3947 | Ineligible intervention |
| Allsop DJ, Rooney K, Arnold JC, Bhardwaj AK, Bruno R, Bartlett DJ*, et al.* Randomised controlled trial (RCT) of daily aerobic exercise for inpatient cannabis withdrawal: a study protocol. *Mental health and physical activity* 2017;**13**:57-67. https://doi.org/10.1016/j.mhpa.2017.06.002 | Ineligible publication type |
| Amann M, Haug S, Wenger A, Baumgartner C, Ebert DD, Berger T*, et al.* The Effects of Social Presence on Adherence-Focused Guidance in Problematic Cannabis Users: protocol for the CANreduce 2.0 Randomized Controlled Trial. *JMIR research protocols* 2018;**7**:e30. https://doi.org/10.2196/resprot.9484 | Ineligible intervention |
| Amaro H, Black DS. Mindfulness-Based Intervention Effects on Substance Use and Relapse Among Women in Residential Treatment: a Randomized Controlled Trial With 8.5-Month Follow-Up Period From the Moment-by-Moment in Women's Recovery Project. *Psychosomatic medicine* 2021;**83**:528-38. https://doi.org/10.1097/PSY.0000000000000907 | Ineligible population |
| Arias AJ, Hammond CJ, Burleson JA, Kaminer Y, Feinn R, Curry JF*, et al.* Temporal dynamics of the relationship between change in depressive symptoms and cannabis use in adolescents receiving psychosocial treatment for cannabis use disorder. *Journal of substance abuse treatment* 2020;**117**:108087. https://doi.org/10.1016/j.jsat.2020.108087 | Ineligible population |
| Arnedt JT, Conroy DA, Stewart H, Yeagley E, Bowyer G, Bohnert KM*, et al.* Cognitive behavioral therapy for insomnia to reduce cannabis use: Results from a pilot randomized controlled trial. *Drug and Alcohol Dependence* 2023;**246**:1-9. https://doi.org/10.1016/j.drugalcdep.2023.109835 | Ineligible population |
| Azhari N, Hu H, O'Malley KY, Blocker ME, Levin FR, Dakwar E. Ketamine-facilitated behavioral treatment for cannabis use disorder: A proof of concept study. *Am J Drug Alcohol Abuse* 2021;**47**:92-7. https://doi.org/10.1080/00952990.2020.1808982 | Ineligible study design |

| | |
|---|---|
| Badour CL, Flanagan JC, Gros DF, Killeen T, Pericot-Valverde I, Korte KJ, *et al.* Habituation of distress and craving during treatment as predictors of change in PTSD symptoms and substance use severity. *Journal of consulting and clinical psychology* 2017;**85**:274-81. https://doi.org/10.1037/ccp0000180 | Ineligible population |
| Bagoien G, Bjorngaard JH, Ostensen C, Reitan SK, Romundstad P, Morken G. The effects of motivational interviewing on patients with comorbid substance use admitted to a psychiatric emergency unit - a randomised controlled trial with two year follow-up. *BMC Psychiatry* 2013;**13**:93. https://doi.org/10.1186/1471-244X-13-93 | Ineligible population |
| Baker A, Lewin T, Reichler H, Clancy R, Carr V, Garrett R, *et al.* Evaluation of a motivational interview for substance use within psychiatric in-patient services. *Addiction* 2002;**97**:1329-37. https://doi.org/10.1046/j.1360-0443.2002.00178.x | Ineligible population |
| Baker A, Turner A, Kay-Lambkin FJ, Lewin TJ. The long and the short of treatments for alcohol or cannabis misuse among people with severe mental disorders. *Addictive behaviors* 2009;**34**:852-8. https://doi.org/10.1016/j.addbeh.2009.02.002 | Ineligible study design |
| Banes KE, Stephens RS, Blevins CE, Walker DD, Roffman RA. Changing motives for use: outcomes from a cognitive-behavioral intervention for marijuana-dependent adults. *Drug and alcohol dependence* 2014;**139**:41-6. https://doi.org/10.1016/j.drugalcdep.2014.02.706 | Ineligible intervention |
| Barrett H, Slesnick N, Brody JL, Turner CW, Peterson TR. Treatment outcomes for adolescent substance abuse at 4- and 7-month assessments. *Journal of Consulting and Clinical Psychology* 2001;**69**:802-13. https://doi.org/10.1037/0022-006X.69.5.802 | Ineligible population |
| Battjes RJ, Gordon MS, O'Grady KE, Kinlock TW, Katz EC, Sears EA. Evaluation of a group-based substance abuse treatment program for adolescents. *Journal of substance abuse treatment* 2004;**27**:123-34. | Ineligible population |
| Baumgartner C, Schaub MP, Wenger A, Malischnig D, Augsburger M, Walter M, *et al.* CANreduce 2.0 Adherence-Focused Guidance for Internet Self-Help Among Cannabis Users: three-Arm Randomized Controlled Trial. *Journal of medical Internet research* 2021;**23**:e27463. https://doi.org/10.2196/27463 | Ineligible population |
| Becker J, Haug S, Sullivan R, Schaub MP. Effectiveness of different Web-based interventions to prepare co-smokers of cigarettes and cannabis for double cessation: a three-arm randomized controlled trial. *Journal of medical Internet research* 2014;**16**:e273. https://doi.org/10.2196/jmir.3246 | Ineligible population |
| Becker SJ, Helseth SA, Janssen T, Kelly LM, Escobar KI, Souza T, *et al.* Parent SMART (Substance Misuse in Adolescents in Residential Treatment): pilot randomized trial of a technology-assisted parenting intervention. *Journal of substance abuse treatment* 2021;**127**:108457. https://doi.org/10.1016/j.jsat.2021.108457 | Ineligible population |
| Beckham JC, Calhoun PS, Hertzberg JS, Budney AJ, Aurora P, Dennis MF, *et al.* A randomized clinical trial of mobile contingency management intervention for cannabis use reduction. *International Journal of Mental Health and Addiction* 2024; https://dx.doi.org/10.1007/s11469-024-01314-z | Ineligible intervention |
| Bentzley JP, Tomko RL, Gray KM. Low Pretreatment Impulsivity and High Medication Adherence Increase the Odds of Abstinence in a Trial of N-Acetylcysteine in Adolescents with Cannabis Use Disorder. *Journal of Substance Abuse Treatment* 2016;**63**:72-7. https://doi.org/10.1016/j.jsat.2015.12.003 | Ineligible outcome |
| Bernstein E, Edwards E, Dorfman D, Heeren T, Bliss C, Bernstein J. Screening and brief intervention to reduce marijuana use among youth and young adults in a pediatric emergency department. *Academic emergency medicine* 2009;**16**:1174-85. https://doi.org/10.1111/j.1553-2712.2009.00490.x | Ineligible population |
| Bhardwaj AK, Allsop DJ, Copeland J, McGregor IS, Dunlop A, Shanahan M, *et al.* Randomised Controlled Trial (RCT) of cannabinoid replacement therapy (Nabiximols) for the management of treatment-resistant cannabis dependent patients: a study protocol. *BMC Psychiatry* 2018;**18**:140. https://doi.org/10.1186/s12888-018-1682-2 | Ineligible intervention* |
| Bhardwaj AK, Mills L, Doyle M, Sahid A, Montebello M, Monds L, *et al.* A phase III multisite randomised controlled trial to compare the efficacy of cannabidiol to placebo in the treatment of cannabis use disorder: the CBD-CUD study protocol. *BMC Psychiatry* 2024;**24**:175. https://doi.org/10.1186/s12888-024-05616-3 | Ineligible publication type |
| Blevins CE, Banes KE, Walker DD, Stephens RS, Roffman RA. The relationship between general causality orientation and treatment outcome among marijuana-dependent adults. *Addictive behaviors* 2016;**53**:196-200. https://doi.org/10.1016/j.addbeh.2015.10.021 | Ineligible study design |
| Blevins CE, Stephens RS, Walker DD, Roffman RA. Situational determinants of use and treatment outcomes in marijuana dependent adults. *Addictive behaviors* 2014;**39**:546-52. https://doi.org/10.1016/j.addbeh.2013.10.031 | Ineligible intervention |
| Blow FC, Walton MA, Bohnert ASB, Ignacio RV, Chermack S, Cunningham RM, *et al.* A randomized controlled trial of brief interventions to reduce drug use among adults in a low-income urban emergency department: the HealthiER You study. *Addiction* 2017;**112**:1395-405. https://doi.org/10.1111/add.13773 | Ineligible population |
| Bonar EE, Cunningham RM, Sweezea EC, Blow FC, Drislane LE, Walton MA. Piloting a brief intervention plus mobile boosters for drug use among emerging adults receiving emergency department care. *Drug and alcohol dependence* 2021;**221**:108625. https://doi.org/10.1016/j.drugalcdep.2021.108625 | Ineligible population |
| Bonar EE, Goldstick JE, Tan CY, Bourque C, Carter PM, Duval ER, *et al.* A remote brief intervention plus social media messaging for cannabis use among emerging adults: A pilot randomized controlled trial in | Ineligible population |

| | |
|---|---|
| emergency department patients. *Addictive Behaviors* 2023;**147**:1-11. https://doi.org/10.1016/j.addbeh.2023.107829 | |
| Borodovsky JT, Sofis MJ, Sherman BJ, Gray KM, Budney AJ. Characterizing cannabis use reduction and change in functioning during treatment: initial steps on the path to new clinical endpoints. *Psychology of addictive behaviors* 2022;**36**:515-25. https://doi.org/10.1037/adb0000817 | Ineligible study design |
| Brellenthin AG, Crombie KM, Hillard CJ, Brown RT, Koltyn KF. Psychological and endocannabinoid responses to aerobic exercise in substance use disorder patients. *Substance Abuse* 2021;**42**:272-83. https://doi.org/10.1080/08897077.2019.1680480 | Ineligible population |
| Brezing CA, Choi CJ, Pavlicova M, Brooks D, Mahony AL, Mariani JJ*, et al.* Abstinence and reduced frequency of use are associated with improvements in quality of life among treatment-seekers with cannabis use disorder. *The American journal on addictions* 2018;**27**:101-7. https://doi.org/10.1111/ajad.12660 | Ineligible study design |
| Brooks AJ, Penn PE. Comparing treatments for dual diagnosis: twelve-step and self-management and recovery training. *American journal of drug and alcohol abuse* 2003;**29**:359-83. https://doi.org/10.1081/ADA-120020519 | Ineligible study design |
| Brown PC, Budney AJ, Thostenson JD, Stanger C. Initiation of abstinence in adolescents treated for marijuana use disorders. *Journal of substance abuse treatment* 2013;**44**:384-90. https://doi.org/10.1016/j.jsat.2012.08.223 | Ineligible study design |
| Brown RA, Abrantes AM, Minami H, Prince MA, Bloom EL, Apodaca TR*, et al.* Motivational Interviewing to Reduce Substance Use in Adolescents with Psychiatric Comorbidity. *Journal of substance abuse treatment* 2015;**59**:20-9. https://doi.org/10.1016/j.jsat.2015.06.016 | Ineligible population |
| Buckner JD, Carroll KM. Effect of anxiety on treatment presentation and outcome: results from the Marijuana Treatment Project. *Psychiatry research* 2010;**178**:493-500. https://doi.org/10.1016/j.psychres.2009.10.010 | Ineligible outcome |
| Buckner JD, Morris PE, Zvolensky MJ. Integrated cognitive-behavioral therapy for comorbid cannabis use and anxiety disorders: the impact of severity of cannabis use. *Experimental and clinical psychopharmacology* 2021;**29**:272-8. https://doi.org/10.1037/pha0000456 | Ineligible outcome[a] |
| Buckner JD, Zvolensky MJ, Lewis EM. On-line personalized feedback intervention for negative affect and cannabis: a pilot randomized controlled trial. *Experimental and clinical psychopharmacology* 2020;**28**:143-9. https://doi.org/10.1037/pha0000304 | Ineligible population |
| Budney AJ, Stanger C, Tilford JM, Scherer EB, Brown PC, Li Z*, et al.* Computer-assisted behavioral therapy and contingency management for cannabis use disorder. *Psychology of addictive behaviors* 2015;**29**:501-11. https://doi.org/10.1037/adb0000078 | Ineligible intervention |
| Burleson JA, Kaminer Y. Self-efficacy as a predictor of treatment outcome in adolescent substance use disorders. *Addictive behaviors* 2005;**30**:1751-64. https://doi.org/10.1016/j.addbeh.2005.07.006 | Ineligible population |
| Burlew AK, Montgomery L, Kosinski AS, Forcehimes AA. Does treatment readiness enhance the response of African American substance users to Motivational Enhancement Therapy? *Psychology of addictive behaviors* 2013;**27**:744-53. https://doi.org/10.1037/a0031274 | Ineligible study design |
| Capone C, Presseau C, Saunders E, Eaton E, Hamblen J, McGovern M. Is Integrated CBT Effective in Reducing PTSD Symptoms and Substance Use in Iraq and Afghanistan Veterans? Results from a Randomized Clinical Trial. *Cognitive therapy and research* 2018;**42**:735-46. https://doi.org/10.1007/s10608-018-9931-8 | Ineligible population |
| Caviness CM, Hagerty CE, Anderson BJ, de Dios MA, Hayaki J, Herman D*, et al.* Self-efficacy and motivation to quit marijuana use among young women. *The American journal on addictions* 2013;**22**:373-80. https://doi.org/10.1111/j.1521-0391.2013.12030.x | Ineligible study design |
| Chambers JE, Brooks AC, Medvin R, Metzger DS, Lauby J, Carpenedo CM*, et al.* Examining multi-session brief intervention for substance use in primary care: research methods of a randomized controlled trial. *Addiction science & clinical practice* 2016;**11**:8. https://doi.org/10.1186/s13722-016-0057-6 | Ineligible publication type |
| Chermack ST, Bonar EE, Goldstick JE, Winters J, Blow FC, Friday S*, et al.* A randomized controlled trial for aggression and substance use involvement among Veterans: impact of combining Motivational Interviewing, Cognitive Behavioral Treatment and telephone-based Continuing Care. *Journal of substance abuse treatment* 2019;**98**:78-88. https://doi.org/10.1016/j.jsat.2019.01.001 | Ineligible population |
| Choi M, Driver MN, Balcke E, Saunders T, Langberg JM, Dick DM. Bridging the gap between genetic epidemiological research and prevention: A randomized control trial of a novel Personalized Feedback Program for alcohol and cannabis use. *Drug and Alcohol Dependence* 2023;**249**:1-5. https://doi.org/10.1016/j.drugalcdep.2023.110818 | Ineligible population |
| Choo EK, Zlotnick C, Strong DR, Squires DD, Tape C, Mello MJ. BSAFER: a Web-based intervention for drug use and intimate partner violence demonstrates feasibility and acceptability among women in the emergency department. *Substance abuse* 2016;**37**:441-9. https://doi.org/10.1080/08897077.2015.1134755 | Ineligible population |

| | |
|---|---|
| Christoff Ade O, Boerngen-Lacerda R. Reducing substance involvement in college students: a three-arm parallel-group randomized controlled trial of a computer-based intervention. *Addictive behaviors* 2015;**45**:164-71. https://doi.org/10.1016/j.addbeh.2015.01.019 | Ineligible population |
| Clair-Michaud M, Martin RA, Stein LA, Bassett S, Lebeau R, Golembeske C. The Impact of Motivational Interviewing on Delinquent Behaviors in Incarcerated Adolescents. *Journal of substance abuse treatment* 2016;**65**:13-9. https://doi.org/10.1016/j.jsat.2015.09.003 | Ineligible population |
| Cochran G, Stitzer M, Campbell AN, Hu MC, Vandrey R, Nunes EV. Web-based treatment for substance use disorders: differential effects by primary substance. *Addictive behaviors* 2015;**45**:191-4. https://doi.org/10.1016/j.addbeh.2015.02.002 | Ineligible population |
| Cochran G, Stitzer M, Nunes EV, Hu MC, Campbell A. Clinically relevant characteristics associated with early treatment drug use versus abstinence. *Addiction science & clinical practice* 2014;**9**:6. https://doi.org/10.1186/1940-0640-9-6 | Ineligible population |
| Compton WM, Pringle B. Services research on adolescent drug treatment. Commentary on "The Cannabis Youth Treatment (CYT) Study: main findings from two randomized trials". *Journal of Substance Abuse Treatment* 2004;**27**:195-6. https://doi.org/10.1016/j.jsat.2004.07.003 | Ineligible publication type |
| Conner BT, Thompson K, Prince MA, Bolts OL, Contreras A, Riggs NR*, et al.* Results of a randomized controlled trial of the cannabis eCHECKUP TO GO personalized normative feedback intervention on reducing cannabis use, cannabis consequences, and descriptive norms. *Journal of substance use and addiction treatment* 2024;**159**:209267. https://doi.org/10.1016/j.josat.2023.209267 | Ineligible population |
| Copeland J, Rooke S, Rodriquez D, Norberg MM, Gibson L. Comparison of brief versus extended personalised feedback in an online intervention for cannabis users: short-term findings of a randomised trial. *Journal of substance abuse treatment* 2017;**76**:43-8. https://doi.org/10.1016/j.jsat.2017.01.009 | Ineligible intervention |
| Cornelius JR, Aizenstein HJ, Chung TA, Douaihy A, Hayes J, Daley D*, et al.* Paradoxical Decrease in Striatal Activation on an fMRI Reward Task Following Treatment in Youth with Co-morbid Cannabis Dependence/Major Depression. *Adv* 2013;**93**:123-30. | Ineligible study design |
| Cornelius JR, Bukstein OG, Douaihy AB, Clark DB, Chung TA, Daley DC*, et al.* Double-blind fluoxetine trial in comorbid MDD-CUD youth and young adults. *Drug and alcohol dependence* 2010;**112**:39-45. https://doi.org/10.1016/j.drugalcdep.2010.05.010 | Ineligible intervention* |
| Cornelius JR, Salloum IM, Ferrell R, Douaihy AB, Hayes J, Kirisci L*, et al.* Treatment trial and long-term follow-up evaluation among co-morbid youth with major depression and a cannabis use disorder. *Advances in psychology research, Vol 93* 2012:109-21. | Ineligible publication type |
| Cornelius JR, Salloum IM, Ferrell R, Douaihy AB, Hayes J, Kirisci L*, et al.* Treatment Trial and Long-Term Follow-up Evaluation among Comorbid Youth with Major Depression and a Cannabis Use Disorder. *International Journal of Medical & Biological Frontiers* 2012;**18**:399-411. | Ineligible intervention* |
| Correia CJ, Benson TA, Carey KB. Decreased substance use following increases in alternative behaviors: A preliminary investigation. *Addictive Behaviors* 2005;**30**:19-27. https://doi.org/10.1016/j.addbeh.2004.04.006 | Ineligible population |
| Curry JF, Kaminer Y, Goldston DB, Chan G, Wells KC, Burke RH*, et al.* Adaptive Treatment for Youth With Substance Use and Depression: early Depression Response and Short-term Outcomes. *Journal of the American Academy of Child and Adolescent Psychiatry* 2022;**61**:508-19. https://doi.org/10.1016/j.jaac.2021.07.807 | Ineligible population |
| D'Amico EJ, Hunter SB, Miles JN, Ewing BA, Osilla KC. A randomized controlled trial of a group motivational interviewing intervention for adolescents with a first time alcohol or drug offense. *Journal of substance abuse treatment* 2013;**45**:400-8. https://doi.org/10.1016/j.jsat.2013.06.005 | Ineligible population |
| D'Amico EJ, Miles JN, Stern SA, Meredith LS. Brief motivational interviewing for teens at risk of substance use consequences: a randomized pilot study in a primary care clinic. *Journal of substance abuse treatment* 2008;**35**:53-61. https://doi.org/10.1016/j.jsat.2007.08.008 | Ineligible population |
| D'Amico EJ, Parast L, Osilla KC, Seelam R, Meredith LS, Shadel WG*, et al.* Understanding Which Teenagers Benefit Most From a Brief Primary Care Substance Use Intervention. *Pediatrics* 2019;**144**. https://doi.org/10.1542/peds.2018-3014 | Ineligible population |
| D'Amico EJ, Parast L, Shadel WG, Meredith LS, Seelam R, Stein BD. Brief motivational interviewing intervention to reduce alcohol and marijuana use for at-risk adolescents in primary care. *Journal of Consulting and Clinical Psychology* 2018;**86**:775-86. https://doi.org/10.1037/ccp0000332 | Ineligible population |
| D'Souza DC, Cortes-Briones J, Creatura G, Bluez G, Thurnauer H, Deaso E*, et al.* Efficacy and safety of a fatty acid amide hydrolase inhibitor (PF-04457845) in the treatment of cannabis withdrawal and dependence in men: a double-blind, placebo-controlled, parallel group, phase 2a single-site randomised controlled trial. *The Lancet Psychiatry* 2019;**6**:35-45. https://doi.org/10.1016/S2215-0366%2818%2930427-9 | Ineligible intervention |
| Dakwar E, Levin FR. Individual mindfulness-based psychotherapy for cannabis or cocaine dependence: a pilot feasibility trial. *American Journal on Addictions* 2013;**22**:521-6. https://doi.org/10.1111/j.1521-0391.2013.12036.x | Ineligible study design |

| | |
|---|---|
| Darharaj M, Roshanpajouh M, Amini M, Shrier LA, Habibi Asgarabad M. The effectiveness of mobile-based ecological momentary motivational enhancement therapy in reducing craving and severity of cannabis use disorder: study protocol for a randomized controlled trial. *Internet interventions* 2023;**34**:100669. https://doi.org/10.1016/j.invent.2023.100669 | Ineligible intervention |
| Das S, Hickman NJ, Prochaska JJ. Treating Smoking in Adults With Co-occurring Acute Psychiatric and Addictive Disorders. *Journal of addiction medicine* 2017;**11**:273-9. https://doi.org/10.1097/ADM.0000000000000320 | Ineligible population |
| Dash GF, Bryan AD, Montanaro E, Feldstein Ewing SW. Long-Term RCT outcomes for adolescent alcohol and cannabis use within a predominantly Hispanic sample. *Journal of research on adolescence* 2023;**33**:1038-47. https://doi.org/10.1111/jora.12856 | Ineligible population |
| Dawson-Rose C, Draughon JE, Cuca Y, Zepf R, Huang E, Cooper BA*, et al*. Changes in Specific Substance Involvement Scores among SBIRT recipients in an HIV primary care setting. *Addiction science & clinical practice* 2017;**12**:34. https://doi.org/10.1186/s13722-017-0101-1 | Ineligible population |
| de Dios MA, Herman DS, Britton WB, Hagerty CE, Anderson BJ, Stein MD. Motivational and mindfulness intervention for young adult female marijuana users. *Journal of substance abuse treatment* 2012;**42**:56-64. https://doi.org/10.1016/j.jsat.2011.08.001 | Ineligible population |
| DeGrace S, Barrett SP, Yakovenko I, Tibbo PG, Romero-Sanchiz P, Carleton RN*, et al*. Effects of Trauma Cue Exposure and Posttraumatic Stress Disorder (PTSD) on Affect and Cannabis Craving in Cannabis Users With Trauma Histories: use of Expressive Writing as an Online Cue-Reactivity Paradigm. *Canadian journal of psychiatry* 2024; 10.1177/07067437241255104:7067437241255104. https://doi.org/10.1177/07067437241255104 | Ineligible intervention |
| DeMarce JM, Stephens RS, Roffman RA. Psychological distress and marijuana use before and after treatment: testing cognitive-behavioral matching hypotheses. *Addictive behaviors* 2005;**30**:1055-9. https://doi.org/10.1016/j.addbeh.2004.09.009 | Ineligible outcome[b] |
| Dennis M, Godley SH, Diamond G, Tims FM, Babor T, Donaldson J*, et al*. The Cannabis Youth Treatment (CYT) Study: main findings from two randomized trials. *Journal of Substance Abuse Treatment* 2004;**27**:197-213. https://doi.org/10.1016/j.jsat.2003.09.005 | Ineligible population |
| Dennis M, Titus JC, Diamond G, Donaldson J, Godley SH, Tims FM*, et al*. The Cannabis Youth Treatment (CYT) experiment: rationale, study design and analysis plans. *Addiction* 2002;**97**:16-34. https://doi.org/10.1046/j.1360-0443.97.s01.2.x | Ineligible publication type |
| DeWorsop D, Creatura G, Bluez G, Thurnauer H, Forselius-Bielen K, Ranganathan M*, et al*. Feasibility and success of cell-phone assisted remote observation of medication adherence (CAROMA) in clinical trials. *Drug and alcohol dependence* 2016;**163**:24-30. https://doi.org/10.1016/j.drugalcdep.2016.02.045 | Ineligible intervention |
| Dey M, Wenger A, Baumgartner C, Herrmann U, Augsburger M, Haug S*, et al*. Comparing a mindfulness- and CBT-based guided self-help Internet- and mobile-based intervention against a waiting list control condition as treatment for adults with frequent cannabis use: a randomized controlled trial of CANreduce 3.0. *BMC Psychiatry* 2022;**22**:215. https://doi.org/10.1186/s12888-022-03802-9 | Ineligible intervention |
| Diamond G, Godley SH, Liddle HA, Sampl S, Webb C, Tims FM*, et al*. Five outpatient treatment models for adolescent marijuana use: a description of the Cannabis Youth Treatment Interventions. *Addiction* 2002;**97**:70-83. https://doi.org/10.1046/j.1360-0443.97.s01.3.x | Ineligible study design |
| Diamond G, Panichelli-Mindel SM, Shera D, Dennis M, Tims F, Ungemack J. Psychiatric syndromes in adolescents with marijuana abuse and dependency in outpatient treatment. *Journal of child & adolescent substance abuse* 2006;**15**:37-54. | Ineligible study design |
| Diamond GS, Liddle HA, Wintersteen MB, Dennis ML, Godley SH, Tims F. Early therapeutic alliance as a predictor of treatment outcome for adolescent cannabis users in outpatient treatment. *The American journal on addictions* 2006;**15**:26-33. https://doi.org/10.1080/10550490601003664 | Ineligible study design |
| Dillon FR, Turner CW, Robbins MS, Szapocznik J. Concordance among biological, interview, and self-report measures of drug use among African American and Hispanic adolescents referred for drug abuse treatment. *Psychology of Addictive Behaviors* 2005;**19**:404-13. https://doi.org/10.1037/0893-164X.19.4.404 | Ineligible population |
| Drislane LE, Waller R, Martz ME, Bonar EE, Walton MA, Chermack ST*, et al*. Therapist and computer-based brief interventions for drug use within a randomized controlled trial: effects on parallel trajectories of alcohol use, cannabis use and anxiety symptoms. *Addiction* 2020;**115**:158-69. https://doi.org/10.1111/add.14781 | Ineligible population |
| DRKS. The effect of trauma informed Hatha Yoga on psychopathological symptoms and quality of life in patients with a borderline personality disorder and a xomorbid substance use disorder. *https://trialsearchwhoint/Trial2aspx?TrialID=DRKS00011609* 2017. | Ineligible publication type |
| Dunn HK, Litt MD. Decreased drinking in adults with co-occurring cannabis and alcohol use disorders in a treatment trial for marijuana dependence: evidence of a secondary benefit? *Addictive behaviors* 2019;**99**:106051. https://doi.org/10.1016/j.addbeh.2019.106051 | Ineligible outcome |
| Dupont HB, Candel M, Lemmens P, Kaplan CD, van de Mheen D, De Vries NK. Stages of Change Model has Limited Value in Explaining the Change in Use of Cannabis among Adolescent Participants in an | Ineligible study design |

| | |
|---|---|
| Efficacious Motivational Interviewing Intervention. *Journal of psychoactive drugs* 2017;**49**:363-72. https://doi.org/10.1080/02791072.2017.1325030 | |
| Dupont HB, Candel MJ, Kaplan CD, van de Mheen D, de Vries NK. Assessing the Efficacy of MOTI-4 for Reducing the Use of Cannabis Among Youth in the Netherlands: a Randomized Controlled Trial. *Journal of substance abuse treatment* 2016;**65**:6-12. https://doi.org/10.1016/j.jsat.2015.11.012 | Ineligible intervention |
| Easton CJ, Oberleitner LM, Scott MC, Crowley MJ, Babuscio TA, Carroll KM. Differences in treatment outcome among marijuana-dependent young adults with and without antisocial personality disorder. *American journal of drug and alcohol abuse* 2012;**38**:305-13. https://doi.org/10.3109/00952990.2011.643989 | Ineligible study design |
| Eisenberg K, Woodruff SI. Randomized controlled trial to evaluate screening and brief intervention for drug-using multiethnic emergency and trauma department patients. *Addiction science & clinical practice* 2013;**8**:8. https://doi.org/10.1186/1940-0640-8-8 | Ineligible population |
| Elliott JC, Carey KB, Vanable PA. A preliminary evaluation of a web-based intervention for college marijuana use. *Psychology of addictive behaviors* 2014;**28**:288-93. https://doi.org/10.1037/a0034995 | Ineligible population |
| Emery NN, Carpenter RW, Meisel SN, Miranda R. Effects of topiramate on the association between affect, cannabis craving, and cannabis use in the daily life of youth during a randomized clinical trial. *Psychopharmacology* 2021;**238**:3095-106. https://doi.org/10.1007/s00213-021-05925-5 | Ineligible intervention |
| Esposito-Smythers C, Brown LK, Wolff J, Xu J, Thornton S, Tidey J. Substance abuse treatment for HIV infected young people: an open pilot trial. *Journal of Substance Abuse Treatment* 2014;**46**:244-50. https://doi.org/10.1016/j.jsat.2013.07.008 | Ineligible study design |
| Farrow JA, Watts DH, Krohn MA, Olson HC. Pregnant adolescents in chemical dependency treatment. Description and outcomes. *Journal of substance abuse treatment* 1999;**16**:157-61. https://doi.org/10.1016/s0740-5472(98)00022-1 | Ineligible study design |
| Faulkner N, McCambridge J, Slym RL, Rollnick S. It ain't what you do, it's the way that you do it: a qualitative study of advice for young cannabis users. *Drug and alcohol review* 2009;**28**:129-34. https://doi.org/10.1111/j.1465-3362.2008.00033.x | Ineligible study design |
| Fernandes S, Ferigolo M, Benchaya MC, de Campos Moreira T, Pierozan PS, Mazoni CG*, et al.* Brief motivational intervention and telemedicine: A new perspective of treatment to marijuana users. *Addictive Behaviors* 2010;**35**:750-5. https://doi.org/10.1016/j.addbeh.2010.03.001 | Ineligible intervention |
| Ferreiro IC, Cuadra MAR, Serqueda FA, Abad JMH. Impact of Housing First on Psychiatric Symptoms, Substance Use, and Everyday Life Skills Among People Experiencing Homelessness. *Journal of psychosocial nursing and mental health services* 2022;**60**:46-55. https://doi.org/10.3928/02793695-20220316-01 | Ineligible population |
| Fischer B, Dawe M, McGuire F, Shuper PA, Capler R, Bilsker D*, et al.* Feasibility and impact of brief interventions for frequent cannabis users in Canada. *Journal of substance abuse treatment* 2013;**44**:132-8. https://doi.org/10.1016/j.jsat.2012.03.006 | Ineligible population |
| Fischer B, Jones W, Shuper P, Rehm J. 12-month follow-up of an exploratory 'brief intervention' for high-frequency cannabis users among Canadian university students. *Substance abuse treatment, prevention, and policy* 2012;**7**:15. https://doi.org/10.1186/1747-597X-7-15 | Ineligible population |
| Fox CL, Towe SL, Stephens RS, Walker DD, Roffman RA. Motives for cannabis use in high-risk adolescent users. *Psychology of addictive behaviors* 2011;**25**:492-500. https://doi.org/10.1037/a0024331 | Ineligible population |
| Freeman TP, Hindocha C, Baio G, Shaban NDC, Thomas EM, Astbury D*, et al.* Cannabidiol for the treatment of cannabis use disorder: a phase 2a, double-blind, placebo-controlled, randomised, adaptive Bayesian trial. *Lancet Psychiatry* 2020;**7**:865-74. https://doi.org/10.1016/S2215-0366(20)30290-X | Ineligible intervention* |
| French MT, Roebuck MC, Dennis ML, Diamond G, Godley SH, Tims F*, et al.* The economic cost of outpatient marijuana treatment for adolescents: findings from a multi-site field experiment. *Addiction* 2002;**97**:84-97. https://doi.org/10.1046/j.1360-0443.97.s01.4.x | Ineligible population |
| Frewen AR. An examination of withdrawal symptoms and their relationship with outcomes in a combined behavioural and pharmacological intervention for dependent cannabis users. *Macquarie university* 2009. | Ineligible publication type |
| Fuster D, Cheng DM, Wang N, Bernstein JA, Palfai TP, Alford DP*, et al.* Brief intervention for daily marijuana users identified by screening in primary care: a subgroup analysis of the ASPIRE randomized clinical trial. *Substance abuse* 2016;**37**:336-42. https://doi.org/10.1080/08897077.2015.1075932 | Ineligible population |
| Gantner A. Multidimensional family therapy for adolescent clients with cannabis use disorders--Results and experience from the INCANT pilot study. *Praxis der Kinderpsychologie und Kinderpsychiatrie* 2006;**55**:520-32. | Ineligible study design |
| Gantner A, Spohr B. Multidimensional family therapy in practice: Clinical experiences with adolescent cannabis abusers and their families. [*Multidimensionale familientherapie (MDFT) in der praxis: Therapeutische erfahrungen mit jugendlichen cannabisabhangigen und ihren familien]. Sucht* 2010;**56**:71-6. https://doi.org/10.1024/0939-5911/a000002 | Ineligible study design |
| Gates PJ, Norberg MM, Copeland J, Digiusto E. Randomized controlled trial of a novel cannabis use intervention delivered by telephone. *Addiction* 2012;**107**:2149-58. https://doi.org/10.1111/j.1360-0443.2012.03953.x | Ineligible intervention |

| | |
|---|---|
| Ghahari S, Zandnia F, Mazloumirad M, Ghayoomi R, Gheitarani B. The effectiveness of chair work intervention on anxiety and depression in divorced women using Cannabis. *Asian Journal of Psychiatry* 2019;**44**:161-2. https://doi.org/10.1016/j.ajp.2019.07.047 | Ineligible population |
| Gibbons CJ, Nich C, Steinberg K, Roffman RA, Corvino J, Babor TF, *et al.* Treatment process, alliance and outcome in brief versus extended treatments for marijuana dependence. *Addiction* 2010;**105**:1799-808. https://doi.org/10.1111/j.1360-0443.2010.03047.x | Ineligible outcome |
| Giguere S, Potvin S, Beaudoin M, Dellazizzo L, Giguere CE, Furtos A, *et al.* Avatar Intervention for Cannabis Use Disorder in Individuals with Severe Mental Disorders: a Pilot Study. *Journal of personalized medicine* 2023;**13**. https://doi.org/10.3390/jpm13050766 | Ineligible study design |
| Godley MD, Godley SH, Dennis ML, Funk R, Passetti LL. Preliminary outcomes from the assertive continuing care experiment for adolescents discharged from residential treatment. *Journal of substance abuse treatment* 2002;**23**:21-32. https://doi.org/10.1016/s0740-5472(02)00230-1 | Ineligible population |
| Godley MD, Godley SH, Dennis ML, Funk RR, Passetti LL. The effect of assertive continuing care on continuing care linkage, adherence and abstinence following residential treatment for adolescents with substance use disorders. *Addiction* 2007;**102**:81-93. https://doi.org/10.1111/j.1360-0443.2006.01648.x | Ineligible population |
| Godley MD, Godley SH, Dennis ML, Funk RR, Passetti LL, Petry NM. A randomized trial of assertive continuing care and contingency management for adolescents with substance use disorders. *Journal of consulting and clinical psychology* 2014;**82**:40-51. https://doi.org/10.1037/a0035264 | Ineligible population |
| Godley MD, Passetti LL, Subramaniam GA, Funk RR, Smith JE, Meyers RJ. Adolescent Community Reinforcement Approach implementation and treatment outcomes for youth with opioid problem use. *Drug and alcohol dependence* 2017;**174**:9-16. https://doi.org/10.1016/j.drugalcdep.2016.12.029 | Ineligible study design |
| Goldston DB, Curry JF, Wells KC, Kaminer Y, Daniel SS, Esposito-Smythers C, *et al.* Feasibility of an Integrated Treatment Approach for Youth with Depression, Suicide Attempts, and Substance Use Problems. *Evidence-based practice in child and adolescent mental health* 2021;**6**:155-72. https://doi.org/10.1080/23794925.2021.1888664 | Ineligible population |
| Gonzales R, Ang A, Murphy DA, Glik DC, Anglin MD. Substance use recovery outcomes among a cohort of youth participating in a mobile-based texting aftercare pilot program. *Journal of substance abuse treatment* 2014;**47**:20-6. https://doi.org/10.1016/j.jsat.2014.01.010 | Ineligible population |
| Goodness TM, Palfai TP. Electronic screening and brief intervention to reduce cannabis use and consequences among graduate students presenting to a student health center: a pilot study. *Addictive behaviors* 2020;**106**:106362. https://doi.org/10.1016/j.addbeh.2020.106362 | Ineligible population |
| Goti J, Diaz R, Serrano L, Gonzalez L, Calvo R, Gual A, *et al.* Brief intervention in substance-use among adolescent psychiatric patients: a randomized controlled trial. *European child & adolescent psychiatry* 2010;**19**:503-11. https://doi.org/10.1007/s00787-009-0060-5 | Ineligible population |
| Granholm E, Tate SR, Link PC, Lydecker KP, Cummins KM, McQuaid J, *et al.* Neuropsychological functioning and outcomes of treatment for co-occurring depression and substance use disorders. *American journal of drug and alcohol abuse* 2011;**37**:240-9. https://doi.org/10.3109/00952990.2011.570829 | Ineligible population |
| Gray JC, Treloar Padovano H, Wemm SE, Miranda R. Predictors of Topiramate Tolerability in Heavy Cannabis-Using Adolescents and Young Adults: a Secondary Analysis of a Randomized, Double-Blind, Placebo-Controlled Trial. *Journal of clinical psychopharmacology* 2018;**38**:134-7. https://doi.org/10.1097/JCP.0000000000000843 | Ineligible intervention |
| Gray KM, Carpenter MJ, Baker NL, DeSantis SM, Kryway E, Hartwell KJ, *et al.* "A double-blind randomized controlled trial of N-acetylcysteine in cannabis-dependent adolescents": Correction. *The American Journal of Psychiatry* 2012;**169**:869. | Ineligible publication type |
| Gray KM, Carpenter MJ, Baker NL, DeSantis SM, Kryway E, Hartwell KJ, *et al.* A double-blind randomized controlled trial of N-acetylcysteine in cannabis-dependent adolescents. *American journal of psychiatry* 2012;**169**:805-12. https://doi.org/10.1176/appi.ajp.2012.12010055 | Ineligible intervention* |
| Gray KM, Riggs PD, Min S-J, Mikulich-Gilbertson SK, Bandyopadhyay D, Winhusen T. Cigarette and cannabis use trajectories among adolescents in treatment for attention-deficit/hyperactivity disorder and substance use disorders. *Drug and Alcohol Dependence* 2011;**117**:242-7. https://doi.org/10.1016/j.drugalcdep.2011.02.005 | Ineligible study design |
| Gray KM, Sonne SC, McClure EA, Ghitza UE, Matthews AG, McRae-Clark AL, *et al.* A randomized placebo-controlled trial of N-acetylcysteine for cannabis use disorder in adults. *Drug and alcohol dependence* 2017;**177**:249-57. https://doi.org/10.1016/j.drugalcdep.2017.04.020 | Ineligible intervention* |
| Greenfield SF, Trucco EM, McHugh RK, Lincoln M, Gallop RJ. The Women's Recovery Group Study: a Stage I trial of women-focused group therapy for substance use disorders versus mixed-gender group drug counseling. *Drug and alcohol dependence* 2007;**90**:39-47. https://doi.org/10.1016/j.drugalcdep.2007.02.009 | Ineligible population |
| Gryczynski J, Mitchell SG, Ondersma SJ, O'Grady KE, Schwartz RP. Potential radiating effects of misusing substances among medical patients receiving brief intervention. *Journal of substance abuse treatment* 2015;**55**:39-44. https://doi.org/10.1016/j.jsat.2015.02.003 | Ineligible population |

| | |
|---|---|
| Gryczynski J, O'Grady KE, Mitchell SG, Ondersma SJ, Schwartz RP. Immediate Versus Delayed Computerized Brief Intervention for Illicit Drug Misuse. *Journal of addiction medicine* 2016;**10**:344-51. https://doi.org/10.1097/ADM.0000000000000248 | Ineligible population |
| Halliday-Boykins CA, Schaeffer CM, Henggeler SW, Chapman JE, Cunningham PB, Randall J*, et al.* Predicting nonresponse to juvenile drug court interventions. *Journal of substance abuse treatment* 2010;**39**:318-28. https://doi.org/10.1016/j.jsat.2010.07.011 | Ineligible study design |
| Heitmann J, van Hemel-Ruiter ME, Huisman M, Ostafin BD, Wiers RW, MacLeod C*, et al.* Effectiveness of attentional bias modification training as add-on to regular treatment in alcohol and cannabis use disorder: a multicenter randomized control trial. *PloS one* 2021;**16**:e0252494. https://doi.org/10.1371/journal.pone.0252494 | Ineligible population |
| Heitmann J, van Hemel-Ruiter ME, Vermeulen KM, Ostafin BD, MacLeod C, Wiers RW*, et al.* Internet-based attentional bias modification training as add-on to regular treatment in alcohol and cannabis dependent outpatients: a study protocol of a randomized control trial. *BMC Psychiatry* 2017;**17**:193. https://doi.org/10.1186/s12888-017-1359-2 | Ineligible publication type |
| Hendriks V, van der Schee E, Blanken P. Treatment of adolescents with a cannabis use disorder: main findings of a randomized controlled trial comparing multidimensional family therapy and cognitive behavioral therapy in The Netherlands. *Drug and alcohol dependence* 2011;**119**:64-71. https://doi.org/10.1016/j.drugalcdep.2011.05.021 | Ineligible outcome[c] |
| Hendriks V, van der Schee E, Blanken P. Matching adolescents with a cannabis use disorder to multidimensional family therapy or cognitive behavioral therapy: treatment effect moderators in a randomized controlled trial. *Drug and alcohol dependence* 2012;**125**:119-26. https://doi.org/10.1016/j.drugalcdep.2012.03.023 | Ineligible outcome[c] |
| Hendriks VM, Van Der Schee E, Blanken P. Multidimensional family therapy and cognitive behavioral therapy in adolescents with a cannabis use disorder: a randomised controlled study. *Tijdschrift voor psychiatrie* 2013;**55**:747-59. | Ineligible outcome[c] |
| Henggeler SW, Clingempeel WG, Brondino MJ, Pickrel SG. Four-year follow-up of multisystemic therapy with substance-abusing and substance-dependent juvenile offenders. *Journal of the American Academy of Child and Adolescent Psychiatry* 2002;**41**:868-74. https://doi.org/10.1097/00004583-200207000-00021 | Ineligible population |
| Henggeler SW, Pickrel SG, Brondino MJ. Multisystemic treatment of substance-abusing and dependent delinquents: outcomes, treatment fidelity, and transportability. *Mental health services research* 1999;**1**:171-84. https://doi.org/10.1023/a:1022373813261 | Ineligible population |
| Herbst ED, Pennington DL, Borsari B, Manuel J, Yalch M, Alcid E*, et al.* N-acetylcysteine for smoking cessation among dual users of tobacco and cannabis: protocol and rationale for a randomized controlled trial. *Contemporary clinical trials* 2023;**131**:107250. https://doi.org/10.1016/j.cct.2023.107250 | Ineligible publication type |
| Hides L, Carroll S, Scott R, Cotton S, Baker A, Lubman DI. Quik fix: A randomized controlled trial of an enhanced brief motivational interviewing intervention for alcohol/cannabis and psychological distress in young people. *Psychotherapy and Psychosomatics* 2013;**82**:122-4. https://doi.org/10.1159/000341921 | Ineligible population |
| Hill KP, Palastro MD, Gruber SA, Fitzmaurice GM, Greenfield SF, Lukas SE*, et al.* Nabilone pharmacotherapy for cannabis dependence: a randomized, controlled pilot study. *The American journal on addictions* 2017;**26**:795-801. https://doi.org/10.1111/ajad.12622 | Ineligible intervention* |
| Hirschtritt ME, Pagano ME, Christian KM, McNamara NK, Stansbrey RJ, Lingler J*, et al.* Moderators of fluoxetine treatment response for children and adolescents with comorbid depression and substance use disorders. *Journal of substance abuse treatment* 2012;**42**:366-72. https://doi.org/10.1016/j.jsat.2011.09.010 | Ineligible population |
| Hjorthoj C, Posselt CM, Baandrup L. Cannabidiol for cannabis use disorder: Too high hopes? *The Lancet Psychiatry* 2020;**7**:838-9. https://doi.org/10.1016/S2215-0366%2820%2930378-3 | Ineligible publication type |
| Hoch E, Buhringer G, Henker J, Rohrbacher H, Noack R, Pixa A*, et al.* Design of the CANDIS*-study for the treatment of cannabis use disorders: an example of translational research. *Sucht* 2011;**57**:183-92. https://doi.org/10.1024/0939-5911.a000111 | Ineligible study design |
| Hoch E, Noack R, Henker J, Pixa A, Höfler M, Behrendt S*, et al.* Efficacy of a targeted cognitive-behavioral treatment program for cannabis use disorders (CANDIS). *European neuropsychopharmacology* 2012;**22**:267-80. https://doi.org/10.1016/j.euroneuro.2011.07.014 | Ineligible study design |
| Houck JM, Feldstein Ewing SW. Working memory capacity and addiction treatment outcomes in adolescents. *American journal of drug and alcohol abuse* 2018;**44**:185-92. https://doi.org/10.1080/00952990.2017.1344680 | Ineligible population |
| Hser YI, Mooney LJ, Huang D, Zhu Y, Tomko RL, McClure E*, et al.* Reductions in cannabis use are associated with improvements in anxiety, depression, and sleep quality, but not quality of life. *Journal of substance abuse treatment* 2017;**81**:53-8. https://doi.org/10.1016/j.jsat.2017.07.012 | Ineligible study design |
| Humeniuk R, Ali R, Babor T, Souza-Formigoni ML, de Lacerda RB, Ling W*, et al.* A randomized controlled trial of a brief intervention for illicit drugs linked to the Alcohol, Smoking and Substance Involvement | Ineligible population |

| | |
|---|---|
| Screening Test (ASSIST) in clients recruited from primary health-care settings in four countries. *Addiction* 2012;**107**:957-66. https://doi.org/10.1111/j.1360-0443.2011.03740.x | |
| Imel ZE, Baer JS, Martino S, Ball SA, Carroll KM. Mutual influence in therapist competence and adherence to motivational enhancement therapy. *Drug and alcohol dependence* 2011;**115**:229-36. https://doi.org/10.1016/j.drugalcdep.2010.11.010 | Ineligible study design |
| ISRCTN. An evaluation of a brief assessment-led intervention with young non-injecting drug users. *https://trialsearchwhoint/Trial2aspx?TrialID=ISRCTN43192662* 2008. | Ineligible publication type |
| ISRCTN. Mentalisation-Based Treatment for Dual Diagnoses. *https://trialsearchwhoint/Trial2aspx?TrialID=ISRCTN98982683* 2009. | Ineligible publication type |
| ISRCTN. Evaluation of an internet-based one-time counselling intervention for young cannabis- or alcohol-users. *https://trialsearchwhoint/Trial2aspx?TrialID=ISRCTN79857771* 2010. | Ineligible publication type |
| ISRCTN. Evaluation of an online help program for the treatment of depression and drug abuse in Mexico. *https://trialsearchwhoint/Trial2aspx?TrialID=ISRCTN25429892* 2014. | Ineligible publication type |
| Johnston J, Lintzeris N, Allsop DJ, Suraev A, Booth J, Carson DS, *et al.* Lithium carbonate in the management of cannabis withdrawal: a randomized placebo-controlled trial in an inpatient setting. *Psychopharmacology* 2014;**231**:4623-36. https://doi.org/10.1007/s00213-014-3611-5 | Ineligible intervention |
| Jonas B, Tensil MD, Leuschner F, Struber E, Tossmann P. Predictors of treatment response in a web-based intervention for cannabis users. *Internet interventions* 2019;**18**. https://doi.org/10.1016/j.invent.2019.100261 | Ineligible study design |
| Jonas B, Tensil MD, Tossmann P, Strüber E. Effects of Treatment Length and Chat-Based Counseling in a Web-Based Intervention for Cannabis Users: randomized Factorial Trial. *Journal of medical Internet research* 2018;**20**:e166. https://doi.org/10.2196/jmir.9579 | Ineligible intervention |
| Jungerman FS, Andreoni S, Laranjeira R. Short term impact of same intensity but different duration interventions for cannabis users. *Drug and alcohol dependence* 2007;**90**:120-7. https://doi.org/10.1016/j.drugalcdep.2007.02.019 | Ineligible intervention |
| Kadden RM, Litt MD, Kabela-Cormier E, Petry NM. Increased drinking in a trial of treatments for marijuana dependence: substance substitution? *Drug and alcohol dependence* 2009;**105**:168-71. https://doi.org/10.1016/j.drugalcdep.2009.05.024 | Ineligible study design |
| Kalapatapu RK, Campbell A, Aharonovich E, Hu MC, Levin FR, Nunes EV. Demographic and clinical characteristics of middle-aged versus younger adults enrolled in a clinical trial of a web-delivered psychosocial treatment for substance use disorders. *Journal of addiction medicine* 2013;**7**:66-72. https://doi.org/10.1097/ADM.0b013e31827e2d04 | Ineligible study design |
| Kaminer Y, Burleson JA, Burke R, Litt MD. The efficacy of contingency management for adolescent cannabis use disorder: a controlled study. *Substance abuse* 2014;**35**:391-8. https://doi.org/10.1080/08897077.2014.933724 | Ineligible population |
| Kaminer Y, Ohannessian C, Burke R. Retention and treatment outcome of youth with cannabis use disorder referred by the legal system. *Adolescent psychiatry* 2019;**9**:4-10. https://doi.org/10.2174/2210676608666181102145040 | Ineligible study design |
| Kamon J, Budney A, Stanger C. A contingency management intervention for adolescent marijuana abuse and conduct problems. *Journal of the American Academy of Child and Adolescent Psychiatry* 2005;**44**:513-21. https://doi.org/10.1097/01.chi.0000159949.82759.64 | Ineligible study design |
| Kay-Lambkin F, Baker A, Lewin T, Carr V. Acceptability of a clinician-assisted computerized psychological intervention for comorbid mental health and substance use problems: treatment adherence data from a randomized controlled trial. *Journal of medical Internet research* 2011;**13**:e11. https://doi.org/10.2196/jmir.1522 | Ineligible population |
| Kay-Lambkin FJ, Baker AL, Kelly B, Lewin TJ. Clinician-assisted computerised versus therapist-delivered treatment for depressive and addictive disorders: a randomised controlled trial. *Medical journal of Australia* 2011;**195**:S44-50. https://doi.org/10.5694/j.1326-5377.2011.tb03265.x | Ineligible population |
| Kay-Lambkin FJ, Baker AL, Lewin TJ, Carr VJ. Computer-based psychological treatment for comorbid depression and problematic alcohol and/or cannabis use: a randomized controlled trial of clinical efficacy. *Addiction* 2009;**104**:378-88. https://doi.org/10.1111/j.1360-0443.2008.02444.x | Ineligible population |
| Kay-Lambkin FJ, Baker AL, Palazzi K, Lewin TJ, Kelly BJ. Therapeutic Alliance, Client Need for Approval, and Perfectionism as Differential Moderators of Response to eHealth and Traditionally Delivered Treatments for Comorbid Depression and Substance Use Problems. *International journal of behavioral medicine* 2017;**24**:728-39. https://doi.org/10.1007/s12529-017-9676-x | Ineligible population |
| Kells M, Burke PJ, Parker S, Jonestrask C, Shrier LA. Engaging Youth (Adolescents and Young Adults) to Change Frequent Marijuana Use: motivational Enhancement Therapy (MET) in Primary Care. *Journal of pediatric nursing* 2019;**49**:24-30. https://doi.org/10.1016/j.pedn.2019.08.011 | Ineligible study design |
| Kelly JF, Kaminer Y, Kahler CW, Hoeppner B, Yeterian J, Cristello JV*, et al.* A pilot randomized clinical trial testing integrated 12-Step facilitation (iTSF) treatment for adolescent substance use disorder. *Addiction* 2017;**112**:2155-66. https://doi.org/10.1111/add.13920 | Ineligible population |

| | |
|---|---|
| Kemp K, Micalizzi L, Becker SJ, Cheaito A, Suazo NC, Fox K*, et al.* Intervention for marijuana using, court-involved non-incarcerated youth. *Journal of substance use and addiction treatment* 2023;**152**:209100. https://doi.org/10.1016/j.josat.2023.209100 | Ineligible population |
| Killeen TK, McRae-Clark AL, Waldrop AE, Upadhyaya H, Brady KT. Contingency management in community programs treating adolescent substance abuse: a feasibility study. *Journal of child and adolescent psychiatric nursing* 2012;**25**:33-41. https://doi.org/10.1111/j.1744-6171.2011.00313.x | Ineligible population |
| Kim TW, Bernstein J, Cheng DM, Lloyd-Travaglini C, Samet JH, Palfai TP*, et al.* Receipt of addiction treatment as a consequence of a brief intervention for drug use in primary care: a randomized trial. *Addiction* 2017;**112**:818-27. https://doi.org/10.1111/add.13701 | Ineligible population |
| Kober H, Devito EE, Deleone CM, Carroll KM, Potenza MN. Cannabis abstinence during treatment and one-year follow-up: relationship to neural activity in men. *Neuropsychopharmacology* 2014;**39**:2288-98. https://doi.org/10.1038/npp.2014.82 | Ineligible study design |
| Lamontagne Y, Hand I, Annable L, Gagnon MA. Physiological and psychological effects of biological feed-back training (alpha and EMG) among drug using college students. *Encephale* 1977;**3**:203-6. | Ineligible population |
| Lang E, Engelander M, Brooke T. Report of an integrated brief intervention with self-defined problem cannabis users. *Journal of substance abuse treatment* 2000;**19**:111-6. https://doi.org/10.1016/s0740-5472(99)00104-x | Ineligible study design |
| Laporte C, Vaillant-Roussel H, Pereira B, Blanc O, Eschalier B, Kinouani S*, et al.* Cannabis and Young Users-A Brief Intervention to Reduce Their Consumption (CANABIC): a Cluster Randomized Controlled Trial in Primary Care. *Annals of family medicine* 2017;**15**:131-9. https://doi.org/10.1370/afm.2003 | Ineligible population |
| Laporte C, Vaillant-Roussel H, Pereira B, Blanc O, Tanguy G, Frappe P*, et al.* CANABIC: CANnabis and Adolescents: effect of a Brief Intervention on their Consumption--study protocol for a randomized controlled trial. *Trials* 2014;**15**:40. https://doi.org/10.1186/1745-6215-15-40 | Ineligible publication type |
| Lascaux M, Bastard N, Bonnaire C, Couteron J-P, Phan O. INCANT: Comparison of two formalized therapeutic models. *INCANT Une comparaison de deux modeles therapeutiques formalises* 2010;**32**:209-19. | Ineligible study design |
| Lascaux M, Ionescu S, Phan O. Effectiveness of formalised therapy for adolescents with cannabis dependence: A randomised trial. *Drugs: Education, Prevention & Policy* 2016;**23**:404-9. https://doi.org/10.3109/09687637.2016.1153603 | Ineligible intervention |
| Lascaux M, Phan O. Comparison of European therapies for cannabis addiction among adolescents. *Encephale* 2015;**41**:S21-8. https://doi.org/10.1016/j.encep.2014.10.013 | Ineligible study design |
| Latimer WW, Winters KC, D'Zurilla T, Nichols M. Integrated family and cognitive-behavioral therapy for adolescent substance abusers: a stage I efficacy study. *Drug and alcohol dependence* 2003;**71**:303-17. https://doi.org/10.1016/s0376-8716(03)00171-6 | Ineligible population |
| Lee CM, Kilmer JR, Neighbors C, Atkins DC, Zheng C, Walker DD*, et al.* Indicated prevention for college student marijuana use: a randomized controlled trial. *Journal of consulting and clinical psychology* 2013;**81**:702-9. https://doi.org/10.1037/a0033285 | Ineligible population |
| Lee CM, Neighbors C, Kilmer JR, Larimer ME. A brief, web-based personalized feedback selective intervention for college student marijuana use: a randomized clinical trial. *Psychology of addictive behaviors* 2010;**24**:265-73. https://doi.org/10.1037/a0018859 | Ineligible population |
| Lee DC, Budney AJ, Brunette MF, Hughes JR, Etter JF, Stanger C. Outcomes from a computer-assisted intervention simultaneously targeting cannabis and tobacco use. *Drug and alcohol dependence* 2015;**155**:134-40. https://doi.org/10.1016/j.drugalcdep.2015.08.001 | Ineligible study design |
| Lee DC, Schlienz NJ, Herrmann ES, Martin EL, Leoutsakos J, Budney AJ*, et al.* Randomized controlled trial of zolpidem as a pharmacotherapy for cannabis use disorder. *Journal of substance use and addiction treatment* 2024;**156**:209180. https://doi.org/10.1016/j.josat.2023.209180 | Ineligible intervention |
| Lee DC, Walker DD, Hughes JR, Brunette MF, Scherer E, Stanger C*, et al.* Sequential and simultaneous treatment approaches to cannabis use disorder and tobacco use. *Journal of substance abuse treatment* 2019;**98**:39-46. https://doi.org/10.1016/j.jsat.2018.12.005 | Ineligible intervention |
| Lees R, Hines LA, Hindocha C, Baio G, Shaban NDC, Stothart G*, et al.* Effect of four-week cannabidiol treatment on cognitive function: secondary outcomes from a randomised clinical trial for the treatment of cannabis use disorder. *Psychopharmacology* 2023;**240**:337-46. https://doi.org/10.1007/s00213-022-06303-5 | Ineligible outcome |
| Legenbauer T, Baldus C, Jorke C, Kaffke L, Pepic A, Daubmann A*, et al.* Mind it! A mindfulness-based group psychotherapy for substance use disorders in adolescent inpatients. *European child & adolescent psychiatry* 2024; 10.1007/s00787-024-02465-z. https://doi.org/10.1007/s00787-024-02465-z | Ineligible population |
| Lévesque A, Campbell AN, Pavlicova M, Hu MC, Walker R, McClure EA*, et al.* Coping strategies as a mediator of internet-delivered psychosocial treatment: secondary analysis from a NIDA CTN multisite effectiveness trial. *Addictive behaviors* 2017;**65**:74-80. https://doi.org/10.1016/j.addbeh.2016.09.012 | Ineligible population |
| Levin FR, Mariani J, Brooks DJ, Pavlicova M, Nunes EV, Agosti V*, et al.* A randomized double-blind, placebo-controlled trial of venlafaxine-extended release for co-occurring cannabis dependence and depressive disorders. *Addiction* 2013;**108**:1084-94. https://doi.org/10.1111/add.12108 | Ineligible intervention* |

| | |
|---|---|
| Levin FR, Mariani JJ, Brooks DJ, Pavlicova M, Cheng W, Nunes EV. Dronabinol for the treatment of cannabis dependence: a randomized, double-blind, placebo-controlled trial. *Drug and alcohol dependence* 2011;**116**:142-50. https://doi.org/10.1016/j.drugalcdep.2010.12.010 | Ineligible intervention* |
| Levin FR, Mariani JJ, Choi CJ, Basaraba C, Brooks DJ, Brezing CA*, et al.* Non-abstinent treatment outcomes for cannabis use disorder. *Drug and alcohol dependence* 2021;**225**:108765. https://doi.org/10.1016/j.drugalcdep.2021.108765 | Ineligible intervention* |
| Levin FR, Mariani JJ, Pavlicova M, Brooks D, Glass A, Mahony A*, et al.* Dronabinol and lofexidine for cannabis use disorder: a randomized, double-blind, placebo-controlled trial. *Drug and alcohol dependence* 2016;**159**:53-60. https://doi.org/10.1016/j.drugalcdep.2015.11.025 | Ineligible intervention* |
| Levin FR, McDowell D, Evans SM, Nunes E, Akerele E, Donovan S*, et al.* Pharmacotherapy for marijuana dependence: a double-blind, placebo-controlled pilot study of divalproex sodium. *The American journal on addictions* 2004;**13**:21-32. https://doi.org/10.1080/10550490490265280 | Ineligible intervention* |
| Liddle HA, Dakof GA, Parker K, Diamond GS, Barrett K, Tejeda M. Multidimensional family therapy for adolescent drug abuse: results of a randomized clinical trial. *American journal of drug and alcohol abuse* 2001;**27**:651-88. https://doi.org/10.1081/ada-100107661 | Ineligible population |
| Lin JA, Harris SK, Shrier LA. Trait mindfulness and cannabis use-related factors in adolescents and young adults with frequent use. *Substance abuse* 2021;**42**:968-73. https://doi.org/10.1080/08897077.2021.1901179 | Ineligible intervention |
| Lintzeris N, Bhardwaj A, Mills L, Dunlop A, Copeland J, McGregor I*, et al.* Nabiximols for the Treatment of Cannabis Dependence: A Randomized Clinical Trial. *JAMA Internal Medicine* 2019;**179**:1242-53. https://doi.org/10.1001/jamainternmed.2019.1993 | Ineligible intervention* |
| Lintzeris N, Mills L, Dunlop A, Copeland J, McGregor I, Bruno R*, et al.* Cannabis use in patients 3 months after ceasing nabiximols for the treatment of cannabis dependence: results from a placebo-controlled randomised trial. *Drug and alcohol dependence* 2020;**215**:108220. https://doi.org/10.1016/j.drugalcdep.2020.108220 | Ineligible intervention* |
| Litt MD, Kadden RM. Willpower versus "skillpower": examining how self-efficacy works in treatment for marijuana dependence. *Psychology of addictive behaviors* 2015;**29**:532-40. https://doi.org/10.1037/adb0000085 | Ineligible study design |
| Litt MD, Kadden RM, Stephens RS. Coping and self-efficacy in marijuana treatment: results from the marijuana treatment project. *Journal of consulting and clinical psychology* 2005;**73**:1015-25. https://doi.org/10.1037/0022-006X.73.6.1015 | Ineligible outcome |
| Lozano BE, Stephens RS, Roffman RA. Abstinence and moderate use goals in the treatment of marijuana dependence. *Addiction* 2006;**101**:1589-97. https://doi.org/10.1111/j.1360-0443.2006.01609.x | Ineligible outcome |
| Lu S, Cameron K, Ganesan S, Feldman B, McKenna M. A double-blind placebo control pilot study on the safety and tolerability of Nabilone in marijuana users. *Mental Health and Substance Use* 2013;**6**:133-9. https://doi.org/10.1080/17523281.2012.693520 | Ineligible population |
| Macatee RJ, Albanese BJ, Okey SA, Afshar K, Carr M, Rosenthal MZ*, et al.* Impact of a computerized intervention for high distress intolerance on cannabis use outcomes: a randomized controlled trial. *Journal of substance abuse treatment* 2021;**121**:108194. https://doi.org/10.1016/j.jsat.2020.108194 | Ineligible intervention |
| Mariani JJ, Pavlicova M, Jean Choi C, Basaraba C, Carpenter KM, Mahony AL*, et al.* Quetiapine treatment for cannabis use disorder. *Drug and Alcohol Dependence* 2021;**218**. https://doi.org/10.1016/j.drugalcdep.2020.108366 | Ineligible intervention* |
| Marín-Navarrete R, Templos-Nuñez L, Eliosa-Hernández A, Villalobos-Gallegos L, Fernández-Mondragón J, Pérez-López A*, et al.* Characteristics of a treatment-seeking population in outpatient addiction treatment centers in Mexico. *Substance use & misuse* 2014;**49**:1784-94. https://doi.org/10.3109/10826084.2014.931972 | Ineligible study design |
| Mason BJ, Crean R, Goodell V, Light JM, Quello S, Shadan F*, et al.* A proof-of-concept randomized controlled study of gabapentin: effects on cannabis use, withdrawal and executive function deficits in cannabis-dependent adults. *Neuropsychopharmacology* 2012;**37**:1689-98. https://doi.org/10.1038/npp.2012.14 | Ineligible intervention* |
| Mason MJ. Depressive symptoms moderate cannabis use for young adults in a Text-Delivered randomized clinical trial for cannabis use disorder. *Addictive behaviors* 2020;**104**:106259. https://doi.org/10.1016/j.addbeh.2019.106259 | Ineligible intervention |
| Mason MJ, Moore M, Brown A. Young adults' perceptions of acceptability and effectiveness of a text message-delivered treatment for cannabis use disorder. *Journal of substance abuse treatment* 2018;**93**:15-8. https://doi.org/10.1016/j.jsat.2018.07.007 | Ineligible study design |
| Mason MJ, Zaharakis NM, Russell M, Childress V. A pilot trial of text-delivered peer network counseling to treat young adults with cannabis use disorder. *Journal of substance abuse treatment* 2018;**89**:1-10. https://doi.org/10.1016/j.jsat.2018.03.002 | Ineligible intervention |
| McCambridge J, Day M, Thomas BA, Strang J. Fidelity to Motivational Interviewing and subsequent cannabis cessation among adolescents. *Addictive behaviors* 2011;**36**:749-54. https://doi.org/10.1016/j.addbeh.2011.03.002 | Ineligible population |

| | |
|---|---|
| McClure EA, Baker NL, Gray KM. Cigarette smoking during an N-acetylcysteine-assisted cannabis cessation trial in adolescents. *American journal of drug and alcohol abuse* 2014;**40**:285-91. https://doi.org/10.3109/00952990.2013.878718 | Ineligible intervention* |
| McClure EA, Baker NL, Sonne SC, Ghitza UE, Tomko RL, Montgomery L*, et al.* Tobacco use during cannabis cessation: Use patterns and impact on abstinence in a National Drug Abuse Treatment Clinical Trials Network study. *Drug Alcohol Depend* 2018;**192**:59-66. https://doi.org/10.1016/j.drugalcdep.2018.07.018 | Ineligible outcome |
| McClure EA, King JS, Wahle A, Matthews AG, Sonne SC, Lofwall MR*, et al.* Comparing adult cannabis treatment-seekers enrolled in a clinical trial with national samples of cannabis users in the United States. *Drug Alcohol Depend* 2017;**176**:14-20. https://doi.org/10.1016/j.drugalcdep.2017.02.024 | Ineligible study design |
| McClure EA, Sonne SC, Winhusen T, Carroll KM, Ghitza UE, McRae-Clark AL*, et al.* Achieving cannabis cessation -- evaluating N-acetylcysteine treatment (ACCENT): design and implementation of a multi-site, randomized controlled study in the National Institute on Drug Abuse Clinical Trials Network. *Contemporary clinical trials* 2014;**39**:211-23. https://doi.org/10.1016/j.cct.2014.08.011 | Ineligible intervention* |
| McLellan AT. A randomized controlled trial of brief cognitive-behavioral interventions for cannabis use disorder. *Journal of substance abuse treatment* 2001;**21**:65-6. https://doi.org/10.1016/S0740-5472(01)00196-9 | Ineligible publication type |
| McRae-Clark AL, Baker NL, Gray KM, Killeen T, Hartwell KJ, Simonian SJ. Vilazodone for cannabis dependence: a randomized, controlled pilot trial. *The American journal on addictions* 2016;**25**:69-75. https://doi.org/10.1111/ajad.12324 | Ineligible intervention |
| McRae-Clark AL, Baker NL, Gray KM, Killeen TK, Wagner AM, Brady KT*, et al.* Buspirone treatment of cannabis dependence: a randomized, placebo-controlled trial. *Drug and alcohol dependence* 2015;**156**:29-37. https://doi.org/10.1016/j.drugalcdep.2015.08.013 | Ineligible intervention |
| McRae-Clark AL, Baker NL, Sonne SC, DeVane CL, Wagner A, Norton J. Concordance of Direct and Indirect Measures of Medication Adherence in A Treatment Trial for Cannabis Dependence. *Journal of Substance Abuse Treatment* 2015;**57**:70-4. https://doi.org/10.1016/j.jsat.2015.05.002 | Ineligible study design |
| McRae-Clark AL, Carter RE, Killeen TK, Carpenter MJ, Wahlquist AE, Simpson SA*, et al.* A placebo-controlled trial of buspirone for the treatment of marijuana dependence. *Drug and alcohol dependence* 2009;**105**:132-8. https://doi.org/10.1016/j.drugalcdep.2009.06.022 | Ineligible intervention |
| McRae-Clark AL, Carter RE, Killeen TK, Carpenter MJ, White KG, Brady KT. A placebo-controlled trial of atomoxetine in marijuana-dependent individuals with attention deficit hyperactivity disorder. *The American journal on addictions* 2010;**19**:481-9. https://doi.org/10.1111/j.1521-0391.2010.00076.x | Ineligible intervention |
| McRae-Clark AL, Gray KM, Baker NL, Sherman BJ, Squeglia L, Sahlem GL*, et al.* Varenicline as a treatment for cannabis use disorder: a placebo-controlled pilot trial. *Drug and alcohol dependence* 2021;**229**:109111. https://doi.org/10.1016/j.drugalcdep.2021.109111 | Ineligible intervention |
| Meisel SN, Carpenter RW, Treloar Padovano H, Miranda R. Day-level shifts in social contexts during youth cannabis use treatment. *Journal of consulting and clinical psychology* 2021;**89**:251-63. https://doi.org/10.1037/ccp0000647 | Ineligible study design |
| Meisel SN, Treloar Padovano H, Miranda R. Combined pharmacotherapy and evidence-based psychosocial Cannabis treatment for youth and selection of cannabis-using friends. *Drug and alcohol dependence* 2021;**225**:108747. https://doi.org/10.1016/j.drugalcdep.2021.108747 | Ineligible population |
| Mertens JR, Ward CL, Bresick GF, Broder T, Weisner CM. Effectiveness of nurse-practitioner-delivered brief motivational intervention for young adult alcohol and drug use in primary care in South Africa: a randomized clinical trial. *Alcohol and alcoholism* 2014;**49**:430-8. https://doi.org/10.1093/alcalc/agu030 | Ineligible population |
| Mestre-Pinto JI, Fonseca F, Schaub MP, Baumgartner C, Alias-Ferri M, Torrens M. CANreduce-SP-adding psychological support to web-based adherence-focused guided self-help for cannabis users: study protocol for a three-arm randomized control trial. *Trials* 2022;**23**:524. https://doi.org/10.1186/s13063-022-06399-2 | Ineligible publication type |
| Mian MN, Earleywine M. Savoring as an intervention for cannabis use: Acceptability, feasibility, and preliminary results. *Addiction Research & Theory* 2023;**31**:296-305. https://doi.org/10.1080/16066359.2022.2160871 | Ineligible population |
| Mills L, Dunlop A, Montebello M, Copeland J, Bruno R, Jefferies M*, et al.* Correlates of treatment engagement and client outcomes: results of a randomised controlled trial of nabiximols for the treatment of cannabis use disorder. *Substance abuse treatment, prevention, and policy* 2022;**17**:67. https://doi.org/10.1186/s13011-022-00493-z | Ineligible intervention* |
| Mills L, Lintzeris N, Bruno R, Montebello M, Dunlop A, Deacon RM*, et al.* Validation of the Australian Treatment Outcomes Profile for use in clients with cannabis dependence. *Drug and alcohol review* 2020;**39**:356-64. https://doi.org/10.1111/dar.13050 | Ineligible intervention* |
| Miranda R, Treloar H, Blanchard A, Justus A, Monti PM, Chun T*, et al.* Topiramate and motivational enhancement therapy for cannabis use among youth: a randomized placebo-controlled pilot study. *Addiction biology* 2017;**22**:779-90. https://doi.org/10.1111/adb.12350 | Ineligible intervention |

| | |
|---|---|
| Moitra E, Anderson BJ, Stein MD. REDUCTIONS IN CANNABIS USE ARE ASSOCIATED WITH MOOD IMPROVEMENT IN FEMALE EMERGING ADULTS. *Depression and anxiety* 2016;**33**:332-8. https://doi.org/10.1002/da.22460 | Ineligible study design |
| Montebello M, Jefferies M, Mills L, Bruno R, Copeland J, McGregor I*, et al.* Mood, sleep and pain comorbidity outcomes in cannabis dependent patients: findings from a nabiximols versus placebo randomised controlled trial. *Drug and alcohol dependence* 2022;**234**:109388. https://doi.org/10.1016/j.drugalcdep.2022.109388 | Ineligible study design |
| Montgomery L, McClure EA, Tomko RL, Sonne SC, Winhusen T, Terry GE*, et al.* Blunts versus joints: cannabis use characteristics and consequences among treatment-seeking adults. *Drug and alcohol dependence* 2019;**198**:105-11. https://doi.org/10.1016/j.drugalcdep.2019.01.041 | Ineligible study design |
| Moore BA, Budney AJ. Abstinence at intake for marijuana dependence treatment predicts response. *Drug and alcohol dependence* 2002;**67**:249-57. https://doi.org/10.1016/S0376-8716(02)00079-0 | Ineligible study design |
| Murphy SM, Campbell AN, Ghitza UE, Kyle TL, Bailey GL, Nunes EV*, et al.* Cost-effectiveness of an internet-delivered treatment for substance abuse: data from a multisite randomized controlled trial. *Drug and alcohol dependence* 2016;**161**:119-26. https://doi.org/10.1016/j.drugalcdep.2016.01.021 | Ineligible population |
| NCT. Marijuana Treatment Project - 3. *https://clinicaltrialsgov/show/NCT00107588* 2005. | Ineligible publication type |
| NCT. Study Comparing Two Types of Psychotherapy for Treating Depression and Substance Abuse. *https://clinicaltrialsgov/show/NCT00108407* 2005. | Ineligible publication type |
| NCT. Effectiveness of Selegiline in Treating Marijuana Dependent Individuals - 1. *https://clinicaltrialsgov/show/NCT00218517* 2005. | Ineligible publication type |
| NCT. Therapeutic Substance Abuse Treatment in Pregnancy - 1. *https://clinicaltrialsgov/show/NCT00227903* 2005. | Ineligible publication type |
| NCT. Effectiveness of Nefazodone and Bupropion in Treating Marijuana Dependent Individuals. *https://clinicaltrialsgov/show/NCT00249509* 2005. | Ineligible publication type |
| NCT. Reducing Barriers to Drug Abuse Treatment Services. *https://clinicaltrialsgov/show/NCT00273845* 2006. | Ineligible publication type |
| NCT. INCA - Intervention and Neuropsychology in Cannabis Abuse. *https://clinicaltrialsgov/show/NCT00279604* 2006. | Ineligible publication type |
| NCT. Effectiveness Study of Dronabinol and BRENDA for the Treatment of Cannabis Withdrawal. *https://clinicaltrialsgov/show/NCT00480441* 2007. | Ineligible publication type |
| NCT. A Comparison of Adolescent Group Therapy and Transitional Family Therapy for Adolescent Alcohol and Drug Abusers. *https://clinicaltrialsgov/show/NCT00484367* 2007. | Ineligible publication type |
| NCT. Treatment for Adolescent Marijuana Abuse. *https://clinicaltrialsgov/show/NCT00580671* 2007. | Ineligible publication type |
| NCT. Efficacy of Contingency Management in the Treatment of Adolescents With Cannabis Use Disorders. *https://clinicaltrialsgov/show/NCT00878852* 2009. | Ineligible publication type |
| NCT. Computer-delivered Psychosocial Intervention for Adolescent Substance Use Disorders. *https://clinicaltrialsgov/show/NCT00957775* 2009. | Ineligible publication type |
| NCT. Treatment for Cannabis Withdrawal and Dependence. *https://clinicaltrialsgov/show/NCT01611948* 2011. | Ineligible publication type |
| NCT. Family and Adolescent Motivational Incentives for Leveraging Youth. *https://clinicaltrialsgov/show/NCT01736995* 2012. | Ineligible publication type |
| NCT. Pharmacological Treatment of Comorbid Alcohol and Marijuana Withdrawal and Dependence. *https://clinicaltrialsgov/show/NCT02210195* 2014. | Ineligible publication type |
| NCT. N-Acetylcysteine for Youth Cannabis Use Disorder. *https://clinicaltrialsgov/ct2/show/NCT03055377* 2017. | Ineligible publication type |
| NCT. Peer MI for Substance-using Emerging Adults. *https://clinicaltrialsgov/show/NCT03264872* 2017. | Ineligible publication type |
| NCT. Fatty Acid Amide Hydrolase (FAAH) Inhibitor Treatment of Cannabis Use Disorder (CUD). *https://clinicaltrialsgov/ct2/show/NCT03386487* 2017. | Ineligible publication type |
| NCT. Approach Bias Modification for the Treatment of Cannabis Use Disorder. *https://clinicaltrialsgov/ct2/show/NCT03629990* 2018. | Ineligible publication type |
| NCT. Substance Use Interventions for Truant Adolescents. *https://clinicaltrialsgov/show/NCT03655574* 2018. | Ineligible publication type |
| NCT. Neurobehavioral Measurement of Substance Users in Outpatient Treatment Setting. *https://clinicaltrialsgov/show/NCT03662529* 2018. | Ineligible publication type |
| NCT. Peer MI in FQHCs for Substance-using Emerging Adults. *https://clinicaltrialsgov/show/NCT03758131* 2018. | Ineligible publication type |
| NCT. UH3 Varenicline for Cannabis Use Disorder. *https://clinicaltrialsgov/ct2/show/NCT03980561* 2019. | Ineligible publication type |

| | |
|---|---|
| NCT. Personalized Feedback Intervention to Reduce Risky Cannabis Use. *https://clinicaltrialsgov/show/NCT04060602* 2019. | Ineligible publication type |
| NCT. Social Media Intervention for Cannabis Use in Emerging Adults. *https://clinicaltrialsgov/show/NCT04187989* 2019. | Ineligible publication type |
| NCT. Pilot Test of Parent-Focused Cannabis-Related Actions and Practices Intervention for Adolescent Marijuana Abuse. *https://clinicaltrialsgov/show/NCT04923230* 2021. | Ineligible publication type |
| NCT. Examining Effects of Domain Specific Episodic Future Thinking on Cannabis Use. *https://clinicaltrialsgov/show/NCT05324813* 2022. | Ineligible publication type |
| NCT. Investigating Two rTMS Strategies to Treat Cannabis Use Disorder. *https://clinicaltrialsgov/ct2/show/NCT05720312* 2023. | Ineligible publication type |
| Nizio P, Clausen B, Businelle MS, Ponton N, Jones AA, Redmond BY*, et al.* Mobile Intervention to Address Cannabis Use Disorder Among Black Adults: protocol for a Randomized Controlled Trial. *JMIR research protocols* 2024;**13**:e52776. https://doi.org/10.2196/52776 | Ineligible intervention |
| Notzon DP, Kelly MA, Choi CJ, Pavlicova M, Mahony AL, Brooks DJ*, et al.* Open-label pilot study of injectable naltrexone for cannabis dependence. *The American Journal of Drug and Alcohol Abuse* 2018;**44**:619-27. https://doi.org/10.1080/00952990.2017.1423321 | Ineligible study design |
| O'Farrell TJ, Murphy M, Alter J, Fals-Stewart W. Behavioral family counseling for substance abuse: a treatment development pilot study. *Addictive behaviors* 2010;**35**:1-6. https://doi.org/10.1016/j.addbeh.2009.07.003 | Ineligible population |
| Olthof MIA, Blankers M, van Laar MW, Goudriaan AE. ICan, an Internet-based intervention to reduce cannabis use: study protocol for a randomized controlled trial. *Trials* 2021;**22**:28. https://doi.org/10.1186/s13063-020-04962-3 | Ineligible publication type |
| Olthof MIA, Goudriaan AE, van Laar MW, Blankers M. A guided digital intervention to reduce cannabis use: the ICan randomized controlled trial. *Addiction* 2023;**118**:1775-86. https://doi.org/10.1111/add.16217 | Ineligible population |
| Østergård OK, Del Palacio-Gonzalez A, Nilsson KK, Pedersen MU. The Partners for Change Outcome Management System in the psychotherapeutic treatment of cannabis use: a pilot effectiveness randomized clinical trial. *Nordic journal of psychiatry* 2021;**75**:633-40. https://doi.org/10.1080/08039488.2021.1921265 | Ineligible population |
| Padovano HT, Miranda R, Jr. Using ecological momentary assessment to identify mechanisms of change: An application from a pharmacotherapy trial with adolescent cannabis users. *Journal of Studies on Alcohol and Drugs* 2018;**79**:190-8. https://doi.org/10.15288/jsad.2018.79.190 | Ineligible intervention |
| Palfai TP, Saitz R, Winter M, Brown TA, Kypri K, Goodness TM*, et al.* Web-based screening and brief intervention for student marijuana use in a university health center: pilot study to examine the implementation of eCHECKUP TO GO in different contexts. *Addictive behaviors* 2014;**39**:1346-52. https://doi.org/10.1016/j.addbeh.2014.04.025 | Ineligible population |
| Palfai TP, Tahaney K, Winter M, Saitz R. Readiness-to-change as a moderator of a web-based brief intervention for marijuana among students identified by health center screening. *Drug and alcohol dependence* 2016;**161**:368-71. https://doi.org/10.1016/j.drugalcdep.2016.01.027 | Ineligible population |
| Papinczak ZE, Connor JP, Feeney GFX, Gullo MJ. Additive effectiveness and feasibility of a theory-driven instant assessment and feedback system in brief cannabis intervention: a randomised controlled trial. *Addictive behaviors* 2021;**113**:106690. https://doi.org/10.1016/j.addbeh.2020.106690 | Ineligible population |
| Penetar DM, Looby AR, Ryan ET, Maywalt MA, Lukas SE. Bupropion reduces some of the symptoms of marihuana withdrawal in chronic marihuana users: a pilot study. *Substance abuse: research and treatment* 2012;**6**:63-71. https://doi.org/10.4137/SART.S9706 | Ineligible intervention |
| Peters EN, Leeman RF, Fucito LM, Toll BA, Corbin WR, O'Malley SS. Co-occurring marijuana use is associated with medication nonadherence and nonplanning impulsivity in young adult heavy drinkers. *Addictive behaviors* 2012;**37**:420-6. https://doi.org/10.1016/j.addbeh.2011.11.036 | Ineligible population |
| Peters EN, Nich C, Carroll KM. Primary outcomes in two randomized controlled trials of treatments for cannabis use disorders. *Drug and alcohol dependence* 2011;**118**:408-16. https://doi.org/10.1016/j.drugalcdep.2011.04.021 | Ineligible study design |
| Peters EN, Petry NM, Lapaglia DM, Reynolds B, Carroll KM. Delay discounting in adults receiving treatment for marijuana dependence. *Experimental and clinical psychopharmacology* 2013;**21**:46-54. https://doi.org/10.1037/a0030943 | Ineligible outcome |
| Peterson PL, Baer JS, Wells EA, Ginzler JA, Garrett SB. Short-term effects of a brief motivational intervention to reduce alcohol and drug risk among homeless adolescents. *Psychology of addictive behaviors* 2006;**20**:254-64. https://doi.org/10.1037/0893-164X.20.3.254 | Ineligible population |
| Phan O, Jouanne C, Monge S. A random clinical trial concerning the psychotherapy of adolescents addicted to cannabis. *Annales medico-psychologiques* 2010;**168**:145-51. https://doi.org/10.1016/j.amp.2009.12.013 | Ineligible publication type |
| Piehler TF, Winters KC. Parental involvement in brief interventions for adolescent marijuana use. *Psychology of addictive behaviors* 2015;**29**:512-21. https://doi.org/10.1037/adb0000106 | Ineligible population |

| | |
|---|---|
| Piehler TF, Winters KC. Decision-making style and response to parental involvement in brief interventions for adolescent substance use. *Journal of family psychology* 2017;**31**:336-46. https://doi.org/10.1037/fam0000266 | Ineligible population |
| Prince MA, Collins RL, Wilson SD, Vincent PC. A preliminary test of a brief intervention to lessen young adults' cannabis use: episode-level smartphone data highlights the role of protective behavioral strategies and exercise. *Experimental and clinical psychopharmacology* 2020;**28**:150-6. https://doi.org/10.1037/pha0000301 | Ineligible population |
| Prince MA, Tyskiewicz AJ, Conner BT, Parnes JE, Shillington AM, George MW*, et al.* Mechanisms of change in an adapted marijuana e-CHECKUP TO GO intervention on decreased college student cannabis use. *Journal of substance abuse treatment* 2021;**124**:108308. https://doi.org/10.1016/j.jsat.2021.108308 | Ineligible population |
| Prisciandaro JJ, Mellick W, Squeglia LM, Hix S, Arnold L, Tolliver BK. Results from a randomized, double-blind, placebo-controlled, crossover, multimodal-MRI pilot study of gabapentin for co-occurring bipolar and cannabis use disorders. *Addiction biology* 2022;**27**:e13085. https://doi.org/10.1111/adb.13085 | Ineligible intervention |
| Prochaska JJ, Vogel EA, Chieng A, Kendra M, Baiocchi M, Pajarito S*, et al.* A Therapeutic Relational Agent for Reducing Problematic Substance Use (Woebot): development and Usability Study. *Journal of medical Internet research* 2021;**23**:e24850. https://doi.org/10.2196/24850 | Ineligible study design |
| Quinn CA, Walter ZC, de Andrade D, Dingle G, Haslam C, Hides L. Controlled Trial Examining the Strength-Based Grit Wellbeing and Self-Regulation Program for Young People in Residential Settings for Substance Use. *International journal of environmental research and public health* 2022;**19**. https://doi.org/10.3390/ijerph192113835 | Ineligible study design |
| Rabinovitz S, Nagar M. The effects of craving on implicit cognitive mechanisms involved in risk behavior: Can dialectical behavior therapy in therapeutic communities make a difference? A pilot study. *Therapeutic Communities* 2018;**39**:83-92. https://doi.org/10.1108/TC-12-2017-0034 | Ineligible study design |
| Riggs NR, Conner BT, Parnes JE, Prince MA, Shillington AM, George MW. Marijuana eCHECKUPTO GO: effects of a personalized feedback plus protective behavioral strategies intervention for heavy marijuana-using college students. *Drug and alcohol dependence* 2018;**190**:13-9. https://doi.org/10.1016/j.drugalcdep.2018.05.020 | Ineligible population |
| Riggs P. Want change? Try honey instead of vinegar. *Journal of the American Academy of Child & Adolescent Psychiatry* 2015;**54**:440-1. https://doi.org/10.1016/j.jaac.2015.03.015 | Ineligible publication type |
| Roffman RA, Klepsch R, Wertz JS, Simpson EE, Stephens RS. Predictors of attrition from an outpatient marijuana-dependence counseling program. *Addictive behaviors* 1993;**18**:553-66. https://doi.org/10.1016/0306-4603(93)90071-g | Ineligible study design |
| Roffman RA, Stephens RS, Simpson EE, Whitaker DL. Treatment of marijuana dependence: Preliminary results. *Special Issue: Marijuana--an update* 1988;**20**:129-37. https://doi.org/10.1080/02791072.1988.10524382 | Ineligible outcome[d] |
| Rooke SE, Gates PJ, Norberg MM, Copeland J. Applying technology to the treatment of cannabis use disorder: comparing telephone versus Internet delivery using data from two completed trials. *Journal of Substance Abuse Treatment* 2014;**46**:78-84. https://doi.org/10.1016/j.jsat.2013.08.007 | Ineligible study design |
| Roten A, Baker NL, Gray KM. Cognitive performance in a placebo-controlled pharmacotherapy trial for youth with marijuana dependence. *Addictive behaviors* 2015;**45**:119-23. https://doi.org/10.1016/j.addbeh.2015.01.013 | Ineligible intervention* |
| Roten AT, Baker NL, Gray KM. Marijuana craving trajectories in an adolescent marijuana cessation pharmacotherapy trial. *Addictive behaviors* 2013;**38**:1788-91. https://doi.org/10.1016/j.addbeh.2012.11.003 | Ineligible intervention* |
| Rounsaville DB, Hunkele K, Easton CJ, Nich C, Carroll KM. Making consent more informed: preliminary results from a multiple-choice test among probation-referred marijuana users entering a randomized clinical trial. *Journal of the American Academy of Psychiatry and the Law* 2008;**36**:354-9. | Ineligible study design |
| Ryan SR, Stanger C, Thostenson J, Whitmore JJ, Budney AJ. The impact of disruptive behavior disorder on substance use treatment outcome in adolescents. *Journal of substance abuse treatment* 2013;**44**:506-14. https://doi.org/10.1016/j.jsat.2012.11.003 | Ineligible population |
| Sahlem GL, Kim B, Baker NL, Wong BL, Caruso MA, Campbell LA*, et al.* A preliminary randomized controlled trial of repetitive transcranial magnetic stimulation applied to the left dorsolateral prefrontal cortex in treatment seeking participants with cannabis use disorder. *Drug and alcohol dependence* 2024;**254**:111035. https://doi.org/10.1016/j.drugalcdep.2023.111035 | Ineligible intervention |
| Sahlem GL, Kim B, Baker NL, Wong BL, Caruso MA, Campbell LA*, et al.* A Preliminary Investigation Of Repetitive Transcranial Magnetic Stimulation Applied To The Left Dorsolateral Prefrontal Cortex In Treatment Seeking Participants With Cannabis Use Disorder. *medRxiv* 2023;**12**:12. https://doi.org/10.1101/2023.07.10.23292461 | Ineligible intervention |
| Santisteban DA, Mena MP, McCabe BE. Preliminary results for an adaptive family treatment for drug abuse in Hispanic youth. *Journal of family psychology* 2011;**25**:610-4. https://doi.org/10.1037/a0024016 | Ineligible population |

| | |
|---|---|
| Satre DD, Leibowitz A, Sterling SA, Lu Y, Travis A, Weisner C. A randomized clinical trial of Motivational Interviewing to reduce alcohol and drug use among patients with depression. *Journal of consulting and clinical psychology* 2016;**84**:571-9. https://doi.org/10.1037/ccp0000096 | Ineligible population |
| Satre DD, Parthasarathy S, Young-Wolff KC, Meacham MC, Borsari B, Hirschtritt ME*, et al.* Cost-Effectiveness of Motivational Interviewing to Reduce Alcohol and Cannabis Use Among Patients With Depression. *Journal of studies on alcohol and drugs* 2022;**83**:662-71. https://doi.org/10.15288/jsad.21-00186 | Ineligible population |
| Schaub MP, Haug S, Wenger A, Berg O, Sullivan R, Beck T*, et al.* Can reduce--the effects of chat-counseling and web-based self-help, web-based self-help alone and a waiting list control program on cannabis use in problematic cannabis users: a randomized controlled trial. *BMC Psychiatry* 2013;**13**:305. https://doi.org/10.1186/1471-244X-13-305 | Ineligible publication type |
| Schaub MP, Henderson CE, Pelc I, Tossmann P, Phan O, Hendriks V*, et al.* Multidimensional family therapy decreases the rate of externalising behavioural disorder symptoms in cannabis abusing adolescents: outcomes of the INCANT trial. *BMC Psychiatry* 2014;**14**:26. https://doi.org/10.1186/1471-244X-14-26 | Ineligible outcome |
| Schaub MP, Wenger A, Berg O, Beck T, Stark L, Buehler E*, et al.* A Web-Based Self-Help Intervention With and Without Chat Counseling to Reduce Cannabis Use in Problematic Cannabis Users: three-Arm Randomized Controlled Trial. *Journal of medical Internet research* 2015;**17**:e232. https://doi.org/10.2196/jmir.4860 | Ineligible intervention |
| Schell C, Godinho A, Cunningham JA. Examining the influence of rurality on frequency of cannabis use and severity of consequences as moderated by age and gender. *Addictive behaviors* 2022;**133**:107385. https://doi.org/10.1016/j.addbeh.2022.107385 | Ineligible study design |
| Schluter MG, Hodgins DC, Stea JN, Kilborn ML. Promoting self-change in cannabis use disorder: findings from a randomized trial. *Frontiers in psychiatry* 2022;**13**:1015443. https://doi.org/10.3389/fpsyt.2022.1015443 | Ineligible intervention |
| Schneegans A, Bourgognon F, Albuisson E, Schwan R, Arfa M, Polli L*, et al.* Mindfulness-based relapse prevention for cannabis regular users: preliminary outcomes of a randomized clinical trial. *Encephale* 2022;**48**:241-6. https://doi.org/10.1016/j.encep.2021.02.015 | Ineligible population |
| Schwartz RP, Gryczynski J, Mitchell SG, Gonzales A, Moseley A, Peterson TR*, et al.* Computerized versus in-person brief intervention for drug misuse: a randomized clinical trial. *Addiction* 2014;**109**:1091-8. https://doi.org/10.1111/add.12502 | Ineligible population |
| Scott CK, Dennis ML, Grella CE, Watson DP, Davis JP, Hart MK. Using recovery management checkups for primary care to improve linkage to alcohol and other drug use treatment: a randomized controlled trial three month findings. *Addiction* 2023;**118**:520-32. https://doi.org/10.1111/add.16064 | Ineligible population |
| Scott CK, Dennis ML, Grella CE, Watson DP, Davis JP, Hart MK. A randomized controlled trial of recovery management checkups for primary care patients: twelve-month results. *Alcohol, clinical & experimental research* 2023;**47**:1964-77. https://doi.org/10.1111/acer.15172 | Ineligible population |
| Sharma K, Ghosh A, Krishnan NC, Kathirvel S, Basu D, Kumar A*, et al.* Digital screening and brief intervention for illicit drug misuse in college students: a mixed methods, pilot, cluster, randomized trial from India. *Asian journal of psychiatry* 2023;**81**:103432. https://doi.org/10.1016/j.ajp.2022.103432 | Ineligible population |
| Shekhawat AS, Mathur R, Sarkar S, Kaloiya GS, Balhara YPS. A randomized controlled trial of brief intervention for patients with cannabis use disorder. *Journal of neurosciences in rural practice* 2023;**14**:710-6. https://doi.org/10.25259/JNRP_79_2023 | Ineligible intervention |
| Sherman BJ, Baker NL, McRae-Clark AL. Gender differences in cannabis use disorder treatment: Change readiness and taking steps predict worse cannabis outcomes for women. *Addictive Behaviors* 2016;**60**:197-202. https://doi.org/10.1016/j.addbeh.2016.04.014 | Ineligible study design |
| Sherman BJ, Baker NL, McRae-Clark AL. Effect of oxytocin pretreatment on cannabis outcomes in a brief motivational intervention. *Psychiatry research* 2017;**249**:318-20. https://doi.org/10.1016/j.psychres.2017.01.027 | Ineligible intervention |
| Sherman BJ, Baker NL, Schmarder KM, McRae-Clark AL, Gray KM. Latency to cannabis dependence mediates the relationship between age at cannabis use initiation and cannabis use outcomes during treatment in men but not women. *Drug and alcohol dependence* 2021;**218**:108383. https://doi.org/10.1016/j.drugalcdep.2020.108383 | Ineligible study design |
| Sherman BJ, McRae-Clark AL, Baker NL, Sonne SC, Killeen TK, Cloud K*, et al.* Gender differences among treatment-seeking adults with cannabis use disorder: clinical profiles of women and men enrolled in the achieving cannabis cessation-evaluating N-acetylcysteine treatment (ACCENT) study. *The American journal on addictions* 2017;**26**:136-44. https://doi.org/10.1111/ajad.12503 | Ineligible study design |
| Sherman BJ, Sofis MJ, Borodovsky JT, Gray KM, McRae-Clark AL, Budney AJ. Evaluating cannabis use risk reduction as an alternative clinical outcome for cannabis use disorder. *Psychology of addictive behaviors* 2022;**36**:505-14. https://doi.org/10.1037/adb0000760 | Ineligible study design |
| Short NA, Zvolensky MJ, Schmidt NB. A pilot randomized clinical trial of Brief Behavioral Treatment for Insomnia to reduce problematic cannabis use among trauma-exposed young adults. *Journal of substance abuse treatment* 2021;**131**:108537. https://doi.org/10.1016/j.jsat.2021.108537 | Ineligible population |

| | |
|---|---|
| Shrier LA, Burke PJ, Kells M, Scherer EA, Sarda V, Jonestrask C, *et al.* Pilot randomized trial of MOMENT, a motivational counseling-plus-ecological momentary intervention to reduce marijuana use in youth. *mHealth* 2018;**4**:29. https://doi.org/10.21037/mhealth.2018.07.04 | Ineligible intervention |
| Shrier LA, Harris SK. Associations of momentary mindfulness with affect and cannabis desire in a trial of cannabis use interventions with and without momentary assessment. *Journal of Adolescent Health* 2023;**72**:126-9. https://doi.org/10.1016/j.jadohealth.2022.09.002 | Ineligible intervention |
| Sinadinovic K, Johansson M, Johansson AS, Lundqvist T, Lindner P, Hermansson U. Guided web-based treatment program for reducing cannabis use: a randomized controlled trial. *Addiction science & clinical practice* 2020;**15**:9. https://doi.org/10.1186/s13722-020-00185-8 | Ineligible intervention |
| Sinha R, Easton C, Renee-Aubin L, Carroll KM. Engaging young probation-referred marijuana-abusing individuals in treatment: a pilot trial. *The American journal on addictions* 2003;**12**:314-23. | Ineligible intervention |
| Smith JW, Schmeling G, Knowles PL. A marijuana smoking cessation clinical trial utilizing THC-free marijuana, aversion therapy, and self-management counseling. *Journal of Substance Abuse Treatment* 1988;**5**:89-98. https://doi.org/10.1016/0740-5472(88)90018-9 | Ineligible study design |
| Sofis MJ, Lemley SM, Jacobson NC, Budney AJ. Initial evaluation of domain-specific episodic future thinking on delay discounting and cannabis use. *Experimental and Clinical Psychopharmacology* 2022;**30**:918-27. https://doi.org/10.1037/pha0000501 | Ineligible intervention |
| Spillane NS, Schick MR, Hostetler KL, Trinh CD, Kahler CW. Results of a pilot study examining the effect of positive psychology interventions on cannabis use and related consequences. *Contemporary clinical trials* 2023;**131**:107247. https://doi.org/10.1016/j.cct.2023.107247 | Ineligible population |
| Spring B, Ferguson MJ. CALM technology-supported intervention: synopsis of evidence for an emerging class of practice tool. *Translational behavioral medicine* 2011;**1**:8-9. https://doi.org/10.1007/s13142-011-0031-5 | Ineligible publication type |
| Squeglia LM, Baker NL, McClure EA, Tomko RL, Adisetiyo V, Gray KM. Alcohol use during a trial of N-acetylcysteine for adolescent marijuana cessation. *Addictive Behaviors* 2016;**63**:172-7. https://doi.org/10.1016/j.addbeh.2016.08.001 | Ineligible outcome |
| Stanger C, Ryan SR, Scherer EA, Norton GE, Budney AJ. Clinic- and home-based contingency management plus parent training for adolescent cannabis use disorders. *Journal of the American Academy of Child and Adolescent Psychiatry* 2015;**54**:445-53.e2. https://doi.org/10.1016/j.jaac.2015.02.009 | Ineligible population |
| Stanger C, Scherer EA, Vo HT, Babbin SF, Knapp AA, McKay JR, *et al.* Working memory training and high magnitude incentives for youth cannabis use: a SMART pilot trial. *Psychology of addictive behaviors* 2020;**34**:31-9. https://doi.org/10.1037/adb0000480 | Ineligible intervention |
| Stein LA, Clair M, Lebeau R, Colby SM, Barnett NP, Golembeske C, *et al.* Motivational interviewing to reduce substance-related consequences: effects for incarcerated adolescents with depressed mood. *Drug and alcohol dependence* 2011;**118**:475-8. https://doi.org/10.1016/j.drugalcdep.2011.03.023 | Ineligible population |
| Stein LA, Lebeau R, Colby SM, Barnett NP, Golembeske C, Monti PM. Motivational interviewing for incarcerated adolescents: effects of depressive symptoms on reducing alcohol and marijuana use after release. *Journal of studies on alcohol and drugs* 2011;**72**:497-506. https://doi.org/10.15288/jsad.2011.72.497 | Ineligible population |
| Stein LAR, Martin R, Clair-Michaud M, Lebeau R, Hurlbut W, Kahler CW, *et al.* A randomized clinical trial of motivational interviewing plus skills training vs. Relaxation plus education and 12-Steps for substance using incarcerated youth: effects on alcohol, marijuana and crimes of aggression. *Drug and alcohol dependence* 2020;**207**:107774. https://doi.org/10.1016/j.drugalcdep.2019.107774 | Ineligible population |
| Stephens RS, Walker R, DeMarce J, Lozano BE, Rowland J, Walker D, *et al.* Treating cannabis use disorder: exploring a treatment as needed model with 34-month follow-up. *Journal of substance abuse treatment* 2020;**117**:108088. https://doi.org/10.1016/j.jsat.2020.108088 | Ineligible intervention |
| Stephens RS, Wertz JS, Roffman RA. Self-efficacy and marijuana cessation: a construct validity analysis. *Journal of consulting and clinical psychology* 1995;**63**:1022-31. https://doi.org/10.1037//0022-006x.63.6.1022 | Ineligible study design |
| Sugarman DE, De Aquino JP, Poling J, Sofuoglu M. Feasibility and effects of galantamine on cognition in humans with cannabis use disorder. *Pharmacology, biochemistry, and behavior* 2019;**181**:86-92. https://doi.org/10.1016/j.pbb.2019.05.004 | Ineligible intervention |
| Sweeney MM, Rass O, DiClemente C, Schacht RL, Vo HT, Fishman MJ, *et al.* Working memory training for adolescents with cannabis use disorders: A randomized controlled trial. *Journal of Child & Adolescent Substance Abuse* 2018;**27**:211-26. https://doi.org/10.1080/1067828X.2018.1451793 | Ineligible intervention |
| The Marijuana Treatment Project Research G. Brief treatments for cannabis dependence: Findings from a randomized multisite trial. *Addictive behaviors: New readings on etiology, prevention, and treatment* 2009; https://doi.org/10.1037/11855-017 | Ineligible publication type |
| Thornton CC, Patkar AA, Murray HW, Mannelli P, Gottheil E, Vergare MJ, *et al.* High- and low-structure treatments for substance dependence: role of learned helplessness. *American journal of drug and alcohol abuse* 2003;**29**:567-84. https://doi.org/10.1081/ada-120023459 | Ineligible population |

| | |
|---|---|
| Timko C, Debenedetti A, Billow R. Intensive referral to 12-Step self-help groups and 6-month substance use disorder outcomes. *Addiction* 2006;**101**:678-88. https://doi.org/10.1111/j.1360-0443.2006.01391.x | Ineligible population |
| Tolou-Shams M, Dauria E, Conrad SM, Kemp K, Johnson S, Brown LK. Outcomes of a family-based HIV prevention intervention for substance using juvenile offenders. *Journal of substance abuse treatment* 2017;**77**:115-25. https://doi.org/10.1016/j.jsat.2017.03.013 | Ineligible population |
| Tomko RL, Baker NL, Hood CO, Gilmore AK, McClure EA, Squeglia LM*, et al.* Depressive symptoms and cannabis use in a placebo-controlled trial of N-Acetylcysteine for adult cannabis use disorder. *Psychopharmacology* 2020;**237**:479-90. https://doi.org/10.1007/s00213-019-05384-z | Ineligible outcome |
| Tomko RL, Baker NL, McClure EA, Sonne SC, McRae-Clark AL, Sherman BJ*, et al.* Incremental validity of estimated cannabis grams as a predictor of problems and cannabinoid biomarkers: evidence from a clinical trial. *Drug and alcohol dependence* 2018;**182**:1-7. https://doi.org/10.1016/j.drugalcdep.2017.09.035 | Ineligible study design |
| Tomko RL, Wolf BJ, McClure EA, Carpenter MJ, Magruder KM, Squeglia LM*, et al.* Who Responds to a Multi-Component Treatment for Cannabis Use Disorder? Using Multivariable and Machine Learning Models to Classify Treatment Responders and Non-Responders. *Addiction* 2023;**118**:1965-74. https://doi.org/10.1111/add.16226 | Ineligible study design |
| Tossmann H-P, Jonas B, Tensil M-D, Lang P, Struber E. A controlled trial of an internet-based intervention program for cannabis users. *Cyberpsychology, Behavior, and Social Networking* 2011;**14**:673-9. https://doi.org/10.1089/cyber.2010.0506 | Ineligible intervention |
| Tossmann P, Jonas B, Rigter H, Gantner A. Treating adolescents with cannabis use disorder with Multidimensional Family Therapy (MDFT): main results of a Randomized Controlled Trial (RCT). *Sucht* 2012;**58**:157-66. https://doi.org/10.1024/0939-5911.a000180 | Ineligible outcome[c] |
| Treloar Padovano H, Miranda R. Using Ecological Momentary Assessment to Identify Mechanisms of Change: an Application From a Pharmacotherapy Trial With Adolescent Cannabis Users. *Journal of studies on alcohol and drugs* 2018;**79**:190-8. https://doi.org/10.15288/jsad.2018.79.190 | Ineligible intervention |
| Trigo JM, Lagzdins D, Rehm J, Selby P, Gamaleddin I, Fischer B*, et al.* Effects of fixed or self-titrated dosages of Sativex on cannabis withdrawal and cravings. *Drug and alcohol dependence* 2016;**161**:298-306. https://doi.org/10.1016/j.drugalcdep.2016.02.020 | Ineligible intervention |
| Trigo JM, Soliman A, Quilty LC, Fischer B, Rehm J, Selby P*, et al.* Nabiximols combined with motivational enhancement/cognitive behavioral therapy for the treatment of cannabis dependence: a pilot randomized clinical trial. *PloS one* 2018;**13**:e0190768. https://doi.org/10.1371/journal.pone.0190768 | Ineligible intervention* |
| Ullrich HS, Torbati A, Fan W, Arbona C, Cano MA, Essa S*, et al.* Race, psychosocial characteristics, and treatment outcomes among individuals undergoing treatment for cannabis use disorder: a latent profile analysis based on preferred method of using cannabis. *Journal of substance abuse treatment* 2021;**131**:108561. https://doi.org/10.1016/j.jsat.2021.108561 | Ineligible study design |
| van der Baan HS, D'Escury-Koenigs ALC, Wiers RW. The effectiveness of cognitive bias modification in reducing substance use in detained juveniles: An RCT. *Journal of Behavior Therapy and Experimental Psychiatry* 2024;**82**:1-9. https://doi.org/10.1016/j.jbtep.2023.101916 | Ineligible population |
| van der Pol TM, Henderson CE, Hendriks V, Schaub MP, Rigter H. Multidimensional Family Therapy Reduces Self-Reported Criminality Among Adolescents With a Cannabis Use Disorder. *International journal of offender therapy and comparative criminology* 2018;**62**:1573-88. https://doi.org/10.1177/0306624X16687536 | Ineligible outcome |
| van der Pol TM, Hendriks V, Rigter H, Cohn MD, Doreleijers TAH, van Domburgh L. Multidimensional family therapy in adolescents with a cannabis use disorder: long-term effects on delinquency in a randomized controlled trial. *Child Adolescent Psychiatry Mental Health* 2018;**12**:44. https://doi.org/10.1186/s13034-018-0248-x. | Ineligible outcome |
| Vendetti J, McRee B, Miller M, Christiansen K, Herrell J. Correlates of pre-treatment drop-out among persons with marijuana dependence. *Addiction* 2002;**97**:125-34. https://doi.org/10.1046/j.1360-0443.97.s01.8.x | Ineligible study design |
| Waldron HB, Slesnick N, Brody JL, Turner CW, Peterson TR. Treatment outcomes for adolescent substance abuse at 4- and 7-month assessments. *Journal of consulting and clinical psychology* 2001;**69**:802-13. | Ineligible population |
| Waldron HB, Turner CW, Ozechowski TJ. Profiles of drug use behavior change for adolescents in treatment. *Addictive behaviors* 2005;**30**:1775-96. https://doi.org/10.1016/j.addbeh.2005.07.001 | Ineligible study design |
| Walker DD, Stephens RS, Towe S, Banes K, Roffman R. Maintenance Check-ups Following Treatment for Cannabis Dependence. *Journal of substance abuse treatment* 2015;**56**:11-5. https://doi.org/10.1016/j.jsat.2015.03.006 | Ineligible intervention |
| Waller R, Bonar EE, Fernandez AC, Walton MA, Chermack ST, Cunningham RM*, et al.* Exploring the components of an efficacious computer brief intervention for reducing marijuana use among adults in the emergency department. *Journal of substance abuse treatment* 2019;**99**:67-72. https://doi.org/10.1016/j.jsat.2019.01.014 | Ineligible study design |

| | |
|---|---|
| Walton MA, Bohnert K, Resko S, Barry KL, Chermack ST, Zucker RA*, et al.* Computer and therapist based brief interventions among cannabis-using adolescents presenting to primary care: one year outcomes. *Drug and alcohol dependence* 2013;**132**:646-53. https://doi.org/10.1016/j.drugalcdep.2013.04.020 | Ineligible population |
| Walukevich-Dienst K, Lewis EM, Neighbors C, Green JC, Buckner JD. Online personalized feedback intervention reduces cannabis-related problems among college students with high problem distress. *Experimental and clinical psychopharmacology* 2021;**29**:14-22. https://doi.org/10.1037/pha0000361 | Ineligible intervention |
| Walukevich-Dienst K, Neighbors C, Buckner JD. Online personalized feedback intervention for cannabis-using college students reduces cannabis-related problems among women. *Addictive behaviors* 2019;**98**:106040. https://doi.org/10.1016/j.addbeh.2019.106040 | Ineligible intervention |
| Wang R, Trigo JM, Foll BL. Effects of sub-chronic nabiximols on biological markers of individuals undergoing a clinical trial for the treatment of cannabis use disorder. *American journal of translational research* 2023;**15**:5228-38. | Ineligible outcome |
| Wanmaker S, Leijdesdorff SMJ, Geraerts E, van de Wetering BJM, Renkema PJ, Franken IHA. The efficacy of a working memory training in substance use patients: a randomized double-blind placebo-controlled clinical trial. *Journal of clinical and experimental neuropsychology* 2018;**40**:473-86. https://doi.org/10.1080/13803395.2017.1372367 | Ineligible population |
| Ward CL, Mertens JR, Bresick GF, Little F, Weisner CM. Screening and brief intervention for substance misuse: does it reduce aggression and HIV-related risk behaviours? *Alcohol and alcoholism (Oxford, Oxfordshire)* 2015;**50**:302-9. https://doi.org/10.1093/alcalc/agv007 | Ineligible population |
| Weinstein AM, Miller H, Bluvstein I, Rapoport E, Schreiber S, Bar-Hamburger R*, et al.* Treatment of cannabis dependence using escitalopram in combination with cognitive-behavior therapy: a double-blind placebo-controlled study. *American journal of drug and alcohol abuse* 2014;**40**:16-22. https://doi.org/10.3109/00952990.2013.819362 | Ineligible intervention* |
| Wesley MC, Minatrea NB, Watson JC. Animal-assisted therapy in the treatment of substance dependence. *Anthrozoos* 2009;**22**:137-48. https://doi.org/10.2752/175303709X434167 | Ineligible population |
| Weymann N, Baldus C, Miranda A, More K, Reis O, Thomasius R. Trainer effects in a group training for young cannabis consumers - Results of the multisite trial "CAN Stop." [*Trainereffekte in einem gruppentraining fur junge cannabiskonsumenten Ergebnisse der Multicenterstudie "CAN Stop"]. Sucht* 2011;**57**:193-202. https://doi.org/10.1024/0939-5911.a000106 | Ineligible study design |
| White HR, Morgan TJ, Pugh LA, Celinska K, Labouvie EW, Pandina RJ. Evaluating two brief substance-use interventions for mandated college students. *Journal of studies on alcohol* 2006;**67**:309-17. https://doi.org/10.15288/jsa.2006.67.309 | Ineligible population |
| White HR, Mun EY, Pugh L, Morgan TJ. Long-term effects of brief substance use interventions for mandated college students: sleeper effects of an in-person personal feedback intervention. *Alcoholism, clinical and experimental research* 2007;**31**:1380-91. https://doi.org/10.1111/j.1530-0277.2007.00435.x | Ineligible population |
| Winters KC, Stinchfield RD, Opland E, Weller C, Latimer WW. The effectiveness of the minnesota model approach in the treatment of adolescent drug abusers. *Addiction* 2000;**95**:601-12. https://doi.org/10.1046/j.1360-0443.2000.95460111.x | Ineligible study design |
| Wolff J, Esposito-Smythers C, Frazier E, Stout R, Gomez J, Massing-Schaffer M*, et al.* A randomized trial of an integrated cognitive behavioral treatment protocol for adolescents receiving home-based services for co-occurring disorders. *Journal of substance abuse treatment* 2020;**116**:108055. https://doi.org/10.1016/j.jsat.2020.108055 | Ineligible population |
| Worley MJ, Tate SR, Brown SA. Mediational relations between 12-Step attendance, depression and substance use in patients with comorbid substance dependence and major depression. *Addiction* 2012;**107**:1974-83. https://doi.org/10.1111/j.1360-0443.2012.03943.x | Ineligible study design |
| Xu X, Yonkers KA, Ruger JP. Economic evaluation of a behavioral intervention versus brief advice for substance use treatment in pregnant women: results from a randomized controlled trial. *BMC pregnancy and childbirth* 2017;**17**:83. https://doi.org/10.1186/s12884-017-1260-5 | Ineligible population |
| Yamada C, Siste K, Hanafi E, Ophinni Y, Beatrice E, Rafelia V*, et al.* Relapse prevention group therapy via video-conferencing for substance use disorder: protocol for a multicentre randomised controlled trial in Indonesia. *BMJ open* 2021;**11**:e050259. https://doi.org/10.1136/bmjopen-2021-050259 | Ineligible publication type |
| Yurasek AM, Dennhardt AA, Murphy JG. A randomized controlled trial of a behavioral economic intervention for alcohol and marijuana use. *Experimental and Clinical Psychopharmacology* 2015;**23**:332-8. https://doi.org/10.1037/pha0000025 | Ineligible population |

Listed reason for exclusion is the first criterion met in the following hierarchy: ineligible publication type, study design, population, intervention, setting, outcome, and duplicate.

* Study of pharmacological interventions in which all arms received an identical adjunct psychosocial intervention.

[a] Secondary paper linked to included study (Buckner 2019). Not used in extraction or risk of bias assessments.

[b] Secondary paper linked to included study (Stephens 2000). Not used in extraction or risk of bias assessments.

[c] Secondary paper linked to included study (Rigter 2013). Not used in extraction or risk of bias assessments.

[d] Secondary paper linked to included study (Stephens 1994). Not used in extraction or risk of bias assessments.
