## Supplementary material for "Effectiveness and safety of psychosocial interventions for the treatment of cannabis use disorder: a systematic review and meta-analysis": SuppInfo08

### SUPPORTING INFORMATION 8. PROGRESS-PLUS CHARACTERISTICS

Numbering of tables is specific to this Supporting Information document. References relating to this Supporting Information are included at the end of this document.

Table 1 presents characteristics of participants in the included studies that are considered to stratify health opportunities and outcomes.[1] We also present a narrative summary of the relevant characteristics.

The mean age of participants in the included studies ranged from 16 years[2-4] to 48 years.[5] Five studies specifically focused on younger participants, only recruiting people aged 25 years or under.[2-4,6,7]

Data were predominantly from the United States (15 studies).[2,4-17] Two studies were conducted in Iran,[18,19] and one study was conducted in each of Turkey,[20] Australia,[21] Germany[22] and Pakistan.[23] The final study was a multi-site European trial, conducted in Belgium, France, Germany, Switzerland and the Netherlands.[3] Place of residence was not well described in the studies, but several authors reported that those with difficulties attending the treatment site were excluded, including those with excessive commuting distance, transportation difficulties, or unstable living situations.[11,13-15,23] Consequently, those in more rural areas, with poorer transport links, and people living with homelessness or in temporary housing may have been excluded from these studies.

For five studies, almost all participants were White (90-100%).[4,10,11,16,17] Although not reported, this may also be the case for Copeland 2001,[21] where an Australian cohort were recruited, but only the proportion of Aboriginal or Torres Strait Islanders was given (3.1%). Most of the remaining studies recruited a more diverse sample of participants, but still with a majority of White people, ranging from 51% to 71% of participants, with only 12% to 23.6% Black participants.[2,5,8,9,13-15] Two studies reported the inclusion of 60-64% Black participants.[6,12] Wolitzky-Taylor 2022[7] included a more diverse range of ethnicities, including 25% Hispanic and 17% Asian participants. Six studies did not provide information on race/ethnicity of participants,[3,18-20,22,23] but three of these did report some language restrictions – limiting participation to those who spoke the local language.[3,22,23]

Twelve studies reported that the majority of participants were working, on either a full-time or part-time basis.[8-17,21,23] Of the remaining studies, three did not provide information on employment status, but specifically enrolled young people, aged ≤18.[2-4] Two included a majority of participants who were unemployed (51% in Carroll 2006, 85% in Budak 2024).[6,20] Hoch 2014[22] indicated that 33.3% of participants were employed and 19.6% were at school, but did not account for the remaining participants. Four studies did not provide any information on employment status,[5,7,18,19] although one of these only recruited veterans.[5]

All of the included studies recruited a majority of male participants, with an average of 80.4% males across the studies. This ranged from a minimum of 56.4% males[9] to 100% males.[18-20] One study reported pregnancy as an exclusion criterion.[5]

Education was reported differently across the studies. Seven studies indicated the mean number of years of education for participants, which was 13.5 years across the studies.[8,10,11,13-16] Ten further studies provided a breakdown of educational attainment across participants.[6,9,12,17-23] They predominantly included a mix of participants, some of whom had completed secondary school only, some with college education, and some with university degrees. A small number of studies required participants to have specific educational attainment for participation, including either a reading age at fifth-grade level,[2,13,14] English-literacy[21] or an IQ ≥70.[3]

Only eight studies reported information on socioeconomic status. Four provided information on income; either monthly income (mean $906 USD in Budney 2000, and $1294 USD in Budney 2006),[10,11] or annual income (mean $20,220 USD in Stephens 1994, 75% with income ≥ $10,000 USD in Litt 2020).[15,17] Three studies provided information on socioeconomic status, including Hoch 2014[22] (described as predominantly lower/lower-middle social class), Khalily 2023[23] (predominantly middle social class), and Stanger 2009[4] (average socioeconomic status of 7.05 on the Hollingshead scale, corresponding to roles such as teachers and administrators). Two studies gave information on home ownership (46% in Stephens 1994, 47% in Babor 2004).[8,17]

Ten studies reported on the number of participants who were married or co-habiting; on average across the studies this was 36% of participants.[8,13-17,20-23] Four studies reported on the percentage of participants who were never married, which ranged from 57.5 to 96%.[6,10-12] Two studies in younger participants reported on the percentage who were living with family (89.7% in Rigter 2013, 100% in Stanger 2009).[3,4]

Fourteen studies excluded individuals with serious mental health issues, including psychosis or suicidal ideation.[2,4-7,9-11,13,16-18,20,22] One study excluded any participant with any DSM-IV Axis 1 (mental health) conditions, unless they were sufficiently mild as to not interfere with treatment.[21] Eight excluded participants who required inpatient treatment for a condition, or with other serious medical problems.[2,3,6,10-13,15] Three studies specifically included participants with certain psychological features, including people with features of anxiety of depression[9,19] or more negative affect.[7]

Finally, although several studies included participants involved with or referred by the legal system, four studies excluded those in whom incarceration was imminent,[8,10-12] and one excluded participant who had any legal problems.[13]

**Table 1.** PROGRESS-Plus characteristics

| Study ID | Age (years) | Place of residence | Race, ethnicity, culture, language | Occupation | Gender/sex (% male) | Education | Socio-economic Status | Social capital | Other PROGRESS details |
|---|---|---|---|---|---|---|---|---|---|
| Babor 2004[8] | M=36.45 (SD=8.45), range 18-62 | United States | 71% White, 16% Hispanic, 12% African American, 1% Other | 71% employed full-time, 13% employed part-time, 12% unemployed, 4% student/retired/ homemaker | 71 | M=14.28 years of education (SD=2.30) | 47% own residence, 51% rent residence, 2% live in room/ shelter | 60% not married; Excluded those who were unable to provide a contact person | Legal issues: Excluded people in whom legal status might have interfered with treatment (e.g., mandated treatment, pending jail sentencing) |
| Buckner 2019[9] | M=23.15 (SD=7.38), range 18-65 [inclusion criterion] | United States | 63.6% Non-Hispanic White, 23.6% Non-Hispanic African American, 10.9% Hispanic White, 1.8% Multiracial | 21.8% employed full-time, 47.2% employed part-time | 56.4 | 14.5% high school education, 63.6% some college education, 5.5% technical degree, 16.4% Bachelor's degree | NR | NR | Mental health: Included only those who met DSM-5 criteria for an anxiety disorder. Excluded those with psychiatric disorders that precluded participation (e.g. psychosis) and those with severe suicidal ideation |
| Budak 2024[20] | Mean NR<br>51.6% aged 18-28; 35% aged 29-39; 13.3% aged 40-50 | Turkey | NR | 15% working, 85% not working | 100 | 8.3% literate, 25% primary education, 38.3% secondary education, 28.3% university education | NR | 38.3% married, 61.7% single; 11.6% living alone, 88.3% living with family; 13.33% had children | Mental health: Excluded those with comorbid psychiatric diagnoses, and those with 'communication problems' |
| Budney 2000[10] | M=32.85 (SD=8.52), range ≥18 [inclusion criterion] | United States | 100% Caucasian | 65% employed full time | 85 | M=13.25 years of education (SD=2.58) | Monthly income M=$906 (SD=$814) USD | 57.5% never married | Legal issues: 32.5% involved with legal system; Excluded those with imminent incarceration.<br>Mental health: Excluded those with active psychosis, severe psychiatric/medical disorders that would impede attendance for outpatient counselling. |
| Budney 2006[11] | M=33.1 (SD=10.3), range ≥18 [inclusion criterion] | United States | 96% White | 58% employed full-time | 77 | M=12.83 years of education (SD=4.15) | Monthly income M=$1294 (SD=1273) USD | 58% never married | Legal issues: 32% of participants involved with the |

| Study ID | Age (years) | Place of residence | Race, ethnicity, culture, language | Occupation | Gender/sex (% male) | Education | Socio-economic Status | Social capital | Other PROGRESS details |
|---|---|---|---|---|---|---|---|---|---|
| | | Excluded those with unstable living situation. | | | | | | | legal system. Excluded those with imminent incarceration.<br>Mental health: Excluded those with active psychosis/ severe medical or psychiatric disorder. |
| Carroll 2006[6] | M=21 (SD=2.1), range 18-25 [inclusion criterion] | United States | 60% African American, 13% Latin American, 23% European American | 21% employed full-time job, 28% employed part-time, 51% unemployed | 90 | 35% high school graduates, 18% completed some college-level work, 48% did not complete high school | NR | 96% never married | Legal issues: All referred by probation service.<br>Mental health: Excluded those who required inpatient treatment, had current psychotic disorder or severe medical problems, or had a risk of homicide. |
| Carroll 2012[12] | M=25.7 (SD=7.1), range ≥18 [inclusion criterion] | United States | 18.9% Caucasian, 63.8% African American, 12.6% Hispanic, 0.8% Asian, 3.9% Biracial | 40.9% unemployed | 84.3 | 62.2% completed high school | NR | 90.6% never married/ living alone | Legal issues: 93.7% referred by criminal justice system. Excluded those where imminent incarceration was likely.<br>Mental health: Excluded those who required inpatient treatment |
| Copeland 2001[21] | M=32.3 (SD=7.9), range 18-59 | Australia | 3.1% Aboriginal or Torres Strait Islander | 63.8% full-time/self-employment, 17.0% part-time/casual employment, 11.8% unemployed, 4.8% government benefit/ pension, 5.2% student* | 69.4* | 57.6% completed secondary school, 42.3% did not complete secondary school, 22.7% university qualification;<br>All English-literate for inclusion | NR | 47.9% married/ co-habiting, 16.6% with partner, not co-habiting, 9.2% separated/ divorced, 26.6% single/ never married* | Mental health: Excluded participants with DSM-IV Axis I diagnoses if symptoms were not currently stable or sufficiently mild as to cause no impact on their ability to participate |
| Davoudi 2021a[18] | M=26.41 (SD=6.65), range 18-45 | Iran | NR | NR | 100 | 13% no higher education, 48% diploma, 39% | NR | NR | Mental health: Excluded those with current or past |

| Study ID | Age (years) | Place of residence | Race, ethnicity, culture, language | Occupation | Gender/sex (% male) | Education | Socio-economic Status | Social capital | Other PROGRESS details |
|---|---|---|---|---|---|---|---|---|---|
| | | | | | | university student or graduate;<br>At least secondary education for inclusion | | | history of major psychiatric disorders |
| Davoudi 2021b[19] | M=25.85 (SD=4.99), range 18-45 [inclusion criterion] | Iran | NR | NR | 100 | 8% below high-school diploma level, 50% achieved high school diploma, 42% university student or graduate;<br>A minimum of a secondary school diploma for inclusion | NR | NR | Mental health: Included only those with a score of at least 13 on the Beck Depression and Anxiety questionnaire |
| Hoch 2014[22] | M=26.6 (SD=8.2), range 16-63 | Germany | Fluent in the German language for inclusion | 33.3% employed, 19.6% at school | 86.7 | Secondary educational attainment: 26.7% Hauptschule (vocational secondary school), 33.3% Realschule, Fachschule (intermediate secondary school), 7.9% Fachhoch-schulreife (advanced vocational certificate); 19.2% Abitur (school leaving examination), 11.7% school dropout, 1.3% other, 19.6% currently at school | Social class: 17.4% lower, 27.2% lower middle, 45.3% middle, 9.1% upper middle, 1.1% upper;<br>Income: 46.5% salary, 3% partner's salary, 1.5% pension, 0.7% property income, 32.1% unemployment compensation, 7.8% apprenticeship, 29.5% other | 86% not married | Mental health: Excluded those with severe major depression, panic disorder, psychotic disorders or suicidal tendencies.<br>Learning disability: Excluded those with learning disability/pervasive developmental disorder. |

| Study ID | Age (years) | Place of residence | Race, ethnicity, culture, language | Occupation | Gender/sex (% male) | Education | Socio-economic Status | Social capital | Other PROGRESS details |
|---|---|---|---|---|---|---|---|---|---|
| Kadden 2007[13] | M=32.7 (SD=9.6), range ≥18 [inclusion criterion] | United States<br><br>Excluded those with lack of reliable transport to the treatment site, or excessive commuting distance | 60% White | 73% employed | 71 | M=13 years of education (SD= 1.8);<br>Reading ability at least at fifth grade level for inclusion | NR | 45.4% married | Legal issues: Excluded those with legal problems.<br>Mental health: Excluded those with acute medical or psychiatric problems that required inpatient treatment (e.g., acute psychosis, or serious suicide/homicide risk). |
| Kaminer 2017[2] | M=16.11 (SD NR), range 13-18 | United States | Race: 77% Caucasian, 16% Black or African American, 2% Asian, 1% American Indian/Alaska Native, 4% More than one race;<br>Ethnicity: 32% Hispanic or Latino, 68% not Hispanic or Latino* | NR | 83* | Comprehend and read English at a fifth-grade level for inclusion | NR | NR | Mental health: Excluded those suicidal in the past 30 days, and those with any medical conditions compromising the ability to regularly participate in the study, including anyone with schizophrenia |
| Khalily 2023[23] | M=24.7 (SD=3.4), range 18-30 [inclusion criterion] | Pakistan<br>Excluded those who were unable to engage in study procedures at the study site due to transport difficulties | Indigenously adapted intervention, conducted using Urdu | 24% student, 6% government employee, 9% private, 61% self-employee | 95 | 24% primary, 48% secondary, 28% graduation | Socioeconomic status: 26% low, 61% middle, 13% high | 55% joint family, 45% nuclear family; 28% married, 70% unmarried, 3% divorced | NR |
| Litt 2013[14] | M=32.7 (SD=10), range ≥18 [inclusion criterion] | United States<br>Excluded those with lack of reliable transportation to treatment site, or lack of stable residence | 68.1% White, 2.2% Black, 16.4% Hispanic, 3.3% other | 74.9% employed full or part-time | 68 | M=13.2 years of education (SD=2.2);<br>Excluded those who had tested reading ability below the fifth grade level | NR | 34.9% married or co-habiting | NR |

| Study ID | Age (years) | Place of residence | Race, ethnicity, culture, language | Occupation | Gender/sex (% male) | Education | Socio-economic Status | Social capital | Other PROGRESS details |
|---|---|---|---|---|---|---|---|---|---|
| Litt 2020[15] | M=36 (SD=12), range ≥18 [inclusion criterion] | United States<br>Excluded those with lack of reliable transport or excessive commute | 51% White, 28% Black, 14% Hispanic, 7% Other | 60% employed full or part-time | 58 | M=13.7 years of education (SD=5.8) | 75% with income ≥$10,000 USD per year | 20% living with spouse/ partner | Mental health: Excluded those who required inpatient treatment |
| NCT02102230 2014[5] | M=48.34 (SD=15.83), range 19-64 | United States | Race: 61.7% White, 20% Black or African American, 15% unknown or not reported, 1.7% American Indian or Alaska Native, 1.7% Asian, 0% Native Hawaiian or Other Pacific Islander or More than one race;<br>Ethnicity: 10% Hispanic or Latino, 81.7% Not Hispanic or Latino, 8.3% unknown or NR | All veterans | 95 | NR | NR | NR | Mental health: Excluded those with history of, or current, psychotic symptoms, active suicidal/homicidal intent.<br>Reproductive health: Excluded those with current pregnancy. |
| Rigter 2013[3] | M=16.3 (SD=1.2), range 13-18 [inclusion criterion] | Belgium, France, Germany, Netherlands, Switzerland | Excluded those with inability to speak and read the local language | NR | 85 | 75% attended school;<br>Excluded those with IQ <70 | NR | 89.7% living with family | Mental health: Excluded those with mental disorders requiring inpatient treatment |
| Stanger 2009[4] | M=16 (SD=1.05), range 12-18 [inclusion criterion] | United States | 91.3% Caucasian, 5.8% African American, 2.9% Hispanic | NR | 82.6 | | M=7.05 (SD=1.59) on Hollingshead scale | All living with parent/ guardian (who agreed to participate) | Legal issues: 22% 'legal problems'.<br>Mental health: Excluded those with active psychosis, suicidal behaviour or severe medical illness. |
| Stephens 1994[17] | M=31.91 (SD NR), range 18-65 | United States | 95% White | 85% employed | 76 | 40% completed some college** | Annual income M=$20,220 (SD=$14,354) USD; 46% home ownership; 66% | 44% married | Mental health: Excluded those with psychosis |

| Study ID | Age (years) | Place of residence | Race, ethnicity, culture, language | Occupation | Gender/sex (% male) | Education | Socio-economic Status | Social capital | Other PROGRESS details |
|---|---|---|---|---|---|---|---|---|---|
| | | | | | | | ability to pay bills | | |
| Stephens 2000[16] | M=34 (SD=6.85), range | United States | 95% Caucasian* | 76% employed full time* | 77* | M=14 years of education (SD=2.80)* | NR | 55% single* | Mental health: Excluded those with severe psychological distress (suicidal intentions or psychotic thought process) |
| Wolitzky-Taylor 2022[7] | M=22.16 (SD=1.98), range 18–25 [inclusion criterion] | United States | 42% White, 4% Black, 17% Asian, 6% Pacific Islander, 6% Multi-racial, 25% Hispanic/Latino; fluent in English for inclusion | NR | 57.7 | NR | NR | NR | Learning disability: Excluded those with marked cognitive impairment.<br>Mental health: Excluded those with unstable manic or psychotic symptoms, and those with severe suicidality. Included only those with score >1 SD above the norm on the Positive and Negative Affect Scale-Negative Affect Subscale and > 1 SD above the norm on either the Anxiety Sensitivity Index, the Distress Tolerance Scale, or the suppression subscale of the Emotion Regulation Questionnaire. |

Note: No study provided information regarding religion.

* Characteristic is only reported for the entire cohort of randomized participants. For the purposes of the review, only some groups of this trial were eligible for inclusion.

** Characteristic is only reported for those who completed the trial.

M, mean; NR, not reported; SD, standard deviation; USD, United States dollars.
