## Supplementary material for "Effectiveness and safety of psychosocial interventions for the treatment of cannabis use disorder: a systematic review and meta-analysis": SuppInfo09

### SUPPORTING INFORMATION 9. RISK OF BIAS ASSESSMENT

The main sources of bias were: a lack of any pre-specified analysis plan to allow us to assess whether results were selected among multiple analyses/measures; high proportions of missing data that were likely to depend on the true value of the outcome (e.g., due to participant relapse); the use of self-reported data gathered from participants who were likely aware of their group allocation; a lack of information regarding the randomization and allocation concealment and baseline differences between intervention groups suggesting a problem with randomization; and a lack of clarity regarding the number of participants who were randomized to each intervention and/or included in analyses. The detailed assessments for each Risk of Bias 2 domain are presented in Table 1. Numbering of tables is specific to this Supporting Information document. References relating to this Supporting Information are included at the end of this document.

**Table 1.** Risk of Bias 2 assessment for each study and outcome at the end of treatment

| Study | Interventions | Outcome | Domain 1 | Domain 2 | Domain 3 | Domain 4 | Domain 5 | Overall | Comments |
|---|---|---|---|---|---|---|---|---|---|
| Babor 2004[1] | 1. Wait<br>2. MET/CBT | Continuous abstinence at end of treatment | Low | Low | High | High | Some concerns | High | D3: Some missing data and likely that it could depend on the true value of the outcome.<br>D4: The outcome was self-reported and participants were likely aware of their group allocation.<br>D5: No pre-specified analysis plan. |
| | | Completion of treatment | Low | Low | Low | Low | Low | Low | |
| | | Frequency of cannabis use at end of treatment | Low | Low | Low | High | Some concerns | High | D4: The outcome was self-reported and participants were likely aware of their group allocation.<br>D5: No pre-specified analysis plan. |
| | | Quantity of cannabis use at end of treatment | Low | Low | High | High | Some concerns | High | D3: Some missing data and likely that it could depend on the true value of the outcome.<br>D4: The outcome was self-reported and participants were likely aware of their group allocation.<br>D5: No pre-specified analysis plan. |
| Buckner 2019[2] | 1. MET/CBT<br>2. ICART | Point abstinence at end of treatment | Low | Some concerns | High | Low | Low | High | D2: Number of participants randomized to each arm is uncertain, therefore analysis approach is unclear.<br>D3: Some missing data and likely that it could depend on the true value of the outcome. |
| | | Completion of treatment | Low | Some concerns | Low | Low | Low | Some concerns | D2: Number of participants randomized to each arm is uncertain, therefore analysis approach is unclear. |
| | | Quantity of cannabis use at end of treatment | Low | Some concerns | High | High | Low | High | D2: Number of participants randomized to each arm is uncertain, therefore analysis approach is unclear.<br>D3: Some missing data and likely that it could depend on the true value of the outcome. |

| Study | Interventions | Outcome | Domain 1 | Domain 2 | Domain 3 | Domain 4 | Domain 5 | Overall | Comments |
|---|---|---|---|---|---|---|---|---|---|
| | | | | | | | | | D4: The outcome was self-reported and participants were likely aware of their group allocation. |
| Budak 2024[3] | 1. Wait<br>2. Mind/Edu | Completion of treatment | High | Low | Low | Low | Low | High | D1: Descriptions of randomization procedure throughout the manuscript are conflicting, and limited information regarding allocation process. |
| Budney 2000[4] | 1. MET/BT<br>2. MET/BT/ CM-ab | Point abstinence at end of treatment | Low | Low | High | Low | Some concerns | High | D3: Limited information regarding missing data and likely that it could depend on the true value of the outcome.<br>D5: No pre-specified analysis plan. |
| | | Continuous abstinence at end of treatment | Low | Low | High | Low | Some concerns | High | D3: Limited information regarding missing data and likely that it could depend on the true value of the outcome.<br>D5: No pre-specified analysis plan |
| | | Completion of treatment | Low | Low | Low | High | Low | High | D4: Outcome measured as the number of participants who attended at least one treatment session and gave a urine specimen during the last two weeks of the trial. Inappropriate for this outcome. |
| | | Duration of continuous abstinence at end of treatment | Low | Low | High | Low | Some concerns | High | D3: Some missing data and likely that it could depend on the true value of the outcome.<br>D5: No pre-specified analysis plan. |
| | | Frequency of cannabis use at end of treatment | Low | Low | High | High | Some concerns | High | D3: Some missing data and likely that it could depend on the true value of the outcome.<br>D4: The outcome was self-reported and participants were likely aware of their group allocation.<br>D5: No pre-specified analysis plan. |
| Budney 2006[5] | 1. CM-ab<br>2. MET/CBT/ CM-at<br>3. MET/CBT/ CM-ab | Point abstinence at end of treatment | Low | Low | High | Low | Some concerns | High | D3: Some missing data and likely that it could depend on the true value of the outcome.<br>D5: No pre-specified analysis plan. |
| | | Continuous abstinence at end of treatment | Low | Low | High | Low | Some concerns | High | D3: Some missing data and likely that it could depend on the true value of the outcome.<br>D5: No pre-specified analysis plan. |
| | | Completion of treatment | Low | Low | Low | Low | Low | Low | |
| | | Duration of continuous abstinence at end of treatment | Low | Low | High | Low | Some concerns | High | D3: Some missing data and likely that it could depend on the true value of the outcome.<br>D5: No pre-specified analysis plan. |
| | | Frequency of cannabis use at end of treatment | Low | Low | High | High | Some concerns | High | D3: Some missing data and likely that it could depend on the true value of the outcome.<br>D4: The outcome was self-reported and participants were likely aware of their group |

| Study | Interventions | Outcome | Domain 1 | Domain 2 | Domain 3 | Domain 4 | Domain 5 | Overall | Comments |
|---|---|---|---|---|---|---|---|---|---|
| | | | | | | | | | allocation.<br>D5: No pre-specified analysis plan. |
| Carroll 2006[6] | 1. NS<br>2. CM-ab-at/ NS<br>3. MET/CBT<br>4. MET/CBT/ CM-ab-at | Completion of treatment | Some concerns | Low | Low | Low | Low | Some concerns | D1: Limited information regarding randomization and allocation process. |
| | | Duration of continuous abstinence at end of treatment | Some concerns | Low | High | Low | Some concerns | High | D1: Limited information regarding randomization and allocation process<br>D3: Some missing data and likely that it could depend on the true value of the outcome.<br>D5: No pre-specified analysis plan. |
| | | Frequency of cannabis use at end of treatment | Some concerns | Low | High | Low | Some concerns | High | D1: Limited information regarding randomization and allocation process<br>D3: Some missing data and likely that it could depend on the true value of the outcome.<br>D5: No pre-specified analysis plan. |
| Carroll 2012[7] | 1. CM-ab<br>2. MET/CBT<br>3. MET/CBT/ CM-at<br>4. MET/CBT/ CM-ab | Completion of treatment | Low | Low | Low | Low | Low | Low | |
| | | Duration of continuous abstinence at end of treatment | Low | Low | High | High | Some concerns | High | D3: Some missing data and likely that it could depend on the true value of the outcome.<br>D4: The outcome was self-reported and participants were likely aware of their group allocation.<br>D5: Insufficient detail in the trial registration to know whether this analysis was pre-specified. |
| | | Frequency of cannabis use at end of treatment | Low | Low | High | High | Low | High | D3: Some missing data and likely that it could depend on the true value of the outcome.<br>D4: The outcome was self-reported and participants were likely aware of their group allocation. |
| Davoudi 2021a[8] | 1. NS<br>2. DBT | Point abstinence at end of treatment | Some concerns | Low | High | Low | Some concerns | High | D1: Limited information regarding allocation process.<br>D3: Some missing data and likely that it could depend on the true value of the outcome.<br>D5: Only retrospective trial registration available. Some inconsistencies with the paper. |
| | | Completion of treatment | Some concerns | Low | Low | Low | Low | Some concerns | D1: Limited information regarding allocation process. |
| | | Frequency of cannabis use at end of treatment | Some concerns | Low | High | Low | Some concerns | High | D1: Limited information regarding allocation process.<br>D3: Some missing data and likely that it could depend on the true value of the outcome.<br>D5: Only retrospective trial registration available. Some inconsistencies with the paper. |
| | | Cravings at end of treatment | Some concerns | Low | High | Low | Some concerns | High | D1: Limited information regarding allocation process.<br>D3: Some missing data and likely that it could depend on the true value of the outcome. |

| Study | Interventions | Outcome | Domain 1 | Domain 2 | Domain 3 | Domain 4 | Domain 5 | Overall | Comments |
|---|---|---|---|---|---|---|---|---|---|
| | | | | | | | | | D5: Only retrospective trial registration available. Some inconsistencies with the paper. |
| Davoudi 2021b[9] | 1. NS<br>2. ACT | Point abstinence at end of treatment | Some concerns | Low | Low | Low | Some concerns | Some concerns | D1: Limited information regarding allocation process, and baseline differences between intervention groups.<br>D5: Only retrospective trial registration available. Some inconsistencies with the paper. |
| | | Completion of treatment | Some concerns | Low | Low | Low | Low | Some concerns | D1: Limited information regarding allocation process, and baseline differences between intervention groups. |
| | | Frequency of cannabis use at end of treatment | Some concerns | Some concerns | Some concerns | Some concerns | Some concerns | Some concerns | D1: Limited information regarding allocation process, and baseline differences between intervention groups.<br>D2: Appears that a per protocol analysis was used, but judged unlikely to have substantial impact on the results.<br>D3: Some missing data but reasons for exclusion provided - unlikely that missingness depended on the true value of outcome.<br>D4: Outcome self-reported but it is likely that some blinding was attempted.<br>D5: Only retrospective trial registration available. Some inconsistencies with the paper. |
| Hoch 2014[10] | 1. Wait<br>2. MET/CBT | Point abstinence at end of treatment | Low | Low | High | Some concerns | High | High | D3: Limited information regarding missing data and likely that it could depend on the true value of the outcome.<br>D4: Limited information regarding measurement methods.<br>D5: Discrepancy between the trial registration and the article in the definition of outcome. |
| Kadden 2007[11] | 1. NS<br>2. CM-ab<br>3. MET/CBT<br>4. MET/CBT/ CM-ab | Continuous abstinence at end of treatment | Low | Low | Low* | High | Some concerns | High | D4: The outcome was self-reported and participants were likely aware of their group allocation.<br>D5: No pre-specified analysis plan. Unclear why self-report was preferred over urine samples. |
| | | Completion of treatment | Low | Low | Low | Low | Low | Low | |
| | | Duration of continuous abstinence at end of treatment | Low | Low | High | High | Some concerns | High | D3: Some missing data and likely that it could depend on the true value of the outcome.<br>D4: The outcome was self-reported and participants were likely aware of their group allocation.<br>D5: No pre-specified analysis plan. Unclear why self-report was preferred over urine samples. |
| | | Frequency of cannabis use at end of treatment | Low | Low | High | High | Some concerns | High | D3: Some missing data and likely that it could depend on the true value of the outcome.<br>D4: The outcome was self-reported and participants were likely aware of their group allocation.<br>D5: No pre-specified analysis plan. |

| Study | Interventions | Outcome | Domain 1 | Domain 2 | Domain 3 | Domain 4 | Domain 5 | Overall | Comments |
|---|---|---|---|---|---|---|---|---|---|
| Kaminer 2017[12] | 1. MET/CBT<br>2. ComReinf | Point abstinence at end of treatment | Some concerns | Low | High | Low | Low | High | D1: Limited information regarding allocation process, and baseline differences between intervention groups.<br>D3: Some missing data and likely that it could depend on the true value of the outcome. |
| | | Completion of treatment | Some concerns | Low | Low | Low | Low | Some concerns | D1: Limited information regarding allocation process, and baseline differences between intervention groups. |
| Khalily 2023[13] | 1. NS<br>2. ComReinf | Continuous abstinence at end of treatment | Low | Low | Low | High | High | High | D4: The outcome was self-reported and participants were likely aware of their group allocation.<br>D5: No pre-specified analysis plan. The result could have been selected from multiple eligible outcome measurements. |
| | | Completion of treatment | Low | Low | Low | Low | High | High | D5: No pre-specified analysis plan. The result could have been selected from multiple eligible outcome measurements. |
| Litt 2013[14] | 1. NS<br>2. MET/CBT/ CM-ab<br>3. MET/CBT/ CM-at | Continuous abstinence at end of treatment | Low | Some concerns | High | High | Some concerns | High | D2: Number of participants included in analysis unclear.<br>D3: Some missing data and likely that it could depend on the true value of the outcome.<br>D4: The outcome was self-reported and participants were likely aware of their group allocation.<br>D5: No pre-specified analysis plan. Unclear why self-report was preferred over urine samples. |
| | | Duration of continuous abstinence at end of treatment | Low | Some concerns | High | High | Some concerns | High | D2: Number of participants included in analysis unclear.<br>D3: Some missing data and likely that it could depend on the true value of the outcome.<br>D4: The outcome was self-reported and participants were likely aware of their group allocation.<br>D5: No pre-specified analysis plan. Unclear why self-report was preferred over urine samples. |
| | | Frequency of cannabis use at end of treatment | Low | Some concerns | High | High | Some concerns | High | D2: Number of participants included in analysis unclear.<br>D3: Some missing data and likely that it could depend on the true value of the outcome.<br>D4: The outcome was self-reported and participants were likely aware of their group allocation.<br>D5: No pre-specified analysis plan. |
| Litt 2020[15] | 1. MET/CBT<br>2. MET/CBT/ CM-ab<br>3. IATP<br>4. IATP/CM-ab | Continuous abstinence at end of treatment | Low | Low | Some concerns | Some concerns | Some concerns | Some concerns | D3: Some missing data which could depend on the true value of outcome. There was a larger proportion of events relative to missing data, but we considered that missing data could still potentially impact the outcome.<br>D4: Outcome assessment was unblinded (self-reported) but considered unlikely to influence the outcome based on urine verification.<br>D5: No pre-specified analysis plan. |
| | | Completion of treatment | Low | Low | Low | Low | Low | Low | |

| Study | Interventions | Outcome | Domain 1 | Domain 2 | Domain 3 | Domain 4 | Domain 5 | Overall | Comments |
|---|---|---|---|---|---|---|---|---|---|
| | | Duration of continuous abstinence at end of treatment | Low | Low | High | Some concerns | Some concerns | High | D3: Some missing data and likely that it could depend on the true value of the outcome.<br>D4: Outcome assessment was unblinded (self-reported) but interventions are all of similar intensity, and there may be no pre-conceived notion regarding superiority.<br>D5: No pre-specified analysis plan. |
| | | Frequency of cannabis use at end of treatment | Low | Low | High | Some concerns | Some concerns | High | D3: Some missing data and likely that it could depend on the true value of the outcome.<br>D4: Outcome assessment was unblinded (self-reported) but interventions are all of similar intensity, and there may be no pre-conceived notion regarding superiority.<br>D5: No pre-specified analysis plan. |
| NCT02102230 2014[16] | 1. NS<br>2. CBT-I<br>3. CBT-I | Point abstinence at end of treatment | Some concerns | High | High | Some concerns | High | High | D1: Limited information regarding randomization and allocation process.<br>D2: Trial terminated early. Unclear why data not available for all randomized participants.<br>D3: Some missing data and likely that it could depend on the true value of the outcome.<br>D4: Outcome assessment was unblinded (self-reported) but interventions are all of similar intensity, and there may be no pre-conceived notion regarding superiority.<br>D5: Discrepancy between statistical analysis plan and the article in the definition of outcome. Abstinence measured by self-report and urine testing. However, only self-reported outcome available. |
| | | Completion of treatment | Some concerns | High | Low | Low | Low | High | D1: Limited information regarding randomization and allocation process.<br>D2: Trial terminated early. Unclear why data not available for all randomized participants. |
| | | Adverse events | Some concerns | High | High | Some concerns | Some concerns | High | D1: Limited information regarding randomization and allocation process.<br>D2: Trial terminated early. Unclear why data not available for all randomized participants.<br>D3: High proportion of missing data. No information regarding those who did not complete treatment.<br>D4: Limited information regarding measurement methods.<br>D5: No mention of this outcome in the statistical analysis plan. |
| | | Frequency of cannabis use at end of treatment | Some concerns | High | High | Some concerns | Some concerns | High | D1: Limited information regarding randomization and allocation process.<br>D2: Trial terminated early. Unclear why data not available for all randomized participants.<br>D3: Some missing data and likely that it could depend on the true value of the outcome.<br>D4: Outcome assessment was unblinded (self-reported) but interventions are all of similar intensity, and there may be no pre-conceived notion regarding superiority.<br>D5: Discrepancy between statistical analysis plan and the article in the definition of outcome. |

| Study | Interventions | Outcome | Domain 1 | Domain 2 | Domain 3 | Domain 4 | Domain 5 | Overall | Comments |
|---|---|---|---|---|---|---|---|---|---|
| Rigter 2013[17] | 1. MET/CBT<br>2. MDFT | Completion of treatment | Some concerns | Low | Low | Low | Some concerns | Some concerns | D1: There is some information about allocation concealment but there is a risk that a sequence could be predicted due to very small block size.<br>D5: Data at the end of the treatment period are not fully reported. |
| | | Frequency of cannabis use at end of treatment | Some concerns | Low | High | High | Some concerns | High | D1: There is some information about allocation concealment but there is a risk that a sequence could be predicted due to very small block size.<br>D3: Some missing data and likely that it could depend on the true value of the outcome.<br>D4: The outcome was self-reported and participants were likely aware of their group allocation.<br>D5: No pre-specified analysis plan. |
| Stanger 2009[18] | 1. MET/CBT/ CM-at/NS<br>2. MET/CBT/ CM-ab/NS | Point abstinence at end of treatment | Low | Low | Some concerns | Low | Some concerns | Some concerns | D3: Some missing data which could depend on the true value of outcome. However, the extent of missingness is considered to be smaller for the assessment of point abstinence outcome.<br>D5: No pre-specified analysis plan. |
| | | Continuous abstinence at end of treatment | Low | Low | High | Some concerns | Some concerns | High | D3: Some missing data and likely that it could depend on the true value of the outcome.<br>D4: Outcome assessors were not blinded and it is possible that the outcome was self-reported.<br>D5: No pre-specified analysis plan. |
| | | Completion of treatment | Low | Low | Low | Low | Low | Low | |
| | | Duration of continuous abstinence at end of treatment | Low | Low | High | Some concerns | Some concerns | High | D3: Some missing data and likely that it could depend on the true value of the outcome.<br>D4: Outcome assessors were not blinded and it is possible that the outcome was self-reported.<br>D5: No pre-specified analysis plan. |
| | | Frequency of cannabis use at end of treatment | Low | Low | Some concerns | High | Some concerns | High | D3: Some missing data which could depend on the true value of outcome. However, the extent is considered to be small.<br>D4: The outcome was self-reported and participants were likely aware of their group allocation.<br>D5: No pre-specified analysis plan. |
| | | Adverse events | Low | Low | Some concerns | Some concerns | Some concerns | Some concerns | D3: Some missing data which could depend on the true value of outcome. However, the extent is considered to be small.<br>D4: Outcome assessment was likely unblinded. No indication that knowledge of the intervention influenced reporting of adverse events (no events were reported).<br>D5: No pre-specified analysis plan. |
| Stephens 1994[19] | 1. NS<br>2. RelPrev | Continuous abstinence at end of treatment | Some concerns | Low | Low | Some concerns | Some concerns | Some concerns | D1: Limited information regarding allocation process, and baseline differences between intervention groups.<br>D4: Outcome assessment was unblinded (self-reported) but considered unlikely to |

| Study | Interventions | Outcome | Domain 1 | Domain 2 | Domain 3 | Domain 4 | Domain 5 | Overall | Comments |
|---|---|---|---|---|---|---|---|---|---|
| | | | | | | | | | influence the outcome based on collateral reports. D5: No pre-specified analysis plan. |
| | | Frequency of cannabis use at end of treatment | Some concerns | High | High | Some concerns | High | High | D1: Limited information regarding allocation process, and baseline differences between intervention groups. D2: Analysis did not include all randomized participants. D3: Some missing data and likely that it could depend on the true value of the outcome. D4: Outcome assessment was unblinded (self-reported) but considered unlikely to influence the outcome based on collateral reports. D5: No pre-specified analysis plan. Discrepancy between methods and results sections - outcome was not reported at stated timepoint. |
| Stephens 2000[20] | 1. Wait<br>2. RelPrev | Continuous abstinence at end of treatment | Some concerns | Some concerns | High | High | Some concerns | High | D1: Limited information regarding allocation process, and baseline differences between intervention groups. D2: Proportion of participants attending treatment outside of trial (higher in control than intervention). D3: Some missing data and likely that it could depend on the true value of the outcome. D4: The outcome was self-reported and participants were likely aware of their group allocation. D5: No pre-specified analysis plan. |
| | | Completion of treatment | Some concerns | Some concerns | Low | Low | Low | Some concerns | D1: Limited information regarding allocation process, and baseline differences between intervention groups. D2: Proportion of participants attending treatment outside of trial (higher in control than intervention). |
| | | Frequency of cannabis use at end of treatment | Some concerns | Some concerns | High | High | Some concerns | High | D1: Limited information regarding allocation process, and baseline differences between intervention groups. D2: Proportion of participants attending treatment outside of trial (higher in control than intervention). D3: Some missing data and likely that it could depend on the true value of the outcome. D4: The outcome was self-reported and participants were likely aware of their group allocation. D5: No pre-specified analysis plan. |
| Wolitzky-Taylor 2022[21] | 1. MET/CBT<br>2. AMT | Completion of treatment | Some concerns | Low | Low | Low | Low | Some concerns | D1: Limited information regarding allocation process. |
| | | Frequency of cannabis use at end of treatment | Some concerns | Low | High | High | High | High | D1: Limited information regarding allocation process. D3: Some missing data and likely that it could depend on the true value of the outcome. D4: The outcome was self-reported and participants were likely aware of their group allocation. D5: Pre-specified primary outcome measure was not reported. |

*In Kadden 2007, for continuous abstinence at end of treatment, the contrast between interventions MET/CBT and NS was rated as high risk for Domain 3. All remaining contrasts for this outcome were rated as Low risk for Domain 3, as indicated in the table.

Note that Copeland 2001 is not included in this table as it only reported outcomes at medium follow-up.

D1, risk of bias arising from the randomization process; D2, risk of bias due to deviations from intended interventions; D3, risk of bias due to missing outcome data; D4, risk of bias in measurement of the outcome; D5, risk of bias in selection of the reported result.

ACT, acceptance and commitment therapy; AMT, affect management therapy; BT, behavioural therapy; CBT, cognitive-behavioural therapy; CBT-I, CBT for insomnia; CM-ab/at, contingency management based on abstinence/attendance; ComReinf, community reinforcement; DBT, dialectical behavioural therapy; ICART, integrated cannabis and anxiety reduction treatment; MET, motivation enhancement therapy; Mind/Edu, mindfulness psychoeducation; NS, nonspecific comparator; RelPrev, relapse prevention; Wait, waitlist.
