## Supplementary material for "Effectiveness and safety of psychosocial interventions for the treatment of cannabis use disorder: a systematic review and meta-analysis": SuppInfo10

### SUPPORTING INFORMATION 10. RESULTS SYNTHESIS FOR OUTCOMES ASSESSED AT MEDIUM AND LONG FOLLOW-UP

In summary, the findings based on follow-up data suggest that cognitive-behavioural therapy (CBT) and dialectical behavioural/acceptance and commitment therapies (DBT/ACT) versus inactive/nonspecific comparators, CBT plus abstinence- versus attendance-based contingency management (CM), and community reinforcement versus other interventions, may benefit some abstinence outcomes several months post-treatment. Detailed results are presented below. Numbering of figures is specific to this Supporting Information document. References relating to this Supporting Information are included at the end of this document.

#### Point abstinence

Six studies assessed point abstinence at medium term follow-up.[1-6] Medium follow up times ranged from 2 to 6 months post-treatment. Meta-analyses included a maximum of two studies per comparison (Figure 1). The common between-study variance, $\tau^2$, across comparisons was estimated as 0.00 (standard error, SE=0.49). CBT (odds ratio, OR=2.29, 95% confidence interval, CI [0.78; 6.69]) and DBT/ACT (OR=5.19 [1.83; 14.67]) relative to inactive/nonspecific comparators, as well as CBT plus CM-abstinence relative to CBT plus CM-attendance (OR=3.93 [1.57; 9.82]), may increase point abstinence at medium follow-up. However, we note that CIs are very wide, and for CBT are consistent with both an increase and decrease in point abstinence. CBT plus CM-abstinence may result in a reduction in point abstinence compared with CBT alone (OR=0.62 [0.21; 1.88]), although the CIs are consistent with both an increase and decrease in point abstinence.

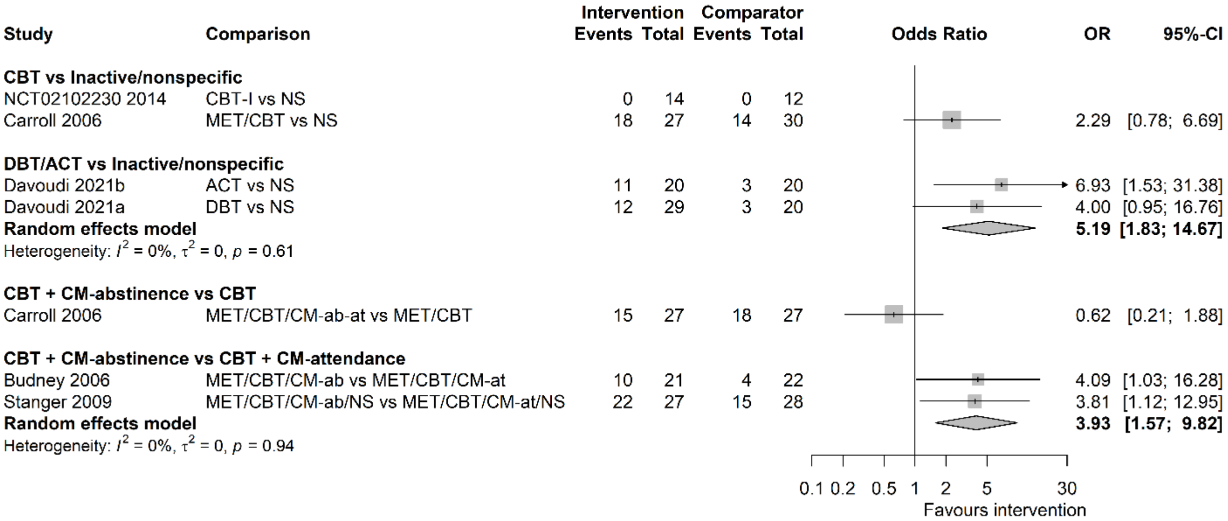


**Figure 1.** Forest plot for random-effects meta-analyses of point abstinence at medium follow-up.
ACT, acceptance and commitment therapy; CBT, cognitive-behavioural therapy; CBT-I, CBT for insomnia; CI, confidence interval; CM-ab/at, contingency management based on abstinence/attendance; DBT, dialectical behavioural therapy; MET, motivation enhancement therapy; NS, nonspecific comparator; OR, odds ratio.

Figure 2 presents the findings from a meta-analysis of two studies for point abstinence at long term follow-up that ranged from 9 to 12 months post-treatment.[5,6] The common $\tau^2$ was estimated as 0.00 (SE=0.5). CBT plus CM-abstinence may increase abstinence relative to CBT plus CM-attendance (OR=2.55 [1.12; 5.81]).

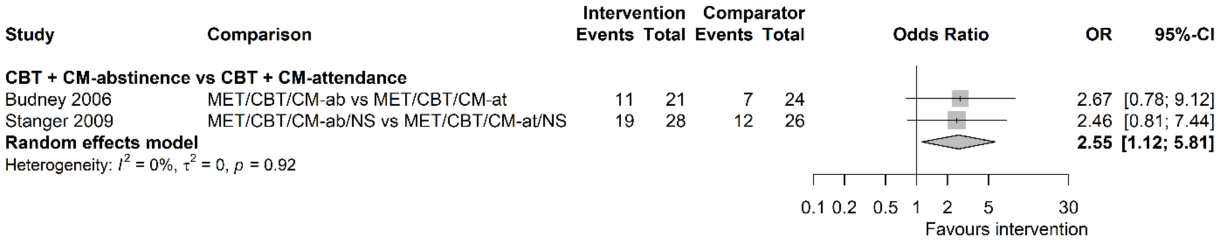


**Figure 2.** Forest plot for random-effects meta-analysis of point abstinence at long follow-up.
CBT, cognitive-behavioural therapy; CI, confidence interval; CM-ab/at, contingency management based on abstinence/attendance; MET, motivation enhancement therapy; NS, nonspecific comparator; OR, odds ratio.

### Continuous abstinence

Eight studies assessed rates of continuous abstinence at medium follow-up,[2,5,7-12] mostly at six months post-treatment (with a single study measuring abstinence at 4.5 months post-treatment). Continuous abstinence was measured over 1-6 months (median=3 months) prior to the follow-up assessment. Meta-analyses included a maximum of four studies per comparison (Figure 3). The common $\tau^2$ across comparisons was estimated as 0.37 (SE=0.39). There may be a favourable effect of CBT plus CM-abstinence over CBT plus CM-attendance (OR=2.85 [0.87; 9.30]), however CIs are consistent with both an increase or decrease in abstinence. For CBT relative to inactive/nonspecific comparators there was little to no evidence of a meaningful effect and CIs are also consistent with both an increase or decrease in abstinence (OR=1.87 [0.80; 4.35]. The same applies to the effect of CBT plus CM-abstinence versus CBT alone (OR=0.85 [0.36; 2.02]). For community reinforcement over nonspecific comparator there is evidence of an effect in favour of community reinforcement but CIs are very wide (OR=30.86 [10.27; 92.69]).

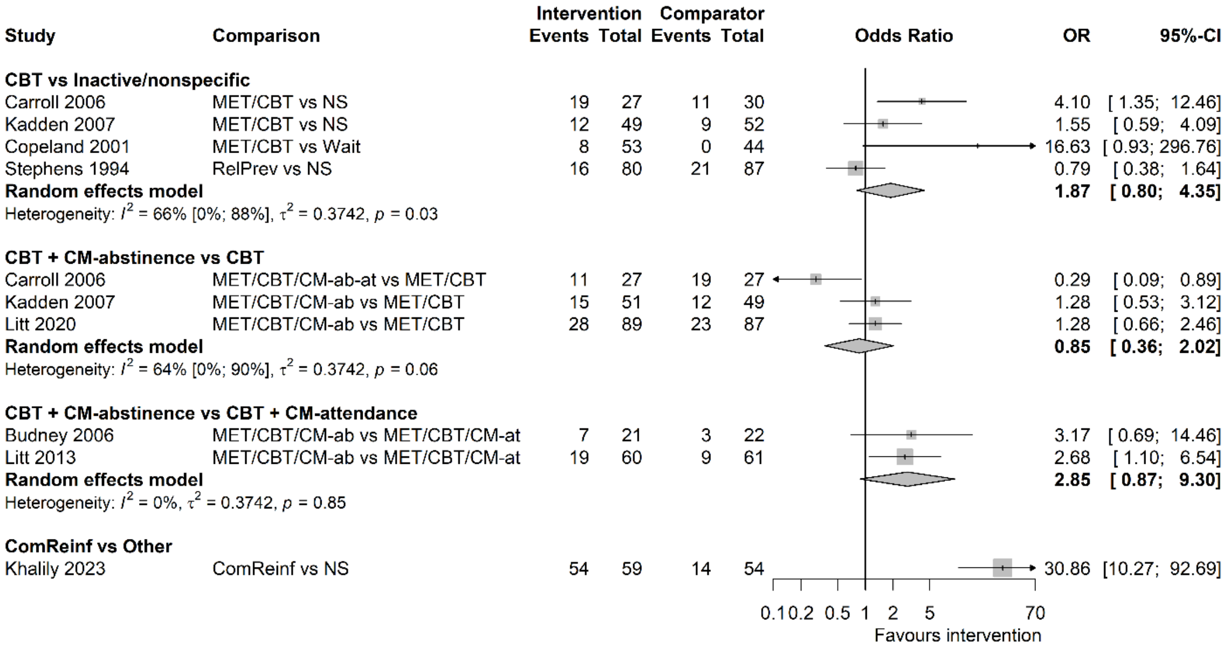


**Figure 3.** Forest plot for random-effects meta-analyses of continuous abstinence at medium follow-up. CBT, cognitive behavioural therapy; CI, confidence interval; CM-ab/at, contingency management based on abstinence/attendance; ComReinf, community reinforcement; MET, motivation enhancement therapy; NS, nonspecific comparator; OR, odds ratio; RelPrev, relapse prevention; Wait, waitlist

Six studies assessed continuous abstinence at long follow-up,[5,7,9-12] five at 12 months post-treatment and one at 7.5 months post-treatment (Khalily 2023).[12] Continuous abstinence was measured over 1-12 months (median=5.25 months) prior to the follow-up assessment. Meta-analyses included a maximum of two studies per comparison (Figure 4). The common $\tau^2$ across comparisons was 0.00 (SE=0.17). The effect of community reinforcement relative to a nonspecific comparator was favourable, suggesting an increase in continuous abstinence (OR=28.17 [9.72; 81.65]). For the remaining interventions the evidence suggests they may have little to no effect on continuous abstinence at long follow-up (CBT versus inactive/nonspecific comparators (OR=1.28 [0.67; 2.44]), CBT plus CM-abstinence versus CBT alone (OR=1.12 [0.66; 1.90]), and CBT plus CM-abstinence versus CBT plus CM-attendance (OR=1.69 [0.77; 3.70])).

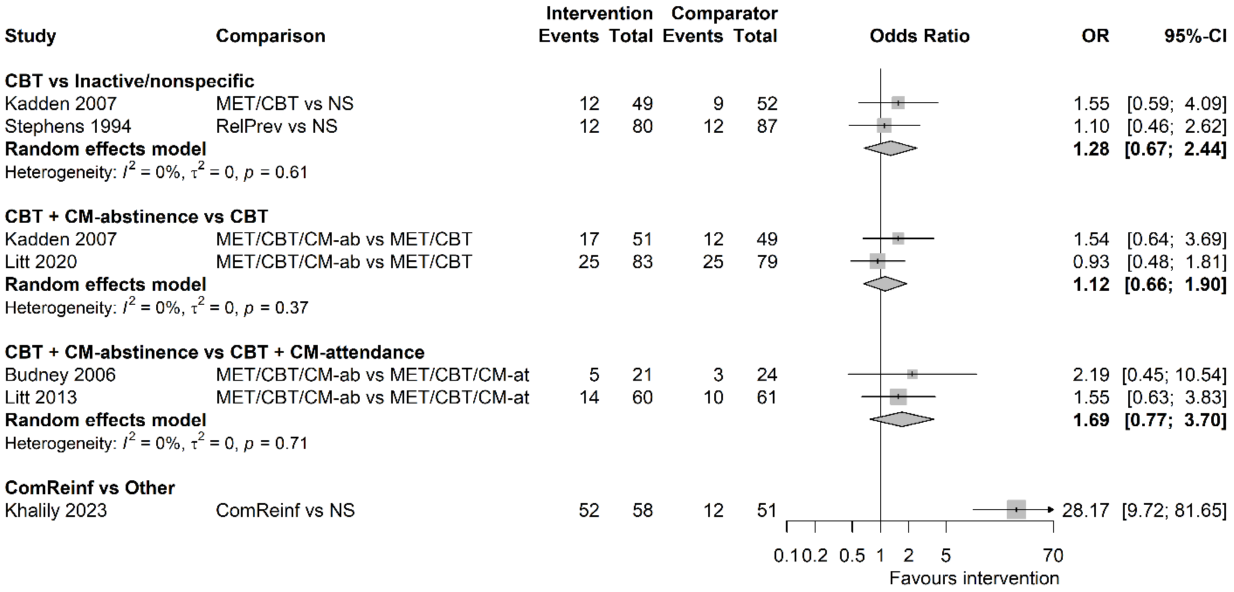


**Figure 4.** Forest plot for random-effects meta-analyses of continuous abstinence at long follow-up.
CBT, cognitive-behavioural therapy; CI, confidence interval; CM-ab/at, contingency management based on abstinence/attendance; ComReinf, community reinforcement; MET, motivation enhancement therapy; NS, nonspecific comparator; OR, odds ratio; RelPrev, relapse prevention; Wait, waitlist

.

### Duration of continuous abstinence

Eight studies reported mean duration of continuous abstinence within the treatment period lasting between 8 and 14 weeks (median=12 weeks), expressed either as number of weeks or days.[2,5-7,10,11,13,14] Most used self-reported abstinence, but three studies determined abstinence by consecutive weekly[2] or twice-weekly[5,14] negative urine tests. Meta-analyses shown in Figure 5 included up to five studies per comparison, with the common $\tau^2$ across the comparisons estimated as 0.05 (SE=0.06).

There is very low certainty evidence that CBT relative to nonspecific comparators (ratio of means, RoM=1.24, 95% CI [0.70; 2.19]), CBT plus CM-abstinence relative to CBT alone (RoM=1.40 [1.02; 1.91]), and CBT plus CM-abstinence relative to CBT plus CM-attendance (RoM=1.31 [0.97; 1.77]), may have little to no effect on the duration of continuous abstinence over the treatment period.

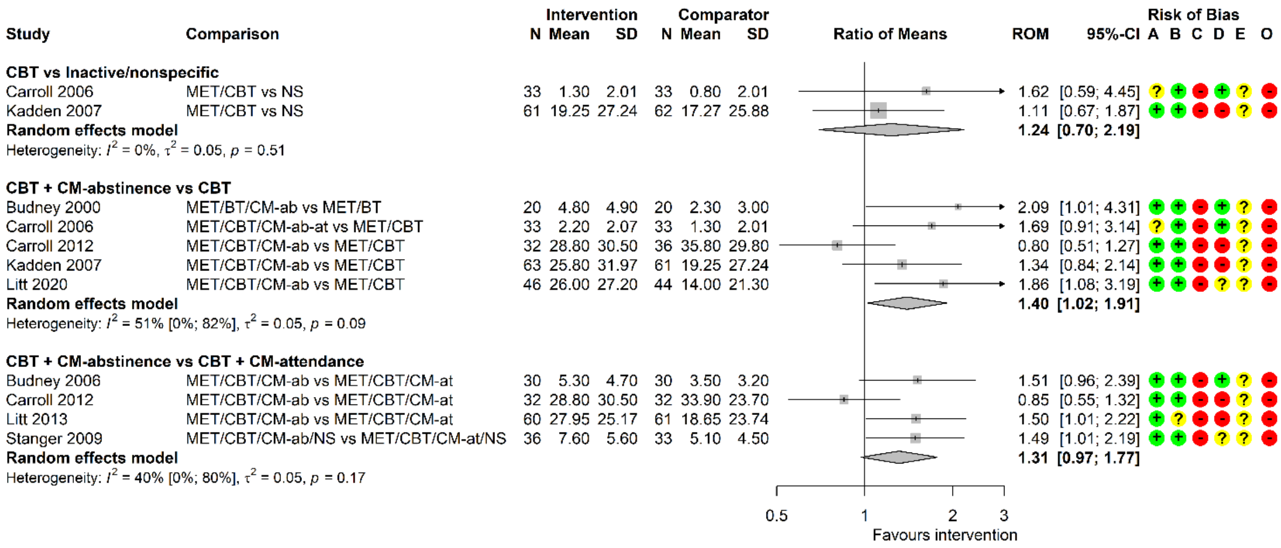


**Figure 5.** Forest plot for random-effects meta-analyses of duration of continuous abstinence at end of treatment. CBT, cognitive-behavioural therapy; CI, confidence interval; CM-ab/at, contingency management based on abstinence/attendance; MET, motivation enhancement therapy; N, number of participants; NS, nonspecific comparator; ROM, ratio of means, SD standard deviation.
Risk of bias (A) arising from the randomization process, (B) due to deviations from intended interventions, (C) due to missing outcome data, (D), in measurement of the outcome, (E) in selection of the reported result, (O) overall; '+', low risk, '?', some concerns, '-', high risk of bias.

No evidence was available for the duration of continuous abstinence at medium follow-up, however, a single study[7] assessed self-reported longest number of days of abstinence over a 12 months follow-up period. As shown in Figure 6, CBT relative to a nonspecific comparator (RoM=1.08 [0.71; 1.65]), as well as CBT plus CM-abstinence relative to CBT alone (RoM=1.17 [0.77; 1.79]), may have little to no effect on the duration of continuous abstinence at long follow-up.

| Study | Comparison | Intervention N | Intervention Mean | Intervention SD | Comparator N | Comparator Mean | Comparator SD | ROM | 95%-CI |
|---|---|---|---|---|---|---|---|---|---|
| **CBT vs Inactive/nonspecific** | | | | | | | | | |
| Kadden 2007 | MET/CBT vs NS | 55 | 103.13 | 122.14 | 54 | 95.49 | 103.54 | 1.08 | [0.71; 1.65] |
| **CBT + CM-abstinence vs CBT** | | | | | | | | | |
| Kadden 2007 | MET/CBT/CM-ab vs MET/CBT | 59 | 120.80 | 135.73 | 55 | 103.13 | 122.14 | 1.17 | [0.77; 1.79] |

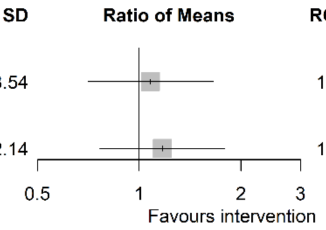


**Figure 6.** Forest plot for relative treatment effects on duration of continuous abstinence at long follow-up.
CBT, cognitive-behavioural therapy; CI, confidence interval; CM-ab, contingency management based on abstinence; MET, motivation enhancement therapy; N, number of participants; NS, nonspecific comparator; ROM, ratio of means; SD, standard deviation.

### Frequency of cannabis use

Fourteen studies reported frequency of cannabis use at medium follow-up.[1-11,13,15,16] Follow-up ranged from 2 to 6 months (median=6 months). Frequency of use was self-reported, and ranged from 1 week to 6 months prior to the follow-up assessment (median=1 month). Meta-analyses shown in Figure 7 included up to five studies per comparison, with the common $\tau^2$ across the comparisons estimated as 0.05 (SE=0.03). At medium follow-up, no intervention had a clinically meaningful effect on reducing frequency of cannabis use and CIs were consistent with both increase and decrease in frequency of cannabis use. CBT (RoM=0.94 [0.73; 1.22]) and DBT/ACT (RoM=0.54 [0.38; 0.75]) relative to inactive/nonspecific comparators, CBT-affect relative to CBT alone (RoM=1.28 [0.62; 2.63]), CBT plus CM-abstinence relative to CBT alone (RoM=1.03 [0.78; 1.35]), CBT plus CM-abstinence relative to CBT plus CM-attendance (RoM=0.87 [0.64; 1.19]), and multidimensional family therapy (MDFT) relative to CBT (RoM=0.80 [0.68; 0.95]).

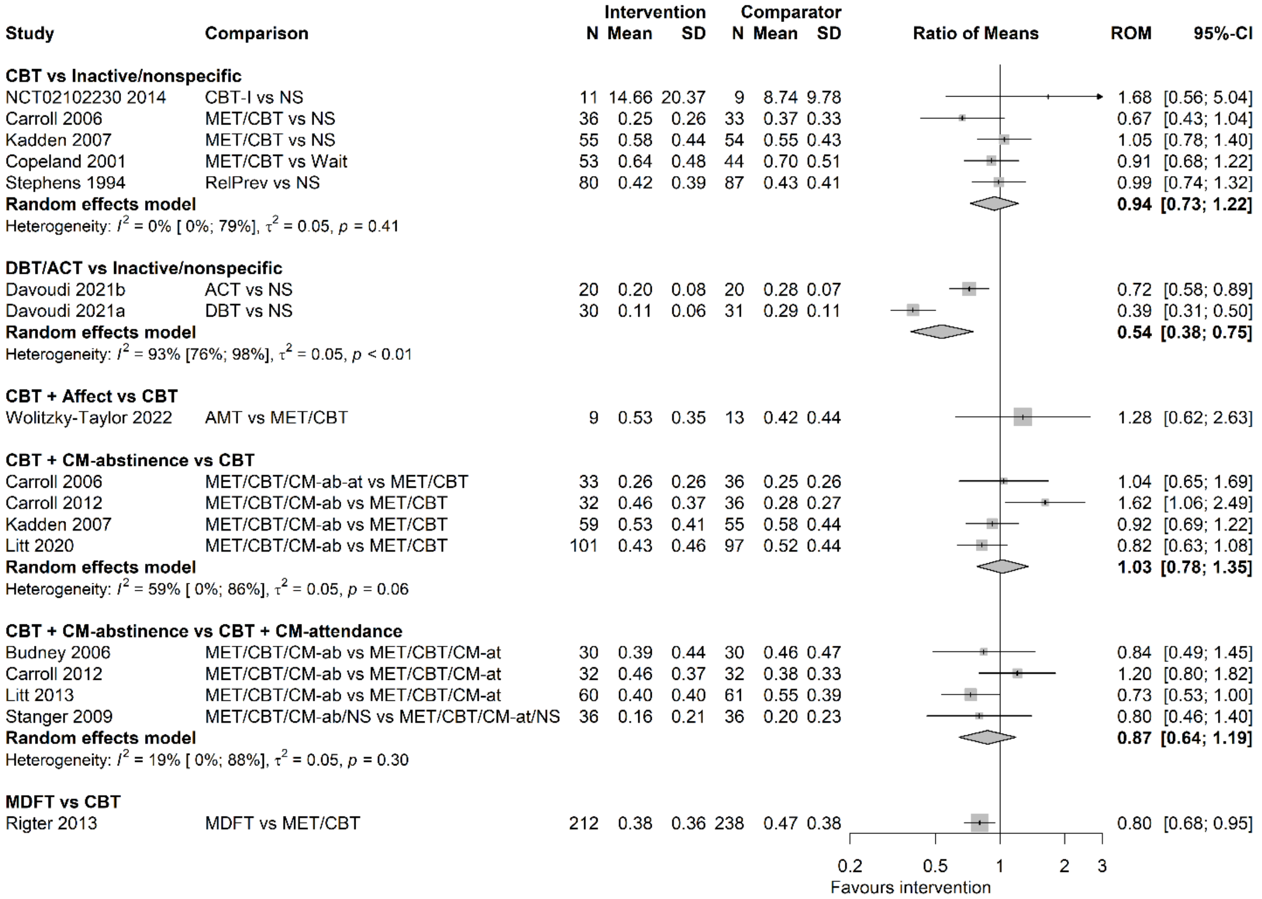


**Figure 7.** Forest plot for random-effects meta-analyses of frequency of cannabis use at medium follow-up. Frequency of use is expressed as proportion of days using for most studies, except for number of times used in the past 7 days in NCT02102230.

ACT, acceptance and commitment therapy; AMT, affect management therapy; CBT, cognitive-behavioural therapy; CBT-I, CBT for insomnia; CI, confidence interval; CM-ab/at, contingency management based on abstinence/attendance; DBT dialectical behavioural therapy; MDFT, multidimensional family therapy; MET, motivation enhancement therapy; N, number of participants; NS, nonspecific comparator; RelPrev, relapse prevention; ROM, ratio of means; SD, standard deviation; Wait, waitlist.

Seven studies reported frequency of cannabis use at long follow-up ranging from 9 to 13 months (median=12 months).[5-7,9-11,13] Frequency of use was self-reported over 1 to 3 months prior to the follow-up assessment (median=3 months). Meta-analyses shown in Figure 8 included no more than four studies per comparison, with the common $\tau^2$ across the comparisons estimated as <0.0001 (SE=0.02). At long follow-up, no intervention had a clinically meaningful effect, and probably had little to no effect on reducing frequency of cannabis use, including CBT relative nonspecific comparators (RoM=0.98 [0.81, 1.19]), CBT plus CM-abstinence versus CBT alone (RoM=0.97 [0.81; 1.17]), where CIs were consistent with both and increase and decrease in frequency of cannabis use, and CBT plus CM-abstinence versus CBT plus CM-attendance (RoM=0.80 [0.65; 0.98]).

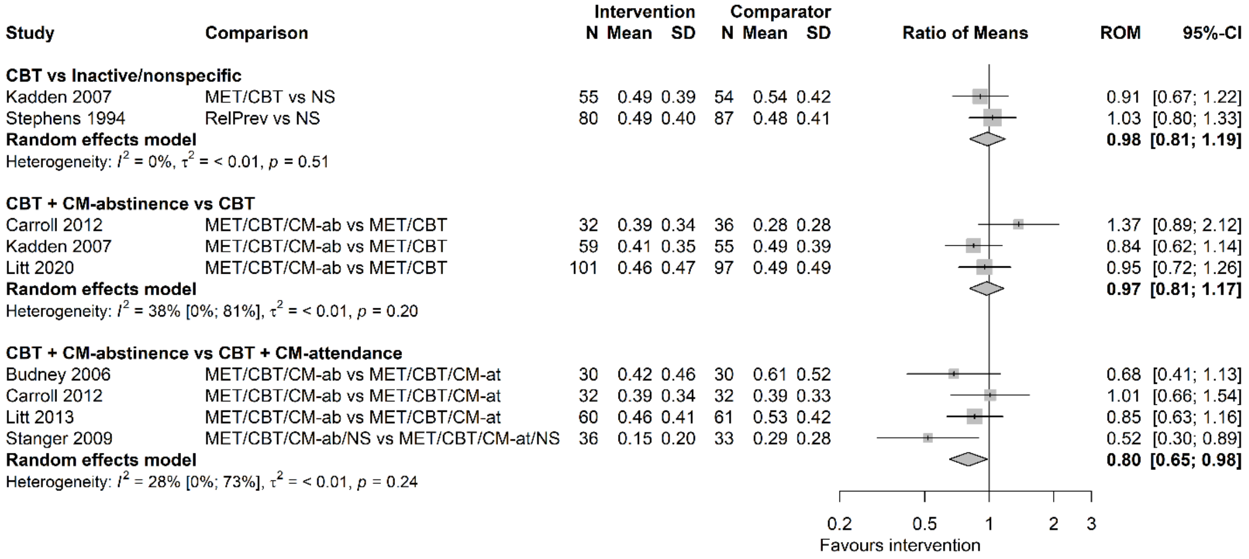


**Figure 8.** Forest plot for random-effects meta-analyses of frequency of cannabis use at long follow-up. Frequency of use is expressed as proportion of days using.
CBT, cognitive-behavioural therapy; CI, confidence interval; CM-ab/at, contingency management based on abstinence/attendance; MET, motivation enhancement therapy; N, number of participants; NS, nonspecific comparator; RelPrev, relapse prevention; ROM, ratio of means; SD, standard deviation; Wait, waitlist.

### Quantity of cannabis use

A single study[8] measured quantity of cannabis used at medium 6-months follow-up, expressed as self-reported number of waterpipes (cones) smoked per day over three days prior to the assessment. The authors considered three cones to be equivalent to one joint. Results suggest that there is probably little to no difference in effect between CBT and a waitlist control for reducing quantity of use (RoM=0.61 [0.48; 0.76]).

### Craving

One study[3] reported craving at medium 2-months follow-up, measured with Marijuana Craving Questionnaire short-form. Results suggested that DBT may have little to no effect on reducing craving relative to a nonspecific comparator (RoM=0.93 [0.84; 1.03]).
